# Cost per episode of diarrhea and respiratory syncytial virus (RSV) in 128 low- and middle-income countries: how well do disease-specific and WHO-CHOICE estimates align?

**DOI:** 10.1101/2024.07.17.24310217

**Authors:** Xiao Li, Joke Bilcke, Ernest O. Asare, Catherine Wenger, Jiye Kwon, Louis Bont, Philippe Beutels, Virginia E. Pitzer

## Abstract

**Objective:** Non-disease-specific WHO-CHOICE unit costs are often used in cost and cost-effectiveness studies in the absence of country-specific data. This study aims to compare reported country-specific disease costs and the corresponding WHO-CHOICE estimates. We use generically defined “diarrhea” (including rotavirus diarrhea) and pathogen-specific “respiratory syncytial virus (RSV)” disease as examples.

**Methods:** We updated systematic reviews for both diseases in low-income (LICs), lower-middle-income (LMICs) and upper-middle-income (UMICs) countries. Diarrheal (including a sub-analysis of rotavirus-specific) and RSV-specific outpatient and inpatient costs per episode were extracted and compared with WHO-CHOICE estimates in the same countries. If a consistent pattern of under- or over-estimation was identified, we quantified the magnitude of the discrepancy. All costs were updated to 2022 international dollar values.

**Results:** Out of 1975 new records identified, 23 new cost studies were included. Including previous reviews, we retained 31 diarrhea and 16 RSV studies for comparison. WHO-CHOICE based direct medical costs were similar for diarrheal disease including rotavirus diarrhea, but lower for RSV-related disease. We estimated the cost per episode of diarrhea and RSV in 128 countries. RSV outpatient cost were adjusted by multiplying WHO-CHOICE costs by 6.89 (95% uncertainty interval: 5.58-8.58) in LICs and LMICs and 5.87 (4.95-6.96) in UMICs; RSV inpatient costs were multiplied by 1.43 (1.01-2.01) and 1.36 (0.82-2.27), respectively.

**Conclusion:** WHO-CHOICE based costs should be used cautiously. They aligned well with studies for diarrheal disease, but underestimate costs of RSV-related disease. More country- and disease-specific cost data are needed, especially for RSV in LICs.

## Introduction

Despite remarkable advancements in reducing under-five mortality rates in the past decades, 5 million children died before reaching five years of age worldwide in 2021^1^. Diarrheal diseases and lower respiratory tract infections remain the leading causes of under-five mortality, especially in low- and middle-income countries^1^. Rotavirus and respiratory syncytial virus (RSV) are major causes of diarrheal and lower respiratory tract diseases, respectively. Four rotavirus vaccines are pre-qualified by the World Health Organization (WHO) for global use to prevent rotavirus-related diarrheal disease ^2^. A maternal RSV vaccine and a novel single-dose long-acting monoclonal antibody have been licensed to protect young infants, and other promising candidates are undergoing clinical trials ^3,4^. Given the scarcity of resources and financial constraints in low- and middle-income countries, the implementation of novel prevention strategies for both rotavirus and RSV need to be carefully considered and weighed against other health interventions. The cost and cost-effectiveness of the new immunization programs can inform decision-making at country and global levels. However, there are limited country-specific cost data to inform such analyses in these countries ^5-7^.

The WHO-CHOICE (CHOosing Interventions that are Cost Effective) project collected country costs data between 2008 and 2010 and performed a multivariate regression analysis to estimate the mean non-disease-specific unit costs at the country and regional level with 95% uncertainty intervals (UI) ^8^. The outpatient unit costs present the cost per visit at five facility levels, including health centers without/with beds, and primary, secondary and tertiary hospitals. The inpatient unit costs present cost per hospital bed-day at three hospital facility levels. Although WHO-CHOICE country-specific outpatient and inpatient unit cost estimates are not disease-specific, they have been widely used to assess the budget impact and cost-effectiveness of healthcare programs when local disease-specific cost data are absent. Recently, several studies have measured the actual costs related to diarrheal (including rotavirus diarrhea) and RSV-related disease in different countries. Here we investigate how the WHO-CHOICE estimates compare to the direct medical costs reported in specific studies for these two diseases.

Incorporating uncertainty in cost and cost-effectiveness analysis is a standard component, recommended by health technology agencies and WHO guidelines ^4,9^. However, many cost studies only provide point estimates (i.e. mean and/or median) or dispersion measures from sample data in a specific setting that might not adequately capture the uncertainty. Given the skewness of the cost data, the uncertainties around costs should also be thoroughly evaluated. This study primarily aims to compare the original direct medical cost estimates of diarrhea and RSV-related diseases from the literature with the model-based WHO-CHOICE estimates, stratified based on the World Bank economic classifications of low-income countries (LICs), lower-middle-income countries (LMICs), and upper-middle-income countries (UMICs). In addition, it aims to estimate the country-specific direct medical cost of diarrheal and RSV-related diseases based on published original data with 95% uncertainty ranges (UI).

## Methods

### Systematic review

A systematic review was conducted to identify country-specific medical costs of diarrheal and RSV-related diseases. Two previous systematic reviews, focused on low- and middle- income countries, were identified; Baral and colleagues assessed the cost of illness for childhood diarrhea (up to July 2018) ^10^, and Wittenauer and colleagues investigated the cost of childhood RSV management (up to January 2022) ^7^. We adapted the previously published search strategies and updated the reviews to include recently published studies. The search was conducted on 27^th^ September 2023 with language restriction to English. The inclusion criteria were studies reporting original cost data among children under five years of age in non-high-income countries. The selection was performed by two independent reviewers (XL and CW). Information regarding our search strategies, review process and data extraction can be found in *Appendix section 1*.

### Comparison

From the systematic reviews, we derived country- and disease-specific direct medical, direct non-medical and indirect costs per outpatient and inpatient episode. The hospital length-of-stay (LoS) data were also extracted from studies conducted in the inpatient setting. Meta-analysis was performed to estimate the mean LoS per diarrheal and RSV-related disease and the corresponding 95% UIs. For studies reporting only median LoS, we converted the median to mean using a method for unknown non-normal distributions (R package “estmeansd”) ^11^. Meta-analysis was conducted using the inverse variance method and random effect model in the base case, but a sensitivity analysis was performed with only mean data (without conversion).

The description of WHO-CHOICE estimates is available in Stenberg et al ^8^.We selected the tertiary hospital unit cost for both outpatient cost per visit and inpatient cost per day. To account for the uncertainty around the mean estimates, 1000 samples were drawn from a lognormal distribution based on the mean and 95% UIs of the WHO-CHOICE estimates. The cost per inpatient episode was calculated as cost per day multiplied by the pooled disease-specific LoS, where the LoS was also sampled from a lognormal distribution determined by the pooled mean and 95% UIs.

Costs reported in the literature and the WHO-CHOICE estimates were first converted to the relevant local currency in the year of the cost value, then inflated to 2022 values using the time- and country-specific consumer price index (CPI) from the World Bank database ^12^. Subsequently, we converted the local currency back to international United States dollars (iUSD) using 2022 country-specific purchasing power parities ^13^.

When consistent divergence between WHO-CHOICE estimates and country- and disease-specific costs were observed, considering uncertainty intervals, we conducted a meta-analysis to quantify under or overestimation. Adjustment factors for outpatient and inpatient costs, respectively, were estimated as follows:

Adjustment factor _outpatient_ = outpatient direct medical cost per episode (literature) / WHO-CHOICE estimates per outpatient episode

Adjustment factor _inpatient_ = inpatient direct medical cost per episode (literature) / (WHO-CHOICE estimates per inpatient day* inpatient LoS)

More details are available in *Appendix 2*.*3*.

### Patient and Public Involvement

It was not appropriate or possible to involve patients or the public in the design, or conduct, or reporting, or dissemination plans of our research, because we used previously published aggregated cost data in multiple countries.

## Results

### Summary findings of the systematic review

The literature search yielded 1769 diarrhea and 210 RSV-related disease records. We included 15 studies on diarrheal disease and seven studies on RSV, in addition to the existing studies identified in the previous systematic reviews ^7,10^. A total of 31 diarrhea (including 17 rotavirus-specific diarrhea studies) and 16 RSV studies were included in the comparison (Figure 1 and *Appendix section 2*.*1*).

**Figure 1:**
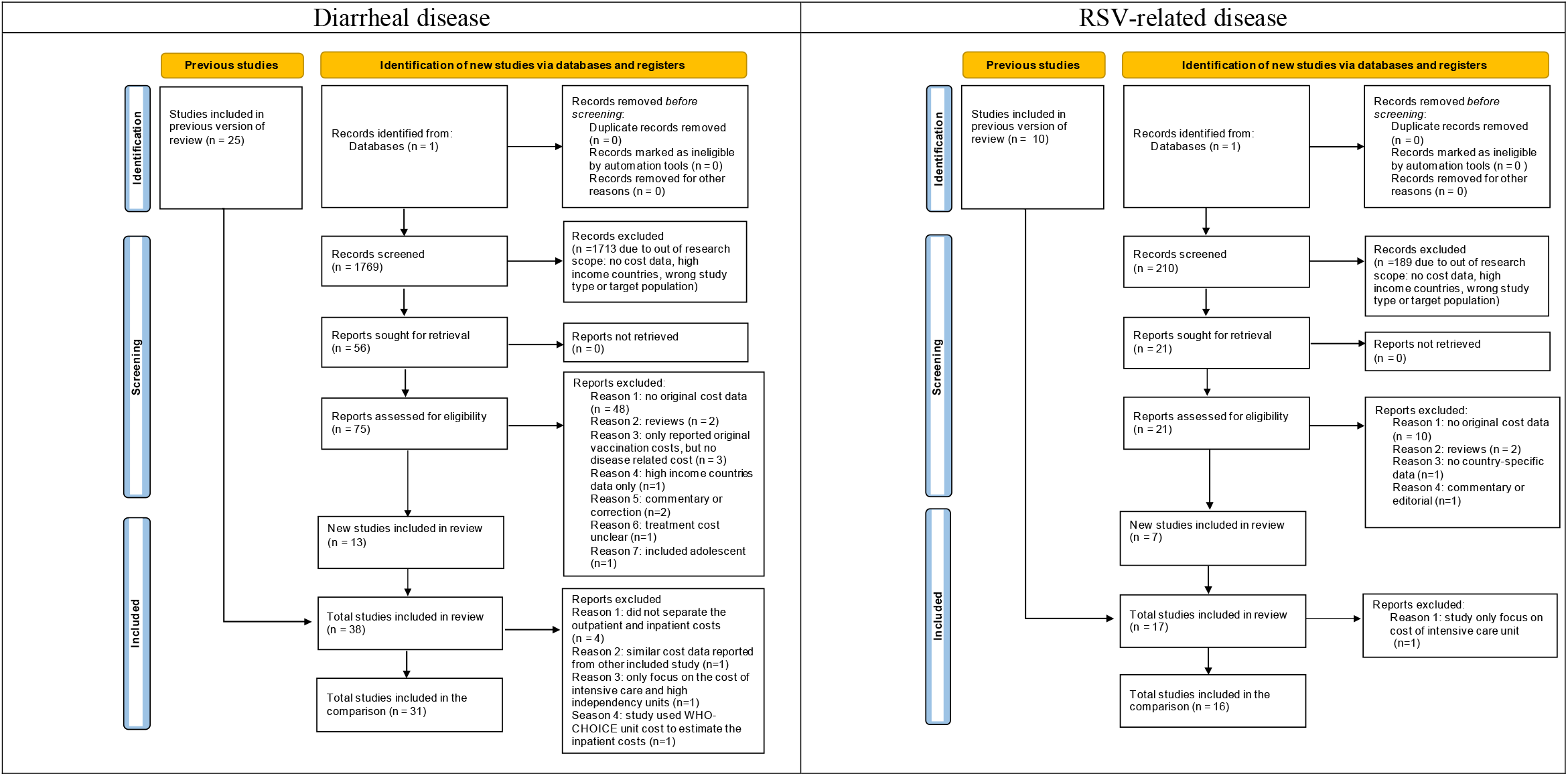
PRISMA flow diagram

There were more studies reporting only inpatient costs than both inpatient and outpatient costs, and the majority of studies were conducted in tertiary hospitals (*Appendix section 2*.*1*). Figure 2 demonstrates the geographic representation of the included studies. Fewer diarrheal studies (n=7) but most RSV studies (n=13) were conducted in UMICs. Limited cost studies exist in LICs, especially for RSV-related disease (n=1). India had the highest number of diarrheal studies (N=6, of which, 2 rotavirus-diarrhea studies), whereas China had the highest number of RSV studies (n=4).

**Figure 2:**
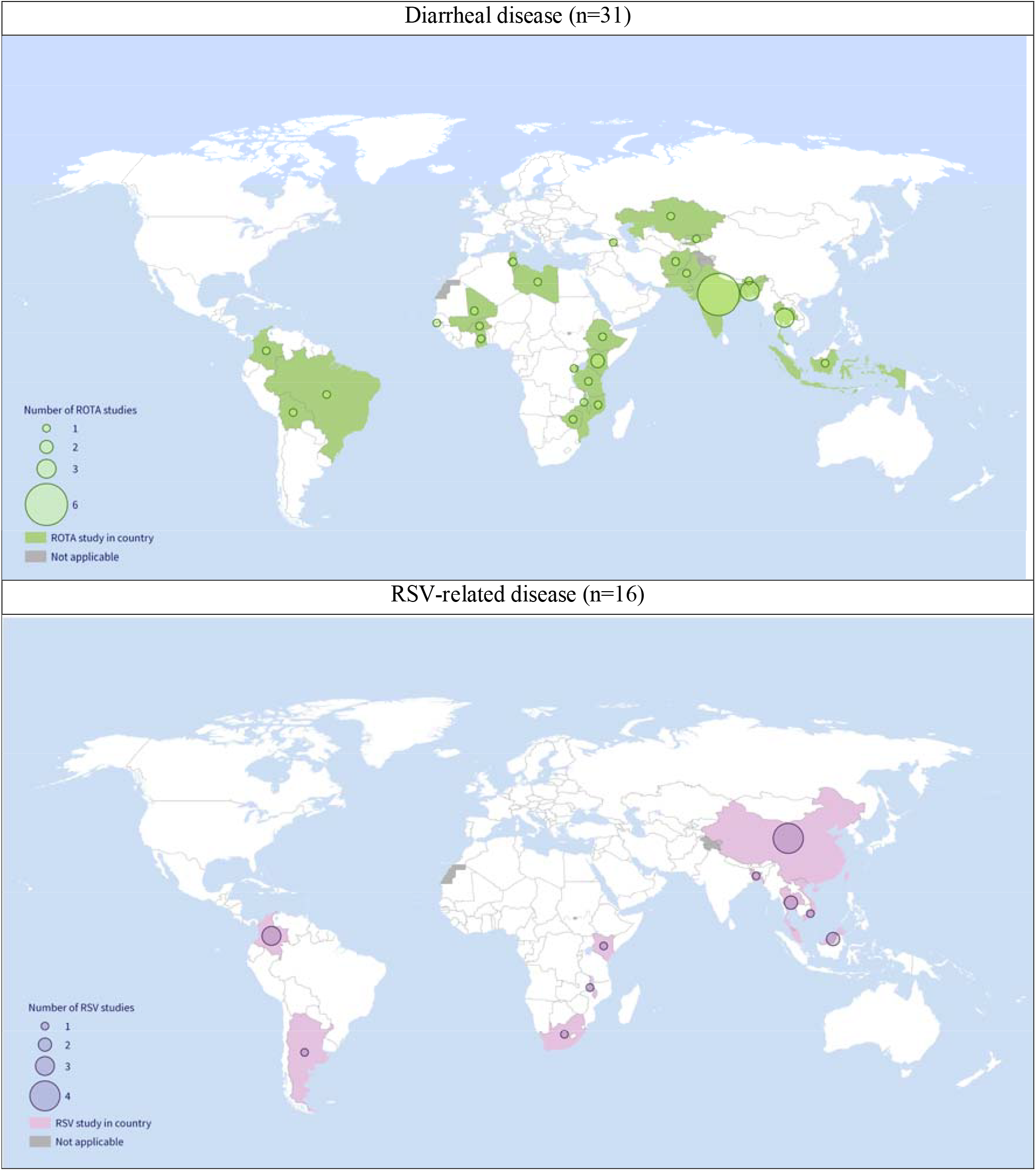
Map of the number of studies for diarrhea (top) and RSV-related diseases (bottom) in low- and middle-income countries. White space: no data or high-income countries. Note: one study included 7 countries

The fine age stratification is important for both diarrhea and RSV empirical studies, because the most severe cases often occur at infancy ^14^. All diarrheal (including rotavirus-diarrheal) studies grouped children under 5 years of age, whereas seven RSV studies reported the direct medical cost stratified by more refined age groups (*Appendix Table 2*.*1 and 2*.*2*). All RSV studies reported laboratory confirmation due to their nature, but confirmatory tests were not conducted in one third of the diarrheal studies (n=12). Among the studies that compared all-cause diarrhea and rotavirus-positive diarrhea, no significant cost difference was found.

#### Pooled hospital length-of-stay for diarrheal and RSV-related diseases

The meta-analysis estimated a longer LoS for RSV-related disease than for diarrheal disease, especially in UMICs (Table 1). Sensitivity analysis excluding median LoS for diarrheal disease shows similar results with overlapping UIs. The data extraction, meta-analyses, and sensitivity analyses are presented in *Appendix section 2*.*2*.

**Table 1:** Pooled estimates of hospital length-of-stay (days) in low- and lower-middle-income countries (LICs and LMICs) and upper-middle-income countries (UMICs).

|  | Low- and lower-middle-income countries |  | Upper-middle-income countries |  |
| --- | --- | --- | --- | --- |
| Disease | Number of studies | Mean (95% uncertainty interval) (days) | Number of studies | Mean (95% uncertainty interval) (days) |
| Diarrhea | N=14 | 4.27 (3.69-4.85) | N=4 | 4.90 (3.86-4.40) |
| RSV | N=2 | 5.17 (1.43-8.91) | N=9 | 6.01 (4.82-7.21) |
| Subgroup |  |  |  |  |
| Rotavirus-specific diarrhea | N=5 | 4.71 (3.41-6.00) | N=2 | 5.70 (2.57-8.82) |

### Comparing the literature with the WHO-CHOICE estimates

We show inpatient and outpatient direct medical costs comparison between WHO-CHOICE estimates and mean costs reported in the literature stratified by income groups for diarrheal (Figure 3) and RSV-related diseases (Figure 4).

**Figure 3:**
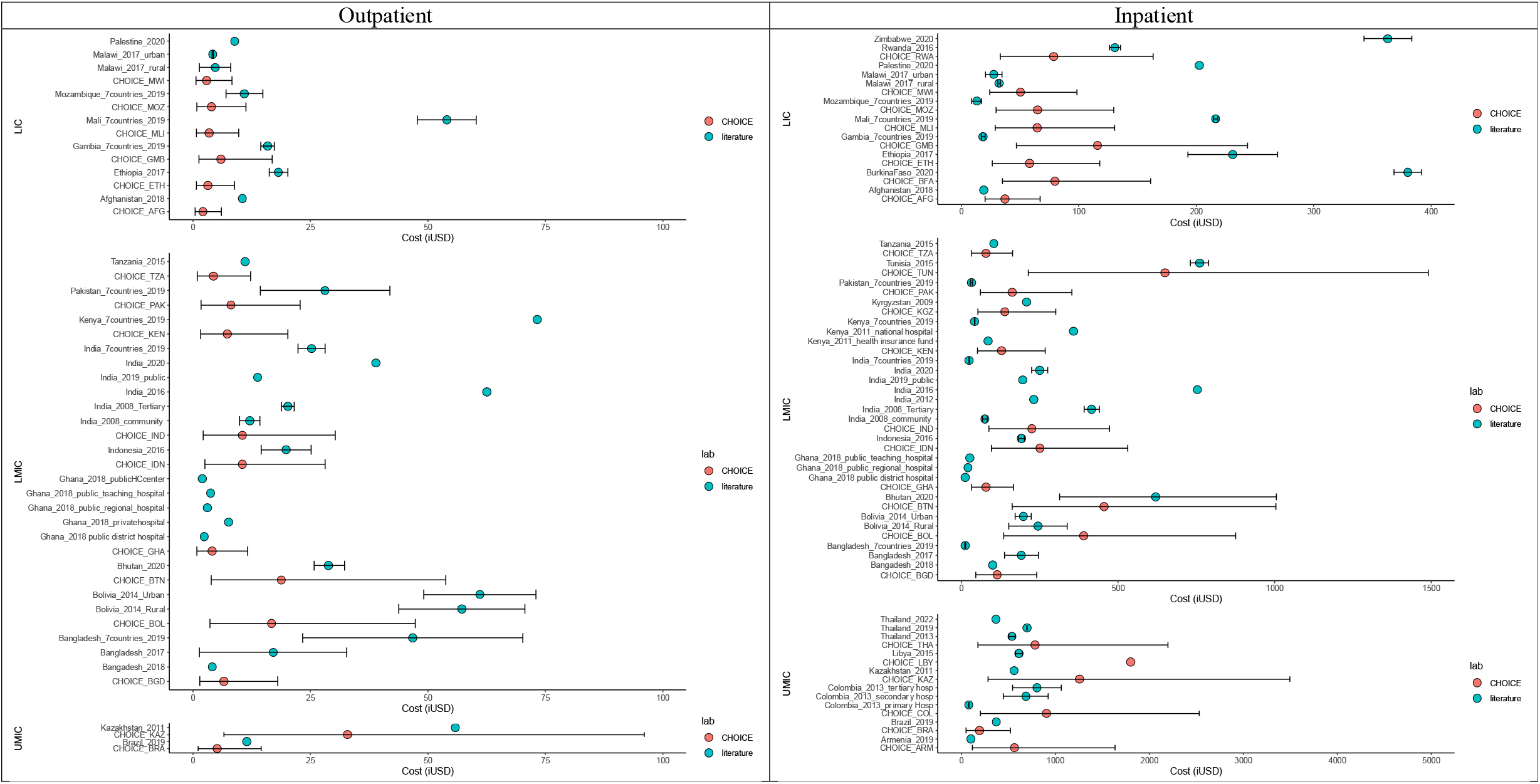
Outpatient (left) and inpatient (right) direct medical costs for diarrheal disease. Red dots represent the WHO-CHOICE estimates, while blue dots represent mean costs reported in the literature (iUSD in 2022 value). Black bars represent the 95% ertainty intervals. Note: WHO-CHOICE estimates were not available for Zimbabwe and Palestine.

**Figure 4:**
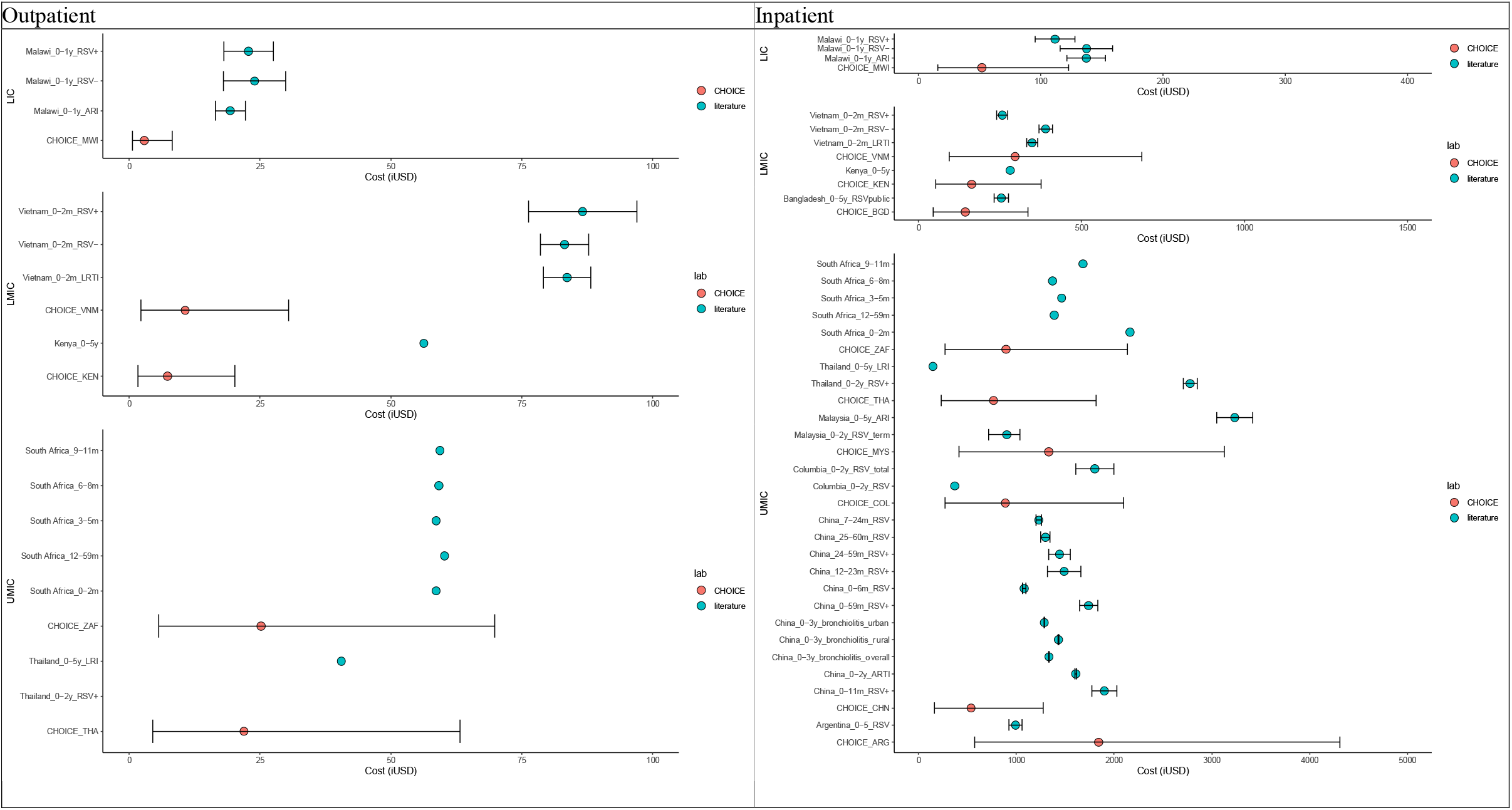
Outpatient (left) and inpatient (right) direct medical costs for RSV disease. Red dots represent the WHO-CHOICE estimates, while blue dots represent mean costs reported in the literature (iUSD in 2022 value). Black bars represent the 95% ertainty intervals.

#### Diarrheal disease

Overall, the diarrheal disease-specific direct medical costs reported in the literature fall within the 95% UIs of WHO-CHOICE outpatient and inpatient estimates, with the exception of Mali and Ethiopia (Figure 3). The direct medical costs for Mali were reported by a multi-country study conducted in seven LICs and LMICs ^15^. In this study, both outpatient and inpatient costs in Mali were higher than the corresponding WHO-CHOICE estimates, but also much higher than the other six countries ^15^. In the same study, the outpatient cost in Kenya was also higher than the WHO-CHOICE outpatient cost (without overlapping 95% UIs); however, the inpatient cost was lower than the WHO-CHOICE inpatient cost per episode (with overlapping 95% UIs) ^15^. Moreover, an Ethiopian study had higher inpatient and outpatient costs than WHO-CHOICE estimates, possibly due to the inclusion of private clinic/hospitals, which accounted for 34% of inpatient and 7% of outpatient cases ^16^. A study conducted in Burkina Faso also estimated a higher inpatient cost compared to WHO-CHOICE estimates. A subgroup analysis focusing on rotavirus-specific diarrhea was also performed, and the findings are consistent with the general diarrheal disease findings (*Appendix Figure 2*.*3*).

For countries with multiple studies, large variations in both outpatient and inpatient costs were observed. For example, six studies from India reported notably divergent costs in both inpatient and outpatient settings. The uncertainty in WHO-CHOICE estimates of outpatient and inpatient costs in India captures some, but not all, of the variability in observed costs.

#### RSV-related disease

The reported average outpatient cost per episode for RSV disease in the literature appeared to be much higher than the corresponding WHO-CHOICE estimates, especially in LICs and LMICs (without overlapping in 95% UIs). A similar pattern was observed for the inpatient costs per episode; however, the 95% UIs of most studies overlapped with the WHO-CHOICE estimates. There was also a within-country variation of inpatient costs observed across four Chinese studies, but the overall variation was relatively modest, likely due to the large sample sizes (range: 261 - 42,928 cases).

Since the observed RSV disease costs and their uncertainty ranges were consistently higher than the WHO-CHOICE estimates, we performed a meta-analysis of the observed costs to estimate adjustment factors that can be applied to the WHO-CHOICE estimates (*Appendix section 2*.*3*). The estimated adjustment factor for outpatient costs was 6.89 (95% UIs: 5.58-8.58) in LICs and LMICs and 5.87 (4.95-6.96) in UMICs. For inpatient costs, they were 1.43 (1.01-2.01) and 1.36 (0.82-2.27), respectively.

### Direct medical costs per country per disease

Based on our comparison, we estimated the mean and 95% UIs of the direct medical costs for diarrheal and RSV disease in both outpatient and inpatient settings across 128 countries (*Appendix section 2*.*5*). For the countries where data were available, we present both WHO-CHOICE estimates and the country- and disease-specific costs. We pooled the costs data for countries with multiple studies available (e.g., India, China). *Appendix Table 2*.*6* reports country-specific direct medical costs for diarrheal disease per outpatient and inpatient episode. For RSV disease, *Appendix Tables 2*.*7* and *2*.*8* report the country-specific direct medical costs with and without adjustment factors, respectively.

## Discussion

We conducted reviews of cost and hospital length-of-stay studies, including meta-analyses to estimate costs per inpatient and outpatient disease episode due to diarrheal and RSV-related diseases in LICs and LMICs and UMICs. Our review shows that there are still limited disease-specific original cost studies in these non-high-income countries, especially for RSV-related disease and for LICs. Moreover, large within-country variations were observed for diarrheal disease in India, likely due to the study location (i.e. cities vs rural areas), years (i.e. 2008 vs 2018) and the methodology (i.e. micro-costing vs expert panel). Modest variation was also observed for RSV-related disease in China.

Our comparison illustrates that WHO-CHOICE estimates provide good approximations for the direct medical costs related to diarrheal disease (including rotavirus diarrhea), but might underestimate the direct medical costs of RSV-related disease, especially for outpatient costs. The WHO-CHOICE estimates provide a useful resource for representing uncertainty in cost estimates and informing economic evaluations of immunization programs in the absence of country-specific cost data, but adjustment factors seem warranted for certain diseases.

There are several potential reasons why the WHO-CHOICE estimates may have consistently been lower than the observed direct medical costs for RSV-related disease. First, WHO-CHOICE estimates did not include the cost of medication, diagnosis and medical procedures. For diarrhea, the medication costs were mainly for oral rehydration solution or intravenous therapy, which accounted for a relatively small proportion of the total outpatient and inpatient direct medical costs ^10^. The costs of laboratory test were not explicitly reported in most diarrhea studies. In contrast, the direct medical costs for RSV-related disease often included costs of laboratory tests, procedures (i.e. oxygen delivery) and imaging (i.e. chest X-ray), in addition to consultation and medication costs. These costs represent a relatively large portion of the overall costs, especially for outpatient cases in tertiary hospitals. For example, Do and colleagues reported the medication and diagnostic costs of RSV disease were 5- and 2-fold higher, respectively, than the consultation fees in the outpatient setting. Moreover, there might be a laboratory test bias that serious cases are more likely to be tested. Second, the pooled LoS was used to calculate the WHO-CHOICE cost per inpatient episode. For both diarrheal and RSV studies, more studies reported median than mean LoS, likely due to the skewness of the data. For our analysis, the pooled LoS estimates were based on reported means (and medians converted to means). For diarrheal disease, sensitivity analysis showed that the LoS using mean estimates only (n=5) was slightly longer than that based on both means and converted medians, but the 95% UIs overlapped. For RSV studies, only one study from a UMIC reported mean LoS (together with sample size and SD). Thus, the pooled mean LoS for RSV disease might be underestimated, resulting in a potential underestimation of the WHO-CHOICE inpatient cost per episode for RSV disease. Third, there are much fewer studies investigating the RSV-related costs, especially for outpatient care. Majority of the RSV outpatient and inpatient costs were collected from tertiary hospitals, hence the average RSV-related cost per episode might be overestimated or not representative. More RSV studies are needed to further enhance the comparison. Fourth, there might be publication bias. Most RSV studies were published between 2020 and 2023, likely due to the advancement in developing RSV interventions. The diarrheal studies were published throughout last decade, and several studies were published after the implementation of rotavirus vaccines. Last, WHO-CHOICE estimates were non-age-specific. Unlike most of the diarrhea studies that included children under 5 years of age, several RSV studies only included very young infants (i.e., less than 3 months or 1 year); this might also explain the higher costs of RSV-related disease.

To the best of our knowledge, this is the first formal comparison between disease-specific costs and WHO-CHOICE estimates, using a general disease of diarrhea and pathogen-specific RSV-related disease as examples. Based on our review and comparison, we reported the direct medical costs with 95% UIs for outpatients and inpatients with diarrheal disease across 128 countries. For RSV, we also estimated these costs with and without adjustment factors that accounted for potential underestimation of the disease-specific costs. In alignment with the recommendation of the WHO-CHOICE project and WHO guidelines ^8,9^, country- and disease-specific original cost data should be used where possible, while properly considering how best to represent uncertainty when extrapolating from specific settings to the entire country. In cases where such studies are unavailable, considering WHO-CHOICE estimates seems justified, especially when accounting for the associated uncertainty intervals. Additionally, it may be prudent to consider the potential underestimation of disease-specific costs by using the adjustment factors.

Furthermore, due to either insufficient data or oversimplified assumptions, many cost and cost-effectiveness studies often use an arbitrary range (e.g. ± 20%) of the mean values to evaluate the uncertainty around parameters, including cost parameters in deterministic one-way or probabilistic sensitivity analyses. Given the skewness of the cost data, the uncertainties around costs should be thoroughly evaluated in order to appropriately conduct an economic evaluation and correctly identify the influential factors ^4,9^. Our study has estimated uncertainty around the direct medical costs, which will facilitate the further economic evaluations.

There are several limitations to our study. First, we used the tertiary hospital unit cost from WHO-CHOICE in order to make a fair comparison, mainly because most of the cost studies were conducted in tertiary hospitals. However, cases in tertiary hospitals might be more severe, especially for outpatient episodes, and this could result in an overestimation of disease-specific costs in countries where country-specific data are unavailable. Secondly, using adjustment factors for the outpatient and inpatient costs of RSV-related disease might be a moderately simplified approach. However, in the absence of country- and disease-specific data, applying adjustment factors to WHO-CHOICE estimates might offer more reliable direct medical costs. Thirdly, we did not explicitly assess direct nonmedical and indirect costs, although we extracted the data throughout our systematic review. We can only compare the direct medical costs with the WHO-CHOICE estimates. Notably, Baral and colleagues proposed an approach to estimate the diarrheal disease related direct nonmedical costs as a proportion of direct medical costs, and indirect costs as country-specific gross domestic product per day multiplied by the duration of illness ^10^. A similar approach might be considered for RSV-related disease. Next, the proportion of direct medical costs paid by patients versus the health care system/insurance (if any) is not often explicitly reported in the studies included in our literature review. Such costs from the patients’ perspective can be a large proportion of their family income and might lead to poverty. These aspects are important to address in future studies. Moreover, for countries with multiple costs studies, we illustrated the variability in costs in Figures 3 and 4, but pooled the data to estimate country-specific costs; this may underestimate the uncertainty in costs for countries with large variations, such as India. Although immunization programs are likely decided on at a national level, these should be informed by regional/area-specific data (including cost data), especially if variation is expected to be large. Typically, the proportion of private health care provision (and higher out-of-pocket expenditures) is associated with degree of urbanization. Lastly, the general CPI was used to inflate the medical costs, because health-sector specific CPIs are not available in most countries analyzed.

## Conclusion

More country- and disease-specific cost data are needed, especially for RSV-related disease in LICs. WHO-CHOICE estimates provide valuable inputs for cost-of-illness, cost-effectiveness, budget-impact and global economic burden of disease studies. As demonstrated by our study, WHO-CHOICE outpatient and inpatient costs are comparable with empirical country-specific cost of diarrheal disease, but they might underestimate the cost of RSV-related disease in children. Hence, a comparison is essential before WHO-CHOICE estimates are directly applied to other infectious diseases. Moreover, the appropriate uncertainty ranges should be accounted for in economic evaluations

## Data Availability

All data produced in the present work are contained in the manuscript and supplement

## Ethics approval

Ethics approval is unnecessary because we used publicly available data.

## Author Contributions

VEP initiated the study, and analyses were conceptualized by XL, JB, EOA, PB and VEP. XL performed systematic literature searches, abstract/full text screening and data extraction. CW conducted independent abstract/full text screening and reviewed the data extraction tables. XL analyzed the data under the supervision of VEP. LB, EOA, JK, JB and PB advised on the methodology, interpretation of the results, and the strengths and limitations of the study. XL, PB and VEP wrote the initial manuscript draft. All authors critically reviewed the manuscript and provided final approval of the manuscript.

## Competing interests

VEP was previously a member of the WHO Immunization and Vaccine-related Implementation Research Advisory Committee (IVIR-AC). Outside the submitted work, PB and LB reports grants from RESCEU and PROMISE, Innovative Medicines Initiative 2 of the European Commission, but they have not received any personal fees or other personal benefits. LB is the founding chairman of the ReSViNET Foundation. He also declares that he has regular interaction with pharmaceutical and other industrial partners. He has not received personal fees or other personal benefits. Outside the submitted work, his institute UMCU has received major funding (>€100,000 per industrial partner) for investigator-initiated studies from AstraZeneca, Sanofi, Janssen, Pfizer, MSD and MeMed Diagnostics. UMCU has received major funding from the Bill and Melinda Gates Foundation. UMCU has received major funding by Julius Clinical for participating in clinical studies sponsored by AstraZeneca, Merck and Pfizer. UMCU received minor funding (€1,000-25,000 per industrial partner) for consultation, DSMB membership or invited lectures by Ablynx, Bavaria Nordic, GSK, Novavax, Pfizer, Moderna, AstraZeneca, MSD, Sanofi, Janssen. Other authors declared no competing interests.

## Funding

This research was supported by a grant from National Institutes of Health/National Institute of Allergy and Infectious Diseases (R01AI112970 and R01AI137093 to VEP).

