## Appendix for "Cost per episode of diarrhea and respiratory syncytial virus (RSV) in 128 low- and middle-income countries: how well do disease-specific and WHO-CHOICE estimates align?"

### 1. Methods

#### 1.1 Search terms and inclusion and exclusion criteria

A systematic review was conducted to identify country-specific medical costs of diarrheal and respiratory syncytial virus (RSV)-related diseases. The search terms were modified in accordance with the published systematic reviews ^1 2^. The search was conducted via PubMed on 27^th^ September 2023.

Table 1.1: Search terms for cost of childhood diarrheal disease via PubMed

|  | keywords | Number of hits |
| --- | --- | --- |
| #1 | rotavirus OR gastroenteritis OR diarrhea* OR diarrhea OR diarrhoea* OR diarrhoea | 394,467 |
| #2 | Cost OR economic OR “cost of illness” OR “burden of illness” OR “illness burden” OR “economic burden” OR expenditure OR funding | 1,912,593 |
| #3 | infant* OR child OR children | 3,702,986 |
| #4 | #1 AND #2 AND #3 | 6,341 |
| Other limits | 2018/06/01 – 2023/09/27, humans, English language | 1769 |

Table 1.2: Search terms for cost of childhood RSV-related disease via PubMed

|  | keywords | Number of hits |
| --- | --- | --- |
| #1 | "human respiratory syncytial virus" OR "respiratory syncytial virus" OR "respiratory syncytial virus infection" OR "respiratory syncytial virus vaccine" OR "rsv" | 23,001 |
| #2 | Cost OR economic OR “cost of illness” OR “burden of illness” OR “illness burden” OR “economic burden” OR expenditure OR funding | 1,912,593 |
| #3 | infant* OR child OR children | 3,702,986 |
| #4 | #1 AND #2 AND #3 | 1,254 |
| Other Limits | 2022/01/01 – 2023/09/27, humans, English language | 210 |

Table 1.3: Inclusion and exclusion criteria

| **PICOS** | **Inclusion** | **Exclusion** |
| --- | --- | --- |
| Population | Children ≤5 years of age | Older children or adults |
| Outcome | Study reporting original cost (including direct and indirect cost); cost must be in numerical values (in any currency) | Studies (including reviews) that refer to cost data from previous studies (need to retrieve the original studies) |
| Study type | - Cost of illness study - Burden of disease study reporting original cost data - Economic evaluation (including cost-effectiveness, cost-utility or cost-benefit study) reporting original cost data | - Reviews related to cost studies (used for reference tracing) - Economic evaluations that refer to other studies for cost data (used for reference tracing) - Conference abstract, comment or letter |
| Country | Low-income, lower-middle-income and upper-middle-income | High-income |
| Language | English | Other languages |

#### 1.2 Review process

The primary reviewer (XL) conducted the literature search, screened and selected the records based on the titles and abstracts according to the inclusion and exclusion criteria. A second independent reviewer (CW) screened 10% of randomly selected titles and abstracts. The two reviewers had a 95% consistency rate, and any disagreements were discussed and resolved. Next, the full texts of articles were assessed by the primary reviewer, and 20% of the articles were again screened by the second reviewer independently. There was only one disagreement (95% consistency rate), and it was discussed and resolved by the two reviewers.

#### 1.3 Data extraction

A data extraction table for each disease was predefined including study characteristics (i.e., study type, period, perspective), direct medical cost, direct non-medical cost, indirect cost and total cost per outpatient and inpatient episode. The primary reviewer (XL) extracted the data, and 20% of the included studies were checked by the second reviewer.

#### 1.4 WHO-CHOICE estimates

The mean of WHO-CHOICE estimates was reported in both international US dollars (iUSD) and in national currency unit in 2010 value, However, the 95% UIs were only reported in iUSD ^3^.

To update the WHO-CHOICE estimates, the iUSD values were first converted to local currency in 2010, then inflated to their 2022 value using the country-specific consumer price index from the World Bank database ^4^. Finally, we converted local currency back to iUSD using 2022 purchasing power parities ^5^. Table 1.4 contains the WHO-CHOICE estimates with uncertainty intervals (UIs) in iUSD 2022 values.

Table 1.4: WHO-CHOICE outpatient and inpatient cost estimates per bed-day in tertiary hospitals (international dollars updated to 2022 value)

| Country | ISO3 | Income Group | Outpatient | | | Inpatient | | |
| --- | --- | --- | --- | --- | --- | --- | --- | --- |
|  |  |  | Mean | Lower 95% uncertainty interval | Upper 95% uncertainty interval | Mean | Lower 95% uncertainty interval | Upper 95% uncertainty interval |
| Afghanistan | AFG | LIC | 2.11 | 0.43 | 6.08 | 5.28 | 2.14 | 10.59 |
| Angola | AGO | LMIC | 16.22 | 3.42 | 46.18 | 90.63 | 34.32 | 189.98 |
| Albania | ALB | UMIC | 25.17 | 5.13 | 72.81 | 154.79 | 58.92 | 323.46 |
| Argentina | ARG | UMIC | 46.03 | 10.99 | 123.73 | 341.41 | 138.46 | 686.41 |
| Armenia | ARM | UMIC | 18.46 | 4.12 | 51.27 | 100.66 | 37.08 | 214.49 |
| Azerbaijan | AZE | UMIC | 16.48 | 3.31 | 48.02 | 108.99 | 44.35 | 218.69 |
| Burundi | BDI | LIC | 2.60 | 0.52 | 7.56 | 6.49 | 2.45 | 13.63 |
| Benin | BEN | LMIC | 4.71 | 1.06 | 13.04 | 17.11 | 7.19 | 33.66 |
| Burkina Faso | BFA | LIC | 4.66 | 0.93 | 13.63 | 14.96 | 6.08 | 30.04 |
| Bangladesh | BGD | LMIC | 6.57 | 1.50 | 18.07 | 22.90 | 9.12 | 46.58 |
| Bulgaria | BGR | UMIC | 32.45 | 6.33 | 95.73 | 234.32 | 91.87 | 480.97 |
| Bosnia & Herzegovina | BIH | UMIC | 22.06 | 4.82 | 61.79 | 135.46 | 53.65 | 276.36 |
| Belarus | BLR | UMIC | 23.03 | 4.89 | 65.41 | 165.93 | 68.32 | 330.48 |
| Belize | BLZ | UMIC | 15.49 | 3.14 | 44.88 | 91.13 | 34.62 | 190.67 |
| Bolivia | BOL | LMIC | 16.72 | 3.58 | 47.32 | 85.73 | 33.00 | 177.94 |
| Brazil | BRA | UMIC | 5.13 | 1.11 | 14.49 | 32.92 | 13.33 | 66.24 |
| Bhutan | BTN | LMIC | 18.79 | 3.92 | 53.80 | 100.60 | 40.14 | 204.27 |
| Botswana | BWA | UMIC | 31.28 | 7.13 | 85.95 | 236.45 | 88.68 | 498.50 |
| Central African Republic | CAF | LIC | 3.46 | 0.70 | 10.02 | 10.20 | 3.99 | 20.99 |
| China | CHN | UMIC | 16.95 | 3.71 | 47.49 | 99.64 | 39.58 | 202.92 |
| Côte d'Ivoire | CIV | LMIC | 6.03 | 1.34 | 16.74 | 22.61 | 8.74 | 46.79 |
| Cameroon | CMR | LMIC | 7.38 | 1.55 | 21.07 | 29.71 | 11.72 | 60.77 |
| Congo - Kinshasa | COD | LIC | 0.78 | 0.16 | 2.24 | 1.73 | 0.66 | 3.62 |
| Congo - Brazzaville | COG | LMIC | 8.58 | 1.94 | 23.68 | 44.14 | 17.89 | 88.77 |
| Colombia | COL | UMIC | 46.37 | 10.17 | 129.76 | 164.45 | 65.52 | 334.26 |
| Comoros | COM | LMIC | 4.39 | 0.76 | 13.58 | 13.43 | 5.06 | 28.26 |
| Cape Verde | CPV | LMIC | 10.99 | 2.47 | 30.41 | 54.27 | 21.91 | 109.42 |
| Costa Rica | CRI | UMIC | 31.11 | 6.88 | 86.71 | 214.12 | 82.89 | 442.91 |
| Djibouti | DJI | LMIC | 7.32 | 1.49 | 21.18 | 30.06 | 12.09 | 60.73 |
| Dominica | DMA | UMIC | 28.53 | 4.33 | 93.40 | 194.74 | 77.51 | 396.09 |
| Dominican Republic | DOM | UMIC | 22.62 | 4.60 | 65.49 | 139.86 | 52.59 | 294.42 |
| Algeria | DZA | LMIC | 16.58 | 3.58 | 46.70 | 103.66 | 37.79 | 222.28 |
| Ecuador | ECU | UMIC | 22.82 | 5.04 | 63.66 | 146.53 | 56.48 | 303.90 |
| Egypt | EGY | LMIC | 16.27 | 3.20 | 47.82 | 87.57 | 34.59 | 178.93 |
| Eritrea | ERI | LIC | 1.99 | 0.45 | 5.51 | 4.92 | 1.79 | 10.57 |
| Ethiopia | ETH | LIC | 3.17 | 0.70 | 8.85 | 10.04 | 3.78 | 21.10 |
| Fiji | FJI | UMIC | 14.40 | 3.05 | 40.98 | 76.93 | 28.48 | 163.44 |
| Micronesia (Federated States of) | FSM | LMIC | 9.43 | 1.96 | 27.03 | 42.90 | 17.47 | 86.04 |
| Gabon | GAB | UMIC | 31.17 | 7.11 | 85.65 | 233.36 | 92.34 | 476.32 |
| Georgia | GEO | UMIC | 15.16 | 3.01 | 44.37 | 81.47 | 31.89 | 167.40 |
| Ghana | GHA | LMIC | 4.08 | 0.87 | 11.60 | 14.74 | 5.48 | 31.24 |
| Guinea | GIN | LIC | 5.60 | 1.22 | 15.70 | 16.86 | 6.82 | 33.96 |
| Gambia | GMB | LIC | 5.93 | 1.25 | 16.90 | 23.22 | 9.29 | 47.08 |
| Guinea-Bissau | GNB | LIC | 3.70 | 0.78 | 10.55 | 12.12 | 4.92 | 24.33 |
| Equatorial Guinea | GNQ | UMIC | 70.27 | 14.15 | 204.41 | 634.66 | 240.22 | 1330.72 |
| Grenada | GRD | UMIC | 25.40 | 5.86 | 69.43 | 169.25 | 67.87 | 342.64 |
| Guatemala | GTM | UMIC | 15.85 | 3.35 | 45.11 | 78.76 | 31.53 | 159.62 |
| Guyana | GUY | UMIC | 8.53 | 1.81 | 24.23 | 38.86 | 14.69 | 81.54 |
| Honduras | HND | LMIC | 13.03 | 2.73 | 37.25 | 63.33 | 25.09 | 129.18 |
| Haiti | HTI | LMIC | 3.89 | 0.75 | 11.55 | 12.54 | 4.92 | 25.74 |
| Indonesia | IDN | LMIC | 10.49 | 2.52 | 28.11 | 53.99 | 22.59 | 106.46 |
| India | IND | LMIC | 10.52 | 2.18 | 30.24 | 48.14 | 20.24 | 94.63 |
| Iran | IRN | LMIC | 18.75 | 3.68 | 55.17 | 133.81 | 51.47 | 277.86 |
| Iraq | IRQ | UMIC | 6.09 | 1.34 | 17.01 | 29.47 | 11.31 | 61.30 |
| Jamaica | JAM | UMIC | 20.02 | 3.92 | 58.93 | 122.72 | 47.84 | 252.77 |
| Jordan | JOR | UMIC | 16.82 | 3.80 | 46.43 | 91.89 | 36.58 | 186.86 |
| Kazakhstan | KAZ | UMIC | 32.89 | 6.56 | 96.08 | 230.07 | 93.33 | 462.48 |
| Kenya | KEN | LMIC | 7.32 | 1.65 | 20.19 | 26.12 | 10.70 | 52.19 |
| Kyrgyzstan | KGZ | LMIC | 6.89 | 1.55 | 19.07 | 28.38 | 10.86 | 59.10 |
| Cambodia | KHM | LMIC | 7.53 | 1.52 | 21.87 | 30.84 | 11.96 | 63.72 |
| Kiribati | KIR | LMIC | 7.76 | 1.56 | 22.63 | 30.52 | 12.13 | 62.11 |
| Laos | LAO | LMIC | 8.48 | 1.89 | 23.56 | 36.08 | 14.32 | 73.48 |
| Lebanon | LBN | LMIC | 36.81 | 7.90 | 104.00 | 266.31 | 102.98 | 551.26 |
| Liberia | LBR | LIC | 4.01 | 0.86 | 11.30 | 10.48 | 3.82 | 22.48 |
| Libya | LBY | UMIC | 43.30 | 9.97 | 118.44 | 330.93 | 134.55 | 664.27 |
| St. Lucia | LCA | UMIC | 28.06 | 6.19 | 78.34 | 187.73 | 76.34 | 376.79 |
| Sri Lanka | LKA | LMIC | 14.34 | 3.36 | 38.91 | 78.79 | 30.69 | 162.36 |
| Lesotho | LSO | LMIC | 4.76 | 1.02 | 13.43 | 17.94 | 6.84 | 37.46 |
| Morocco | MAR | LMIC | 12.79 | 2.73 | 36.26 | 66.05 | 26.66 | 133.19 |
| Moldova | MDA | UMIC | 11.85 | 2.41 | 34.34 | 53.11 | 21.71 | 106.25 |
| Madagascar | MDG | LIC | 3.14 | 0.69 | 8.79 | 9.28 | 3.54 | 19.34 |
| Maldives | MDV | UMIC | 22.40 | 4.52 | 65.12 | 136.10 | 55.65 | 272.24 |
| Mexico | MEX | UMIC | 36.78 | 8.23 | 101.98 | 282.01 | 111.81 | 574.92 |
| Marshall Islands | MHL | UMIC | 7.85 | 1.68 | 22.20 | 33.55 | 13.11 | 69.00 |
| North Macedonia | MKD | UMIC | 29.01 | 6.08 | 82.87 | 193.45 | 76.59 | 394.74 |
| Mali | MLI | LIC | 3.46 | 0.75 | 9.73 | 11.53 | 4.51 | 23.71 |
| Myanmar (Burma) | MMR | LMIC | 3.08 | 0.68 | 8.63 | 10.32 | 3.99 | 21.36 |
| Montenegro | MNE | UMIC | 36.17 | 7.31 | 105.07 | 264.90 | 101.46 | 551.51 |
| Mongolia | MNG | LMIC | 10.14 | 2.14 | 28.87 | 49.95 | 19.61 | 102.45 |
| Mozambique | MOZ | LIC | 3.93 | 0.81 | 11.29 | 11.60 | 4.64 | 23.54 |
| Mauritania | MRT | LMIC | 6.74 | 1.51 | 18.69 | 29.03 | 11.43 | 59.43 |
| Mauritius | MUS | UMIC | 40.60 | 9.30 | 111.30 | 294.67 | 115.84 | 603.87 |
| Malawi | MWI | LIC | 2.89 | 0.61 | 8.25 | 8.28 | 3.22 | 17.08 |
| Malaysia | MYS | UMIC | 32.79 | 6.96 | 93.12 | 246.82 | 99.52 | 497.99 |
| Namibia | NAM | UMIC | 16.41 | 3.40 | 47.16 | 94.50 | 37.33 | 193.09 |
| Niger | NER | LIC | 2.30 | 0.47 | 6.67 | 5.72 | 2.24 | 11.74 |
| Nigeria | NGA | LMIC | 11.05 | 2.32 | 31.51 | 46.68 | 17.94 | 97.00 |
| Nicaragua | NIC | LMIC | 10.94 | 2.48 | 30.15 | 53.02 | 20.91 | 108.44 |
| Nepal | NPL | LMIC | 4.30 | 0.95 | 12.02 | 14.89 | 5.72 | 30.95 |
| Pakistan | PAK | LMIC | 8.08 | 1.73 | 22.85 | 33.83 | 13.15 | 69.81 |
| Peru | PER | UMIC | 22.46 | 5.45 | 59.88 | 146.47 | 55.44 | 307.11 |
| Philippines | PHL | LMIC | 12.22 | 2.70 | 34.09 | 60.16 | 23.16 | 124.88 |
| Palau | PLW | UMIC | 43.39 | 9.64 | 120.72 | 343.19 | 130.79 | 716.66 |
| Papua New Guinea | PNG | LMIC | 8.12 | 1.64 | 23.62 | 31.70 | 12.18 | 65.89 |
| Paraguay | PRY | UMIC | 16.41 | 3.55 | 46.20 | 90.80 | 35.80 | 185.75 |
| Russia | RUS | UMIC | 40.82 | 8.12 | 119.40 | 339.33 | 124.87 | 723.50 |
| Rwanda | RWA | LIC | 4.37 | 0.98 | 12.15 | 14.71 | 5.68 | 30.48 |
| Sudan | SDN | LIC | 16.24 | 3.32 | 46.89 | 67.49 | 24.76 | 144.16 |
| Senegal | SEN | LMIC | 5.88 | 1.23 | 16.81 | 21.67 | 8.66 | 43.97 |
| Solomon Islands | SLB | LMIC | 9.02 | 1.77 | 26.57 | 37.71 | 14.45 | 78.47 |
| Sierra Leone | SLE | LIC | 4.86 | 1.07 | 13.55 | 14.70 | 5.91 | 29.71 |
| El Salvador | SLV | LMIC | 18.15 | 4.06 | 50.30 | 103.06 | 42.23 | 205.88 |
| Serbia | SRB | UMIC | 28.93 | 6.50 | 80.05 | 199.69 | 79.21 | 407.00 |
| Sao Tome and Principe | STP | LMIC | 7.60 | 1.69 | 21.17 | 29.25 | 11.60 | 59.61 |
| Suriname | SUR | UMIC | 32.61 | 6.88 | 92.91 | 191.26 | 77.38 | 385.08 |
| Eswatini | SWZ | LMIC | 12.46 | 2.20 | 38.37 | 65.95 | 26.11 | 134.58 |
| Syria | SYR | LIC | 17.09 | 3.40 | 49.96 | 90.08 | 35.29 | 184.99 |
| Chad | TCD | LIC | 4.30 | 0.96 | 11.92 | 14.70 | 5.74 | 30.25 |
| Togo | TGO | LIC | 3.65 | 0.79 | 10.26 | 10.96 | 4.66 | 21.39 |
| Thailand | THA | UMIC | 21.92 | 4.50 | 63.19 | 141.95 | 56.26 | 289.47 |
| Tajikistan | TJK | LMIC | 4.34 | 1.01 | 11.84 | 16.63 | 6.72 | 33.51 |
| Turkmenistan | TKM | UMIC | 22.52 | 4.83 | 63.68 | 137.81 | 52.20 | 288.83 |
| Timor-Leste | TLS | LMIC | 4.89 | 0.99 | 14.21 | 17.10 | 6.63 | 35.35 |
| Tonga | TON | UMIC | 15.52 | 3.29 | 44.12 | 75.82 | 29.62 | 155.97 |
| Tunisia | TUN | LMIC | 22.07 | 4.37 | 64.71 | 144.64 | 54.21 | 305.07 |
| Turkey | TUR | UMIC | 31.58 | 6.84 | 88.87 | 243.00 | 95.11 | 499.31 |
| Tuvalu | TUV | UMIC | 8.42 | 1.92 | 23.16 | 37.09 | 14.25 | 77.06 |
| Tanzania | TZA | LMIC | 4.35 | 0.94 | 12.25 | 14.63 | 5.55 | 30.64 |
| Uganda | UGA | LIC | 4.21 | 0.86 | 12.15 | 14.12 | 5.48 | 29.16 |
| Ukraine | UKR | LMIC | 12.91 | 3.12 | 34.50 | 74.50 | 29.80 | 151.05 |
| Uzbekistan | UZB | LMIC | 5.64 | 1.19 | 16.05 | 25.23 | 9.74 | 52.29 |
| St. Vincent & Grenadines | VCT | UMIC | 27.69 | 5.95 | 78.20 | 191.79 | 73.38 | 399.54 |
| Vietnam | VNM | LMIC | 10.70 | 2.27 | 30.43 | 47.42 | 19.26 | 95.26 |
| Vanuatu | VUT | LMIC | 11.63 | 2.62 | 32.16 | 59.11 | 23.82 | 119.31 |
| Samoa | WSM | LMIC | 13.29 | 2.82 | 37.73 | 67.06 | 26.68 | 136.42 |
| Yemen | YEM | LIC | 8.01 | 1.81 | 22.09 | 33.96 | 13.64 | 68.70 |
| South Africa | ZAF | UMIC | 25.19 | 5.63 | 69.83 | 165.62 | 64.93 | 339.98 |
| Zambia | ZMB | LIC | 4.31 | 0.99 | 11.78 | 15.58 | 6.00 | 32.34 |

### 2 Appendix Results

#### 2.1 Direct and indirect costs for diarrheal and RSV diseases

Data extraction tables for diarrheal and RSV diseases are presented in Table 2.1 and Table 2.2 , respectively. Six studies (in grey shade) focusing on diarrheal disease were excluded for the comparison with WHO-CHOICE estimates because of the following reasons: the inpatient and outpatient costs were pooled (N=4); study reported outputs of another included study (by same authors and same study period, but different perspectives) (N=1); or focused only on intensive care unit (ICU) and high independency unit costs (N=1). For RSV, one study (in grey shade) focusing on the ICU costs was excluded from the comparison.

Table 2.1: Studies reporting direct and indirect costs for diarrheal disease in children in low- and middle-income countries. Studies with grey background colours were excluded from the comparison.

| Reference | Country (ISO3) | Income group | Disease | Health facility | Sample size | Perspective | Study period | Currency & cost year | Direct medical cost (95% CI) [IQR*^#^*] | Direct non-medical costs | Indirect costs | Total costs (if indirect costs not reported) |
| --- | --- | --- | --- | --- | --- | --- | --- | --- | --- | --- | --- | --- |
| Pradhan et al (2020)^6^ | India (IND) | LMIC | Diarrhea | A tertiary hospital | Inpatient: n=60  Hospital outpatient: n= 60 | Societal | Jan-April 2018 | Cost year unspecified  Currency: Rupee (INR) | **Outpatient**:  Mean: Rs 795 (SD: 203)  Median: Rs 778.5 (IQR: 263)  Min-Max: Rs 370-1206  **Inpatient**  Mean: Rs 5110 (SD: 4018)  Median: Rs 3823 (IQR: 1942)  Min-Max: Rs 1774-17426 | **Outpatient**:   - Transportation:   Mean: 395 (SD 643), median: 230 [IQR: 172], min-max: 85-3600   - Food:   Mean: 74(68)  Median: 45 [81]  Min-max: 0-260   - Other nonmedical:   Mean: 98 (83),  Median: 80 [109]  Min-max: 0-335  **Inpatient**:   - Transportation:   Mean: 1172 (1371), median: 350 [1808]  Min-max: 0-4150   - Food:   Mean:1103 (751),  Median:1000 [799]  Min-max120-3923   - Other nonmedical   Mean: 135 (184)  Median: 100 [135]  Min-max: 0-950 | **Outpatient:**   - Income lost:   Mean: 95 (SD: 25)  Median: 0 [IQR: 0] min-max: 0-720  **Inpatient:**   - Income lost:   Mean: 837 (1170) median: 0 [1500]  Min-max: 0-950 |  |
| Charoenwat et al (2022) ^7^ | Thailand (THA) | UMIC | Diarrhea | National data at hospital level | 579,674 | National healthcare payer | 01/2015-12/2019 | 2015-2019  Thai baht (THB) | **Inpatient:**  Re-estimated average cost per admission  2015: THB: 3492  2016: THB: 3907  2017: THB: 3993  2018: THB: 4033  2019: THB: 4054 |  |  |  |
| Mujuru et al (2020) ^8^ | Zimbabwe (ZWE) | LIC | Diarrhea including rotavirus | A tertiary hospital | 202 | Societal | 06/2018-04/2019 | 2019  USD | Inpatient: median $ 251.74  [IQR: 155.42, 390.96]  RV positive: median $203.56 [137.92, 313.92] | $ 22.50 [15-32] |  | $293.74 [188.42, 427.89]  RV positive: $243.78 [160.92, 323.84] |
| Aliabadi et al (2020)^9^ | Burkina Faso (BFA) | LIC | Rotavirus diarrhea | 2 sentinel hospitals | 211 | Societal | 12/2017-06/2018 | 2018  USD (adjusted for local currency + transaction fees) | **Inpatient**:  Median [IQR]  Government: $75 [59, 97]  Household: $24 [12, 49]  Total: $102 [81, 146] | **Inpatient**:  Median [IQR]  Government: 0 [0, 0]  Household: 14 [8, 22]  Total: 14 [8, 22] |  |  |
| Pempa et al (2020) ^10^ | Bhutan (BTN) | LIC | Rotavirus diarrhea | National health care service costing | Not reported | Government | Not reported | USD | **Outpatient**: $6.82  (range 6.01-6.63)  **Inpatient**: $146.83 (range 74.39-238.14) |  |  |  |
| Debellut (2020) ^11^ | Palestine (PSE) | LIC | Rotavirus diarrhea | MOH data | Not reported | Health system and societal perspective | Not reported | 2018  USD | MOH unit cost  **Outpatient**: $7.63  **Inpatient**: $173.85 per day |  |  |  |
| Constenla et al (2019) ^12^ | India (IND) | LMIC | Rotavirus diarrhea | Cost data based on Census data India | Not reported | Societal | Not applicable (expert discussion panel) | 2018  USD | **Outpatient:**  Primary health center:   - Public $2 - Private $8 (6-9)   Secondary hospital:   - Public $4 (3-8) - Private $9 (6-13)   **Inpatient**  Cost per bed day (secondary hospital)   - Public $57 (range $45-68) - Private $246 (range $197-296) |  |  |  |
| Mosegui et al (2019) ^13^ | Brazil (BRA) | UMIC | Diarrhea | MOH guidebook | Not reported | Brazilian public health  System | Not reported | USD (year not reported) | **Outpatient:**  Medical visit: $10  Nurse visit: $10  **Inpatient:** $324.9 |  |  |  |
| Zimmermann et al (2019) ^14^ | 7 countries | LIC and LMIC | Diarrhea/ rotavirus diarrhea | Sentinel hospitals | 2193 | Societal | 12/2007-03/2011 | 2012 USD | **Inpatient:**  Mean: $18.7 (SD: 21.45)  Median: $12.82  **Outpatient:**  Mean: $10.52 (SD: 23.69)  Median: $5.41 | Transportation:  Mean: $0.94 (SD: 8.67)  Median: $0 | Lost earning  Mean: $2.7 (SD: 5.71)  Median: $1.21 | Mean: $12.83 (SD: 23.37)  Median: $7.4 |
|  | The Gambia (GMB) | LIC |  |  |  |  |  |  | **Inpatient:**  Mean: $14.52 (SD: 29.57)  Median: $8.02  **Outpatient:**  Mean: $4.39 (SD: 3.41)  Median: $3.66 | Transportation:  Mean: $0.66 (SD: 2)  Median: $0 | Lost earning  Mean: $4.59 (SD: 16.67)  Median: $1.4 | Mean: $7.62 (SD: 17.52)  Median: $4.15 |
|  | Mali (MLI) | LIC |  |  |  |  |  |  | **Inpatient:**  Mean: $169.89 (SD: 53.02)  Median: $167.36  **Outpatient:**  Mean: $20.88 (SD: 19.79)  Median: $14.56 | Transportation:  Mean: $0.43  Median: $0  SD: 1.7 | Lost earning  Mean: $2.89 (SD: 10.71)  Median: $0.07 | Mean: $22.66 (SD: 26.02)  Median: $14.59 |
|  | Mozambique (MOZ) | LIC |  |  |  |  |  |  | **Inpatient:**  Mean: $10.34 (SD: 10.23)  Median: $7.34  **Outpatient:**  Mean: $ 4.44 (SD: 15.75)  Median: $2.65 | Transportation:  Mean: $1.06 (SD: 3.98**)**  Median: $0 | Lost earning  Mean: $4.32 (SD: 6.43)  Median: $2.06 | Mean: $7.15 (SD: 13.8)  Median: $3.34 |
|  | Kenya (KEN) | LMIC |  |  |  |  |  |  | **Inpatient:**  Mean: $33.1 (SD: 19.34)  Median: $28  **Outpatient:**  Mean: $15.92 (SD: 8.44)  Median: $14.3 | Transportation:  Mean: $0.23 (SD: 0.85)  Median: $0 | Lost earning  Mean: $14.75 (SD: 11.2)  Median: $11.6 | Mean: $18.7 (SD: 12.62)  Median: $15.14 |
|  | India (IND) | LIMC |  |  |  |  |  |  | **Inpatient**:  Mean: $19.53 (SD: 11.59)  Median: $16.74  **Outpatient**:  Mean: $6.29 (SD: 4.34)  Median: $5.13 | Transportation:  Mean: $1.03 (SD: 1.37)  Median: $0.47 | Lost earning  Mean: $4.62 (SD: 7.59)  Median: $0.13 | Mean: $11.27 (SD: 10.16)  Median: $6.84 |
|  | Bangladesh (BGD) | LMIC |  |  |  |  |  |  | **Inpatient:**  Mean: $19.8 (SD: 13.66)  Median: $13.51  **Outpatient:**  Mean: $9.82 (SD: 56.51)  Median: $4.71 | Transportation:  Mean: $1.65 (SD: 2.3)  Median: $0.79 | Lost earning  Mean: $5.09 (SD: 10.39)  Median: $0.59 | Mean: $15.59 (SD: 38.38)  Median: $9.5 |
|  | Pakistan (PAK) | LMIC |  |  |  |  |  |  | **Inpatient:**  Mean: $7.1 (SD: 0.97)  Median: $7.1  **Outpatient:**  Mean: $6.2 (SD: 23.94)  Median: $1.71 | Transportation:  Mean: $1.26  Median: $0  SD: 22.86 | Lost earning  Mean: $2.24 (SD: 5.88)  Median: $0.11 | Mean: $6.2 (SD: 23.87)  Median: $1.71 |
| Rochanathimoke et al (2019) ^15^ | Thailand (THA) | UMIC | Rotavirus diarrhea | 3 general and 9 district hospitals | 197 | Societal |  | 2014  USD | Mean: $612.37  (95% CI: 550.84-673.90)  Median $792.93 | Travel, food, diapers and other  Mean: $113.91  (95% CI 99.01-128.82)  Median $84.88 | Informal care  Mean: $173.72  (95% CI 155.44-191)  Median: $148.43 | Mean: $903.39  (95% CI $820.46-986.31)  Median: $747.41 |
| Hovhannisyan (2019) ^16^ | Armenia (ARM) | UMIC | Rotavirus diarrhea | A tertiary hospital | 126 | Not reported | 01/2014-07/2014 | Year not reported (assume (2014), Armenian dram (AMD) | Mean: AMD 12968 (calculated:1634000/126) |  |  |  |
| Nonvignon (2018) ^17^ | Ghana (GHA) | LIC | Rotavirus diarrhea | National health insurance scheme 2015 tariff | Not reported | Health care system and societal | Not applicable | 2015 USD | **Inpatient:**  Government / household  Private hospital: $35.02 / $15.67  Public district hospital: $22.71 / $10.17  Public regional hospital: $37.85 /$16.94  Public teaching hospital: $48.57/$21.74  **Outpatient**  Government / household  Private clinics: $3.59 / $1.61  Private hospital: $3.77 /$1.69  Public health centre: $1.61 /$0.72  Public district hospital: $2.47/ $1.10  Public regional hospital: $3.05 /$1.36  Public teaching hospital: $6.15/$2.75 |  |  |  |
| Anwari et al (2018) ^18^ | Afghanistan (AFG) | LIC | Rotavirus diarrhea | Basic package of health service assessment | Not reported | Government and societal | Not applicable | 2016 USD | **Inpatient:**  Government/household: $8.8/ $7.14  **Outpatient:**  Non-severe:  Government/household: $1.72/1.97  Severe:  Government/household: $1.72/7.08 |  |  | Societal cost:  Inpatient: $15.94  **Outpatient:**  Non-severe: $3.69  Severe: $8.80 |
| Saker et al (2018) ^19^ | Bangladesh (BGD) | LMIC | Diarrhea | Tertiary hospitals | Inpatient: 402  Outpatient: 399 | Societal | 01/2015-12/2015 | 2015 BDT | Inpatient: BDT 2104.09  Outpatient: BDT 85.97 |  |  | Inpatient: BDT 6570.79 (SD: 5457.71)  Outpatient: BDT 1767.58 (SD: 3465.92) |
| Memirie et al (2017)^20^ | Ethiopia (ETH) | LIC | Diarrhea | 6 public hospitals, 15 public health centres, 9 health posts, 5 private health facility | Diarrheal: 309  Severe:32 | Household | 08/2013-12/2013 | 2013 USD | Medical household expenditure  Non-severe: $6.24 (SD:11.8)  Severe: $79.27 (SD: 74.38) | Transportation:  Non-severe: $0.99 (SD: 3.30)  Severe: $9.64 (11.02) |  | **Non-severe**:  Health post: $0.97 (SD:2.71)  Health centre: $3.89 (6.13)  Government hospital: $5.66 (5.97)  Private clinical/hospital: $21.41 (11.17)  **Severe**:  Health centre: $15.59  Government hospital: $55.92 (58.96)  Private clinical/hospital: $151.86(84.33) |
| Hendrix et al 2017 ^21^ | Malawi (MWI) | LIC | Diarrhea | A tertiary hospital (urban) and a primary hospital (rural) | Urban: 269  Rural: 22 | Health system and societal | 01/01/2013-21/11/2014 | Malawi Kwcha and USD 2014 | **Inpatient**  Rural/urban: $55.04 (SD: 26.82) / $47.21 (SD 56.69)  **Outpatient**  Rural/urban: $8.08 (SD: 2.29)/ $7.18 (SD 4.05) | Household  **Inpatient**  Rural/urban: $19.16 (SD: 19.30) / $25.36 (SD: 20.47)  **Outpatient**  Rural/urban: $1.81 (SD: 2.58)/ $15.48 (SD 20.75) |  | **Inpatient**  Rural/urban: $76.94 (SD: 32.61) / $73.78 (SD:51.39)  **Outpatient**  Rural/urban: $13.57(SD: 4.70)/ $23.13 (SD: 21.68) |
| Ngabo et al (2016) ^22^ | Rwanda (RWA) | LIC | Diarrhea | 2 public district and 1 teaching hospital | 203 | Health insurance and patients | 11/2013-06/2014 | USD 2014 | Inpatient:  Mean: $44.22 (SD 22.74)  Median: $37.68 | Transportation:  Patients/caregivers: mean $2.06 (SD: 2.56)  Median: $1.20  Visitors:  Mean: $12.79 (SD: 21.79)  Median: $5.99 | Lost income  Mean: $41.78 (SD: 54.44)  Median: $26.95 |  |
| Ruhago et al (2015) ^23^ | Tanzania (TZA) | LIC | Diarrhea | District hospitals | Not reported | Health service provider |  | USD 2012 | Inpatient:  Weighted (urban/rural) average: $8.9 per **day** (average 4 days)  Outpatient:  Weighted (urban/rural) average: $3.8 |  |  |  |
| Soltani et al (2015) ^24^ | Tunisia (TUN) | LMIC | Rotavirus diarrhea | 5 sentinel hospitals | GE: 279  Rotavirus: 65 | Health system | 06/2009-05/2011 | Tunisian dinar (TND) and USD | Inpatient:  Rotavirus: TND 433 (SD: 134) |  |  |  |
| Alkoshi et al (2015) ^25^ | Libya (LBY) | LIC | Rotavirus diarrhea | 3 (referral?) Hospital | 239 | Hospital and patient | 08/2012/04/2013 | USD (year unclear, use 2013) | Hospital perspective: $488.12 (SD: 427.91) [range 85-4615] | Patient perspective  Transportation: $78.42 (SD 69.61) [range 8-773]  Household:  $70.09 (SD 69.46) [16-472] | Lost income: $42.37 (143.69) [0-1223] | $678.99 (SD: 499.12) ($200-5,423) |
| Phavichitr et al (2013) ^26^ | Thailand (THA) | UMIC | Diarrhea, including rotavirus | A tertiary hospital | Control arm: 53 | Health care providers and societal? | 04/2010-09/2011 | Thai Baht (use 2011) | Placebo group:  Median: THB: 4778.75 [IQR: 3639.25-6779.50] |  |  |  |
| Alvis-Guzman et al (2013) ^27^ | Colombia (COL) | UMIC | Diarrhea | Primary, secondary, and tertiary | 1452 | Healthcare system | 2010 database | USD 2010 | Inpatient median cost  Primary: $25.1 [IQR: 17.8-45.9]  Secondary: $34.8 [13.3-169.7]  Tertiary: $175.6 [55.7-357.8]  Critical care: $7184.2 [2270.6-13,672] |  |  |  |
| Sowmyanarayanan (2012) ^28^ | India (IND) | LMIC | Diarrhea, including rotavirus | 10 hospitals, including general /governmental hospitals and a private hospital | 211 | Societal | 11/2008-02/2009 | Indian Rupees (INR) (2009) | Rotavirus positive hospitalised:  Mean: INR 2306 [IQR: 50-9249] |  | Lost wage  INR 314 [0-5000] | Mean: INR 2956 [50-13690] |
| Osano et al (2011) ^29^ | Kenya (KEN) | LMIC | Rotavirus diarrhea | Kenyatta National Hospital (KNH) | 172 | Hospital and societal | 02/2008- 05/2008 | 2008  Kenyan Shilling (KES) | Inpatient admission to discharge: mean cost  Hospital perspective:  KNH: KES 5919.03  National insurance fund (NHIF): KES 14178.21  WHO-CHOICE: KES15038.22 |  | Patient perspective: KES 6505.79 | Societal perspective:  KHN KES 8296.90  NHIF: KES 16556.08  WHO-CHOICE: KES 17416.09 |
| Latipov et al (2011) ^30^ | Kazakhstan (KAZ) | UMIC | Rotavirus diarrhea | Sentinel hospitals | Unknown | Healthcare system and societal | Not applicable | USD 2009 | **Severe cases**:  Health care system: $271.95  **Moderate cases**:  Health care system: $27 |  | **Severe cases**:  Caregivers: $191.64  **Moderate cases**:  Caregivers: $54.6  **Mild cases:**  Caregivers: $21.11 | **Severe cases**:  $454.49  **Moderate cases**: $81.60  **Mild cases:** $21.11 |
| Flem et al (2009) ^31^ | Kyrgyzstan (KGZ) | LMIC | Rotavirus diarrhea | 2 public hospitals | 175 | Health care system and societal | 02/2008-04/2008 | USD 2008 | **Inpatient cost**: mean $39  Of which, Bed days: $26.4 [23.6-29.2] | Transportation $14.5 [range 0-23.67]  Self-provision of food: $24.9 [0-165.1] | $2.5 [24.9-66.5] | $87 [25-666] |
| Mendelsohn et al (2008) ^32^ | India (IND) | LMIC | Rotavirus diarrhea | A tertiary hospital, a community hospital and a government clinic | Tertiary: 114  Community: 18 | Health care system and patient | 11/2005-12/2006 | Indian Rupees (INR) (assume 2006) | **Inpatient cost**:  Tertiary: median: INR 2745.3 [IQR: 1944.9-4210.1]  Community: median: INR 537.9 [393.9-745.4]  **Emergency room**:  Tertiary: median: INR 384.8 [IQR: 223.1-806.3]  Community: median: INR77.8 [56.9-152.3]  **Outpatient cost**:  Tertiary: median: INR 133.4 [82.1-195.4]  Community: median: INR 75.5 [46.9-126.9] | **Inpatient cost**:  Tertiary: median: INR 40 [IQR: 13.9-82.5]  Community: INR: median INR: 39.5 [19.8-124.8]  **Emergency room**:  Tertiary: median INR: 62.5 [28.5-120]  Community: median INR: 16 [8-63]  **Outpatient cost**:  Tertiary: median: INR 12.8 [4.8-40]  Community: median: INR 21 [0-46.3] |  |  |
| Wilopo et al (2009) ^33^ | Indonesia (IDN) | LMIC | Rotavirus diarrhea | Healthcare centres and district-level hospital | Inpatient: 891  Outpatient: 458 | Healthcare system and societal | Survey year 2007 | USD 2007 | **Inpatient:**  Mean $49.66 (95%CI: 47.20-52.10)  **Outpatient:**  Mean $5.13 (3.75-6.51) | **Inpatient:**  Mean $8.69 (95%CI: 8.11-9.27)  **Outpatient:**  Mean $0.36 (0.29-0.43) | **Inpatient:**  Mean $8.71 (95%CI: 7.87-9.55)  **Outpatient:**  Mean $1.28 (0.88-1.69) | **Inpatient:**  Mean $67.06 (95%CI: 64.19-69.92)  **Outpatient:**  Mean $6.78 (5.30-8.26) |
| Rheingans et al (2012) ^34^ * | Bangladesh (BGD) | LMIC | Diarrhea | Healthcare utilization survey | 95 | Household | Not reported | USD, 2011 | Seeking health care: $0.94 (SD: 0.16) | $0.25 (SD: 0.07) | $0.63 (SD: 0.22) | $1.82 (SD: 0.34) |
|  | India (IND) | LMIC |  |  | 92 |  |  |  | Seeking health care: $2.08 (SD: 0.39) | $0.25 (SD: 0.07) | $1 (SD: 0.25) | $3.33 (SD: 0.6) |
|  | Pakistan (PAK) | LMIC |  |  | 349 |  |  |  | Seeking health care: $2.30 (SD: 0.53) | $0.25 (SD: 0.07) | $3.93 (SD: 2.07) | $6.47 (SD: 2.16) |
| Rheingans et al (2012) ^35^ * | Gambia (GMB) | LIC | Diarrhea | Healthcare utilization survey | 259 | Household | Not reported | USD, 2011 | Seeking health care: $0.71 (SD: 0.16) | $0.37 (SD: 0.07) | $1.55 (SD: 0.42) | $2.63 (SD: 0.53) |
|  | Kenya (KEN) | LMIC |  |  | 275 |  |  |  | Seeking health care: $0.70 (SD: 0.09) | $0.55 (SD: 0.28) | $4.99 (SD: 1.41) | $6.24 (SD: 1.45) |
|  | Mali (MAL) | LIC |  |  | 126 |  |  |  | Seeking health care: $2.20 (SD: 0.44) | $0.19 (SD: 0.05) | $1.72 (SD: 0.40) | $4.11 (SD: 0.67) |
| Mathew et al (2016) ^36^ * | India (IND) | LMIC | Rotavirus diarrhea | A tertiary hospital | 84 | Not reported | 2005-2008  And 11/2012-01/2014 | Indian Rupee (INR), year not reported | **ICU**  2005-2008/2012-2014  RV+: median: INR 17,941 [IQR:13951-22930] / 50663 [19366-65324]  Rv-: median: INR 11,614 [10408-16952] / 27106 [14634-30909]  **High dependency unit**  2005-2008/2012-2014  RV+: median INR 5957 [IQR:4732-8709] / 10903 [7723-16300]  Rv-: median: INR 5787 [4169-7461] / 10088 [7356-12814] |  |  |  |
| Riewpaiboon et al (2016) ^37^ * | Vietnam (VTM) | LMIC | Rotavirus diarrhea | Hospital and outpatient clinics | 557 episodes | Societal | Nested surveillance 2009 | USD 2014 | Direct medical cost  RV-: $75.12(SE: 6.4) median: $16.37  RV+: $117.71 (SE 7.92) median $104.55  Total: $87.13 (SE 4.68) median 16.74 | RV-: $21.87(SE: 1.15) median: $18.47  RV+: $22.09 (SE: 1.27) median $18.75  Total: $21.79 (SE: 18.47) median $18.47 | RV-: $60.73(SE: 2.32) median: $48.95  RV+: $77.03 (SE 2.97) median $65.27  Total: $65.74 (SE: 1.73) median $57.11 | RV-: $157.72(SE: 8.17) median: $97.97  RV+: $216.83 (SE 10.37) median $196.44  Total: $174.66 (SE 6.06) median $107.38 |
| Burnett et al (2020) ^38^ | Madagascar (MDG) | LIC | Diarrhea, including rotavirus | A public pediatric referral hospital | 96 children | Societal | 05/2018-11/2018 | Malagasy Ariary and USD 2019 | Used CHOICE data to estimate the inpatient costs  LoS: 3 days (IQR 2-5 days)  Median hospitalisation cost: $83.92 (IQR: 59.13-122.6)  93% received ORS with IV  Medication costs in hospital: median: 23.33 (IQR: 11.70-43.48): mean 38.82 (SD 51.35)  Diagnostic test: Median: 18.64 (12.73-23.67) Mean: 18.58 (SD 8.5)  **Total medical cost:**  Median: $107.22 (IQE: 81.78-143.3)  Mean: $134.69 (SD 110.3) | Median $28.55 (IQR: 10.91-56.12)  mean: $50.06 (SD: 81.71) | Median $18.18 (IQR: 9.09-30.30)  Mean: $22.84 (SD: 19.37) | Median: $156.0 (IQR: 104-210.86)  Mean: $194.03 (SD: 163.72) |
| Jacob et al (2016) ^39^ | India (IND) | LMIC | Diarrhea | A tertiary hospital | Outpatient: 30  Inpatient: 30 | Not reported | 07/2014-09/2014 | Indian Rupee (INR), year not reported (assume to be 2014) | **Inpatient:**  Mean: INR 11,767  Median: INR 7259 [min-max: 2455-47663]  **Outpatient**  Mean: INR 977  Median: INR 591 [min-max: 94-4006] | **Transportation**  **Inpatient:**  Mean: INR 516  Median: INR 135 [min-max: 0-3160]  **Outpatient**  Mean: INR 288  Median INR 135 [min-max: 0-1200]  **Food**  **Inpatient:**  Mean: INR 732  Median: INR 475 [min-max: 0-540]  **Outpatient**  Mean: INR 73  Median: INR 55 [min-max: 0-400] |  | **Inpatient:**  Mean: 13014  Median: 7868 [min-max: 263-49498]  **Outpatient**  Mean: 1338  Median 780 [min-max: 325-6502] |
| Halder et al (2017) ^40^ | Bangladesh (BGD) | LMIC | Diarrhea | Cross sectional survey | Inpatient: 2126 (N for hospitalization incidence)  Outpatient: 808 | Not reported | 09/2007-11/2007 | USD 2007-2008 | **Inpatient:**  Mean: 35.40 (95%CI: 25.46-45.35)  **Outpatient:**  Home care: $0.16 (0.11-0.22)  Qualified practitioner: $2.66(1.58-3.74)  Unqualified: $2.80 (-0.59-6.18)  Both : $3.16 (0.26-6.05) |  |  |  |
| Das et al (2015) ^41^ | Bangladesh (BGD) | LMIC | Diarrhea | A tertiary hospital | 4205, RV+ 1174 | Not reported | 01/2010-12/2012 | USD, year unclear |  |  |  | Overall:  Mean:6.44 (SD: 10.26)  Median 3.04 [range: 0.01-94.35]  RV inpatient  Mean:5.03 (SD: 5.84)  Median 3.08[range: 0.06-48.0] |
| Burke et al (2014) ^42^ | Bolivia (BOL) | LMIC | Diarrhea | 6 healthcare settings across 4 cities | 511 | Family perspective | 2007-2009 | USD 2012 | **With national insurance**  Mean: $8.5 (SEM 1.5)  **Without national insurance**  Mean: $27 (SEM 2.5) | **With national insurance**  Mean: $9.00 (SEM 0.50)  **Without national insurance**  Mean: $5.5 (SEM 0.5) | **With national insurance**  Mean: $18.50 (SEM 1.5)  **Without national insurance**  Mean: $13 (SEM 1.5) | **With national insurance**  Mean: $45.5 (SEM 3.0)  **Without national insurance**  Mean: $25 (SEM 2.0) |
| Burke et al (2013) ^43^ * | Bolivia (BOL) | LMIC | Diarrhea | 6 healthcare settings across 4 cities | 535 (inpatient 250, outpatient 285) | Caregivers’ perspective | 2007-2009 | USD 2012 | **Inpatient**  Urban median: $27.58 [IQR: 5.98-57.89]  Rural median: $26.50 [IQR: 9.07-67.68]  **Outpatients**  Urban median: $4.79 [IQR: 1.87-14.54]  Rural median: $6.91 [IQR: 3.6-18.72] | **Inpatient**  Urban median: $4.03 [IQR 2.63-6.30]  Rural median: $6.34 [IQR 3.31-9.36]  **Outpatients**  Urban median: $2.88 [IQR 1.30-5.62]  Rural median: $4.48 [IQR 2.74-9.5] | **Inpatient**  Urban median: $12 [IQR: 0-28.50]  Rural median: $20.34 [IQR: 11.52-27.60]  **Outpatients**  Urban median: $1.23 [IQR: 0-13.37]  Rural median: $8.64 [IQR: 0-24.48] | **Inpatient**  Urban median: $41.54 [IQR: 10.7-82.19]  Rural median: $10.74 [IQR: 2.45-29.95]  **Outpatients**  Urban median: $10.74 [IQR: 2.45-29.95]  Rural median: $17.63 [IQR: 4.97-36.91] |

** Matthew et al 2016 study was not included in the analysis, because it is mainly focus on intensive care unit and high dependency unit. Burke et al 2014 study was also not included in the analysis, because it is similar analysis as Burke et al 2013 based on same survey. Both Rheingans 2012 studies, Riewpaiboon 2016 and Das 2015 study were also excluded, because they did not explicitly report inpatient and outpatient costs. Burnett et al 2018 study was excluded, because it used the WHO-CHOICE unit cost to estimate the inpatient costs.*

*^#^ Some studies reported IQR as Q3-Q1, whereas others reported the lower and upper quartiles.*

Abbreviations: SE: standard error, SD: standard deviation, CI: confidence intervals, IQR: interquartile range, LIC: low-income countries, LMIC: lower-middle income countries, UMIC: upper-middle income countries, RV: rotavirus. MOH: ministry of health.

Table 2.2: Studies reporting direct and indirect costs for RSV disease in children in low- and middle-income countries. Studies with grey background colours were excluded from the comparison

| Reference | Country (ISO3) | Income group | Disease | Health facility | Sample size | Perspective | Study period | Currency & cost year | Direct medical cost (95% CI) [IQR] | Direct non-medical costs | Indirect costs | Total costs (if indirect costs not reported) |
| --- | --- | --- | --- | --- | --- | --- | --- | --- | --- | --- | --- | --- |
| Tan et al (2023) ^44^ | Thailand (THA) | UMIC | RSV+ | A tertiary hospital | Overall: 1370  Inpatient: 683 | Health care provider | 01/2014-12/2021 | USD 2021 | **Inpatient:**  Median: 2111[IQR: 1379-3182]  **Outpatient:**  Median: $167 [IQR: 90-268]  **Overall**:  Median: $539 [IQR: 167-2106] |  |  |  |
| Moyes et al (2023) ^45^ | South Africa (ZAF) | UMIC | RSV-ARTI | 5 sentinel hospitals | Severe cases: 675, mild cases: 527 | Health care system and societal | 2011-2016 | USD 2022 | **Inpatient**  Cost per episode  0-2 m: $921.66  3-5m: $623.59  6-8m: $583.71  9-11m: $716.68  12-59m: $591.44  **Outpatient**  0-2 months: $24.98  3-5 months: $24.98  6-8 months: $25.20  9-11 months: $25.29  12-59 months: $25.66  Overall: $25 (95% CI: 18.3-31.8) | Out of pocket expense  **Inpatient**  0-2m: $10.66 (1.81-19.48)  3-5m: $11.49 (3.41-19.57)  6-8m: $4.42 (0.31-8.53)  9-11m: $11.73 (1.07-22.39)  12-59m: $22.5 (4.47-45.97)  **Outpatient** 0-2m: $0.57 (0-2.16) 3-5m: $0.57 (0-2.16) 6-8m: $1.28 (0-3.63) 9-11m: $0.71 (0-9.79) 12-59m: $0.86 (0.31-5.10) | **Inpatient**  0-2m: $66.89 (46.82-86.96)  3-5m: $50.72 (35.5-65.94)  6-8m: $46.85 (32.8-60.91)  9-11m: $60.46 (42.32-78.60)  12-59m: $44.27 (30.99-57.55)  **Outpatient**  0-2m: $10.71 (7.50-13.92)  3-5m: $10.71 (7.5-13.92)  6-8m: $10.71 (7.50-13.92)  9-11m: $10.71 (7.5-13.92)  12-59: $10.71 (7.5-13.92) |  |
| Ren et al (2023) ^46^ | China (CHN) | UMIC | RSV-ARI | A tertiary hospital | Inpatient: 261 RSV+  Interviewed 170 |  | 12/202-20/2021 | USD 2021 | **Inpatient**  Overall mean: $1055.3 (CI: 998.2-1112.5)  0-11m: $1152.7 (CI: 1074.5-1230.9)  12-23m: $903.5 (CI: 799.2-1007.7)  24-59m: $875.3 (CI: 809.0-941.5) |  | **Inpatient:**  Overall mean: $83.6 (CI: 77.5-89.7)  0-11m: $91.3 (CI: 83.1-99.5)  12-23m: $66.7 (CI: 57.6-75.9)  24-59m: $74.8 (CI: 61.0-88.7) | **Inpatient:**  Overall mean: $162.4 (CI: 127.9-197.0)  0-11m: $163.6 (CI: 118.7-208.6)  12-23m: $147.0 (68.5-225.4)  24-59m: $175.0 (93.2-256.8) |
| Koltai et al (2023) ^47^ | Kenya | LMIC | RSV-ARTI | Data from moh | Inpatients N=7330  Outpatient N=62604 | Health care system and household | Unknown | USD (assume year 2022) | **Inpatient**  Cost per episode: $102.45  **Outpatient**  $20.91 | Cost to household  Prior to hospitalisation: $34.67 (median 14.23, CI 12.86-56.49)  During hospitalisation: $138.51 (median: 122.7, CI 106.54-170.48)  After hospitalisation: S15.35 (median 1227.7, CI 106.54 -170.48) |  |  |
| Tian et al (2023) ^48^ | China (CHN) | UMIC | Bronchiolitis | 27 tertiary hospitals | 42,928 | Unclear | 01/2016-12/2022 | USD (assume 2022) | Inpatient:  $758.49 (IQR: 601.96-1029.53)  Urban: $748.81 (578.21 - 981.72)  Rural: $836.9 (642.83-1093.31) |  |  |  |
| Sun et al (2022) ^49^ | China (CHN) | UMIC | RSV-ARTI | A tertiary hospital | 7,248 | Unclear (Hospital?) | 10/2014-09/2017 | USD (assume 2022) | **Inpatient**  All children with ARI:  Median charge: $887.5 (IQR: 720.8-1198.6)  <6m: $923.3  6-12m: $785.5  12-24m: $811.7  RSV+  Median: $924.3 |  |  |  |
| Do et al (2022) ^50^ | Vietnam (VNM) | LMIC | RSV-LRTI | A tertiary hospital | **Outpatient**:210  RSV +: 21, RSV- :189  **Inpatient:** 318  RSV+: 84, RSV- :234  **ICU**: 8 RSV+: 0 | Health care system and household | 09/2019-12/2019  A stop during Covid pandemic  10/2020 - 06/2021  10/2021-12/2021 | USD 2022 | **Inpatient**:  Total: median $84 [IQR 50-135]  RSV+: $68 [47-102]  RSV-: 91[59-153]  ICU cost (n=8)  RSV-: $1996 [937-3786]  **Outpatient:**  Total: $22 [IQR: 13-32]  RSV+: $26 [16-23]  RSV-: $21 [13-32] | **Inpatient:**  Total: median $35 [3-87]  RSV+: $27 [3-72]  RSV-: $38 [3-88]  ICU (n=8)  RSV-: $76 [42-112]  **Outpatient:**  Total: $5 [IQR: 2-28]  RSV+: $4 [2-22]  RSV-: $5 [2-28] | **Inpatient:**  Total: median 19 [0-69]  RSV+: 18[0-69]  RSV-: 20 [0-69]  ICU: 85 [6-253]  **Outpatient:**  Total: 11 [2-26]  RSV+: 22 [9-34]  RSV-: 9 [0-25] |  |
| Buendia et al (2021) ^51^ | Columbia (COL) | UMIC | RSV-bronchiolitis | A tertiary hospital | 193 | Societal | 01/2015-12/2016 | USD 2016 | **Inpatient cost** per day: mean: $23.925 (CI: 22.745-25.106)  Medical devices: mean: $10.664 (CI: 10.138-11.190)  ED cost:  Mean: $12.833 (12.2-13.467) |  | **Inpatient:**  Mean $17.236 |  |
| Rodriguez-martinez (2021) ^52^ | Columbia (col) | UMIC | Bronchiolitis | A tertiary hospital | 89 | Healthcare provider | 01/2016-12/2017 | 2017 USD | Hotel cost per unit:  Paediatric ward: $34.7-87.3 per day  PIMC: $156.1-211.7  PICU: $323.1-340.1  **Total inpatient cost:**  Pw (n=57) median: $518.0 [IQR: 217-768]  PIMC (n=25): median $1305.2  PICU (n=7): median $2749.7 [IQR: 1372.7-4156.9] |  |  |  |
| Sam et al (2021) ^53^ | Malaysia (MYS) | UMIC |  | A tertiary hospital | 200 | Health care system and patient | 07/2013 - 07/2015 | USD 2014 | **Inpatient**:  Health care system cost:  Mean: $818 (SD: 538)  Median: $701 [IQR: 527-896] | **Inpatient**  Out-of-pocket costs:  Mean: $49 (SD: 89)  Median $189 [IQR: 140-258] | **Inpatient**  Mean $163 (SD: 448)  Median $98 [IQR: 65-156] |  |
| Comas-Garcia et al (2020) ^54^ * | Columbia (COL) | UMIC | RSV in ICU | A general hospital | 24 | Unclear (hospital?) | 01/2016-12/2017 | USD (assume 2010) | **ICU cost per episode**:  Mean: $6,922.30 ($4,716.5-$12,355.4) |  |  |  |
| Baral et al (2020) ^55^ | Malawi (MWI) | LIC | RSV-ARTI | A tertiary hospital | ARTI: 426  RSV+:78  RSV-: 202 | Health care system and household | 05/2015-09/2016 | 2018 USD | **Inpatient**  All infants: mean $55.78 (CI: 49.35-62.21), median: $41.50  RSV+: mean: $45.37 (CI: 38.74-52.00), median: $41.47  RSV-: mean: $55.85 (47.14-64.57), median: $43.12  **Outpatient**  All infants: mean: $7.84 (CI: 6.68-9.01), median $6.52  RSV+: mean: $9.26 (CI:7.34-11.18), median: 9.78  RSV-: mean: $9.74 (7.33-12.14), median: $9.69 | **Inpatient**:  Household (including indirect cost):  All infants: $14.15 (CI: 11.94-16.37), median: $10.09  RSV+: mean: $16.89 (CI: 7.89-25.89), median: $8.35 /  RSV-: mean $12.57 (CI: 10.95-14.19), median: $10.07  **Outpatient**  All infants: mean: $2.33 (CI: 1.63-3.03) median: $0.67  RSV+: mean: $3.25 (CI: 0.10-6.41), median: 1.11  RSV-: mean: $ 2.04 (CI: 1.19-2.89), median $0.75 | Inpatient:  All infants: mean: $6.83 (CI: 4.85-8.81), median: $3.70  RSV+: mean: $9.28 (CI: 0.6-17.95), median: $2.78  RSV-: mean: $5.95 (CI: 5.07-6.80), median: $4.63  **Outpatient:**  All infants: mean: $1.40 (CI: 0.89-1.91) median: $ 0.42  RSV+ : mean: $2.25 (CI: -0.97-5.48), median:$ 0.56  RSV-: mean: $1.14 (CI: 0.57-1.71), median: 0.37 |  |
| Bhuiyan et al (2017) ^56^ | Bangladesh (BGD) | LMIC | RSV+ | 3 public and 1 private hospital | RSV+: 47 | Societal | 05/2010-10-2010 | USD 2010 | **Inpatient:**  Public hospital: median: $48 [IQR: 36-64]  Private hospital: median: $90 [IQR: 71-112]  Total: median $62 [IQR 43-101] |  | Productivity losses:  Median: 12 days [IQR 8-17 days]  Per day: $1.4 [IQR:1.4-1.6]  Total: $19 [IQR: 11-29] |  |
| Marcone et al (2015) ^57^ | Argentina (ARG) | UMIC | Viral respiratory infection | Hospital records from claim database | Total: 1729  Inpatient: 89  ED visits: 8925 | Healthcare provider | 06/2008-12/2010 | USD 2010 | **Inpatient:**  Median: $529 [IQR 362-789]  Room charge: $310 [227-517]  Consumables $66 [38-129]  Diagnostic: $64 [54-99]  Medication: $54 [27-148]  Specialist consultation: $30 [0-60] |  |  |  |
| Zhang et al (2013)^58^ | China (CHN) | UMIC | RSV-ARTI | A tertiary hospital | 2,721 | Healthcare provider | 01/2005-12/2009 | USD (assume 2012, exchange rate reported) | **Inpatient**  By age  <6m: median $538.55 [IQR: 416.17-691.53]  7-24m: $608.39 [IQR: 474.52-789.84]  25m+: $645.77 [IQR: 509.20-840.66]  Overall cost:  Median: $571.84, [min-max: 45.38-4237.08]  ICU: 909.65 [IQR: 497.61-1367.60]  General ward: 565.36 [IQR 436.62-717.82] |  |  |  |
| Chan et al (2003)^59^ | Malaysia (MYS) | UMIC | RSV+ | A tertiary hospital | 216 | Healthcare provider? | 01/1995-12/1997 | USD 1997 | **Inpatient:**  Median: $169.99 [IQR: 128.08-248.47]  PICU: median: $1297.11 [IQR: 648.55-2806.18]  **Total cost (including PICU):** normal term:  Mean $216.55 (SE: 22.94) (CI: 171.27-261.83)  Premature/underlying condition:  Mean: $2485.7 (SE: 682.46) (CI: 1120.78-3850.62) |  |  |  |
| Bhuket et al (2002) ^60^ | Thailand (THA) | UMIC | RSV-LTI | Cross sectional study, interview caregivers | LRI: 165 | Patient | 03/2000-02/2001 | assume 2002  Thai baht (THB) | **Inpatient:**  Mean: THB 1134  **Outpatient:**  Mean: THB 312 | Inpatient: THB 387  Outpatient: THB 118 | Inpatient: THB 222  Outpatient: THB 73 | Mean: THB 1248  Range: 140-6471  Severe pneumonia: THB 2348  Bronchitis: THB 923 |

** Comas-Garcia et al 2022 were excluded from meta-analysis, because it focussed on ICU cost of nosocomial infections*

Abbreviations: SE: standard error, SD: standard deviation, CI: confidence intervals, IQR: interquartile range, LIC: low-income countries, LMIC: lower-middle income countries, UMIC: upper-middle income countries, ARTI: actuate respiratory tract infections, LRI: lower respiratory infection, (P)ICU: (pediatric) intensive care unit

#### 2.2 Length of hospital stay data for diarrheal and RSV diseases

The length of stay (LoS) data extraction tables for diarrheal and RSV diseases are presented in Table 2.3 and Table 2.4, respectively. For diarrhea, more than half of the studies only reported median LoS, and for RSV, only one study from Colombia reported sample size, mean and standard deviation (SD). A South African study reported age-specific mean LoS, but the age-specific sample size, SD and 95% confidence intervals (CIs) were unknown ^45^.

For both diseases, studies that did not report either sample size, SD or 95% CI were excluded from the meta-analysis. For the studies that only reported median LoS, we first converted median to mean using the R package “estmeansd”. Then we performed a meta-analysis using the “metagen” R package with the inverse variance method, and stratified the outcome by LIC-LMIC vs. UMIC. For diarrheal disease, a sensitivity analysis was also performed which included only mean LoS values.

For diarrheal disease, the pooled LoS using the random effect model was 4.38 (95% CI: 3.77-4.98) days (Figure 2.1). This estimate was lower than both analysis that used only mean LoS (Figure 2.2) and rotavirus-specific diarrhea (Figure 2.3), but the 95% CIs largely overlapped.

For RSV disease, the pooled LoS using the random effect model was 5.86 (95% CI: 4.75-6.97) days (Figure 2.4). Only two studies included in the base-case analysis provided data on mean LoS.

Table 2.3: Length of stay (LoS) for childhood diarrheal disease in low- and middle-income countries

| Reference | Country (ISO3) | Income group | Disease | Health facility | Sample size | Mean LoS in days (SD) [95% CI] | Median LoS [Interquartile range range] | Included in meta-analysis |
| --- | --- | --- | --- | --- | --- | --- | --- | --- |
| Aliabadi et al (2020) ^9^ | Burkina Faso (BFA) | LIC | Diarrheal | 2 sentinel hospitals | 211 |  | 4 [3-6] | Base  case |
| Burnett et al (2020) ^38^ ^#^ | Madagascar (MDG) | LIC | Diarrheal | a tertiary hospital | 96 |  | 3 [2-5] | Base  case |
| Pradhan et al (2020) ^6^ | India (IND) | LMIC | Diarrheal | a tertiary | 60 |  | 5 [3-6] | Base  case |
| Mujuru et al (2020) ^8^ | Zimbabwe (ZWE) | LIC | Diarrheal | a tertiary hospital | 202 |  | 3 [2-6] | Base  case |
| Zimmermann et al (2019) ^14^ | 7 countries:  Kenya (KEN),  Mali (MLI),  Mozambique (MOZ),  The Gambia (GMB),  Bangladesh (BGD),  India (IND)  Pakistan (PAK) | LIC and LMIC | Diarrheal | sentinel hospitals | 2193 | Overall  3.6 (2.8) | Overall  3 | Base case and SA |
| Rochanathimoke et al (2019) ^15^ | Thailand (THA) | UMIC | Rotavirus diarrheal | 3 general and 9 district hospitals | 197 | 4.13 (1.95) | 4 | Base case and SA |
| Hovhannisyan (2019) ^16^ | Armenia (ARM) | UMIC | Rotavirus diarrheal | a tertiary hospital | 126 |  | 6.5 [4-9] | Base  case |
| Saker et al (2018) ^19^ | Bangladesh (BGD) | LMIC | Diarrheal | tertiary hospitals | 402 | 5 |  | No, no SD |
| Memirie et al (2017) ^20^ | Ethiopia (ETH) | LIC | Diarrheal | 6 public hospitals | 32 | 3 |  | No, no SD |
| Hendrix et al (2017) ^21^ | Malawi (MWI) | LIC | Diarrheal | a tertiary hospital (urban) and a primary hospital (rural) | Urban: 269  Rural: 22 |  | Urban: 3[2]  Rural: 2 [2.5] | Base case |
| Ngabo et al (2016) ^22^ | Rwanda (RWA) | LIC | Diarrheal | 2 public district and 1 teaching hospital | 203 | 5.3 (3.9) |  | Base case and SA |
| Ruhago et al (2015) ^23^ | Tanzania (TZA) | LIC | Diarrheal | district hospitals | Not reported | 4 | (2-6) | No, no sample size |
| Soltani et al (2015) ^24^ | Tunisia (TUN) | LMIC | Rotavirus diarrheal | 11 sentinel hospitals | 65 | 6 | 4  Min 1, max 45 | No, no SD |
| Alkoshi et al (2015) ^25^ | Libya (LBY) | LIC | Rotavirus diarrheal | 3 (referral?) hospitals | 239 |  | 3  Min 1, max 15 | Base case |
| Phavichitr et al (2013) ^26^ | Thailand (THA) | UMIC | Diarrheal | a tertiary hospital | Control arm: 53 |  | 3 [2-4] | Base case |
| Alvis-Guzman et al (2013) ^27^ | Colombia (COL) | UMIC | Diarrheal | primary, secondary, and tertiary | 1452 |  | 1 [1-1] | Base case |
| Sowmyanarayanan (2012) ^28^ | India (IND) | LMIC | Diarrheal | 10 hospitals, including general /governmental hospitals and a private hospital | 211 |  | 3 [2-5] | Base case |
| Osano et al (2011) ^29^ | Kenya (KEN) | LMIC | Rotavirus diarrheal | Kenyatta National Hospital | 172 | To going home  5.9 (7.5) | To going home  4 [2-7] | Base case and SA |
| Latipov et al (2011) ^30^ | Kazakhstan (KAZ) | UMIC | Rotavirus diarrheal | sentinel hospitals | unknow | 5.4 |  | No, no sample size |
| Flem et al (2009) ^31^ | Kyrgyzstan (KGZ) | LMIC | Rotavirus diarrheal | 2 public hospitals | 175 | 5.6 |  | No, no SD |
| Riewpaiboon et al (2016) ^37^ | Vietnam (VTM) | LMIC | Rotavirus diarrheal | multiple hospitals and outpatient clinics | 557 | 5.68  (SE:0.18) | 5 | Base case |
| Mendelsohn et al (2008) ^32^ | India (IND) | LMIC | Rotavirus diarrheal | a tertiary hospital and a community hospital | tertiary: 114  community: 18 |  | tertiary hospital: 2 [1.75-4]  community hospital: 3 [1-3] | Base case (tertiary hospital only *) |

* community hospital median removed, as the 3^rd^ quantile range was same as the median. ^#^ Burnett et al 2020 study is included in the LoS estimation, but excluded in comparison (section 2.3), because the inpatient costs were estimated using CHOICE-estimate.

Abbreviations: SE: standard error, SD: standard deviation, LIC: low-income countries, LMIC: lower-middle income countries, UMIC: upper-middle income countries, SA: sensitivity analysis.

Table 2.4: Length of stay of RSV childhood disease in low- and middle-income countries

| Reference | Country (ISO3) | Income group | Disease | Health facility | Sample size | Mean LoS in days (SD) [95% CI] | Median LoS [Interquartile range range] | Included in meta-analysis |
| --- | --- | --- | --- | --- | --- | --- | --- | --- |
| Tan et al (2023)^44^ | Thailand (THA) | UMIC | RSV | A tertiary hospital | 683 |  | 6 [4-9] | Base case |
| Ren et al (2023) ^46^ | China (CHN) | UMIC | RSV | A tertiary hospital | 261 |  | 6 [5-7] |  |
| Moyes et al (2023) ^45^ | South Africa (ZAF) | UMIC | RSV-severe cases | 5 sentinel hospitals | Severe cases: 675 ^§^ | 0-2m: 6.24  3-5m: 4.74  6-8m: 4.37  9-11m: 5.6  12-59m: 4.13 |  | No, no SD |
| Tian et al (2023) ^48^ | China (CHN) | UMIC | bronchiolitis | 27 tertiary hospitals | 42,928 |  | 6 [5-8] | Base case |
| Sun et al (2022) ^49^ | China (CHN) | UMIC | RSV-ARTI | A tertiary hospital | 7,248 |  | 8 [7-10] | Base case |
| Do et al (2022) ^50^ | Vietnam (VNM) | LMIC | RSV-LRTI | A tertiary hospital | 84 |  | 7 [6-8] | Base case |
| Buendia et al (2021) ^51^ | Colombia (COL) | UMIC | RSV-bronchiolitis | Tertiary hospitals | 193 |  | 5.88 [0.39] | Base case |
| Rodriguez-Martinez (2021) ^52^ | Colombia (COL) | UMIC | bronchiolitis | A tertiary hospital | 89 | Overall: 7.0 (4.4)  PW: 4.4 (3.2)  PIMC: 9.0 (4.1)  PICU: 10.3 (3.8) |  | Base case using 4.4 (3.2) |
| Baral et al (2020) ^55^ | Malawi (MWI) | LIC | RSV-ARTI | A tertiary hospital | 78 |  | 3 [2-4] | Base case |
| Marcone et al (2015) ^57^ | Argentina (ARG) | UMIC | Viral respiratory infection | Hospital records from claim database | 89 |  | 3[2-4] | Base case |
| Zhang et al (2013)^58^ | China (CHN) | UMIC | RSV-ARTI | A tertiary hospital | 2,721 |  | 8 [7-9] | Base case |
| Chan et al (2003)^59^ | Malaysia (MYS) | UMIC | RSV-bronchiolitis | A tertiary hospital | 176 term infants | 3.9 (SE: 0.2, 95% CI 3.7-4.3) |  | Base case |

^§^ imputed sample size by age-group is: 0-2m 226, 3-5m: 142, 6-8m: 85, 9-11m: 39, 12-59m: 183

Abbreviation: ARTI: acute respiratory tract infection, LRTI: lower respiratory tract infection, PW: pediatric ward, PIMC: pediatric intermediate medical care, PICU: pediatric intensive care unit, SE: standard error, SD: standard deviation, LIC: low-income countries, LMIC: lower-middle income countries, UMIC: upper-middle income countries.

Figure 2.1: Pooled average diarrheal disease length of hospital stay (days) based on mean and median converted to mean.

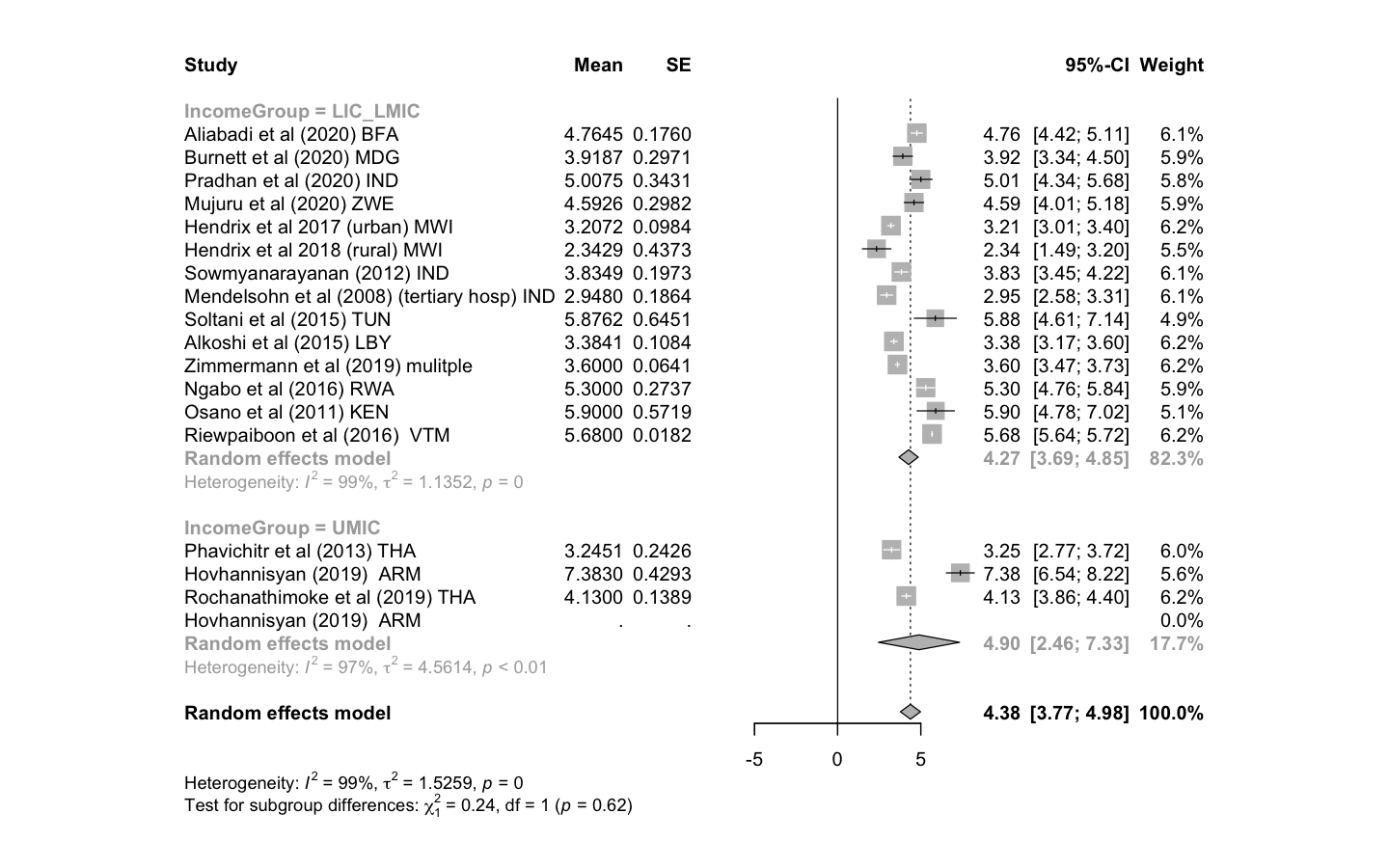

Figure 2.2: Pooled average diarrheal disease hospital length of hospital stay (days) based on mean only.

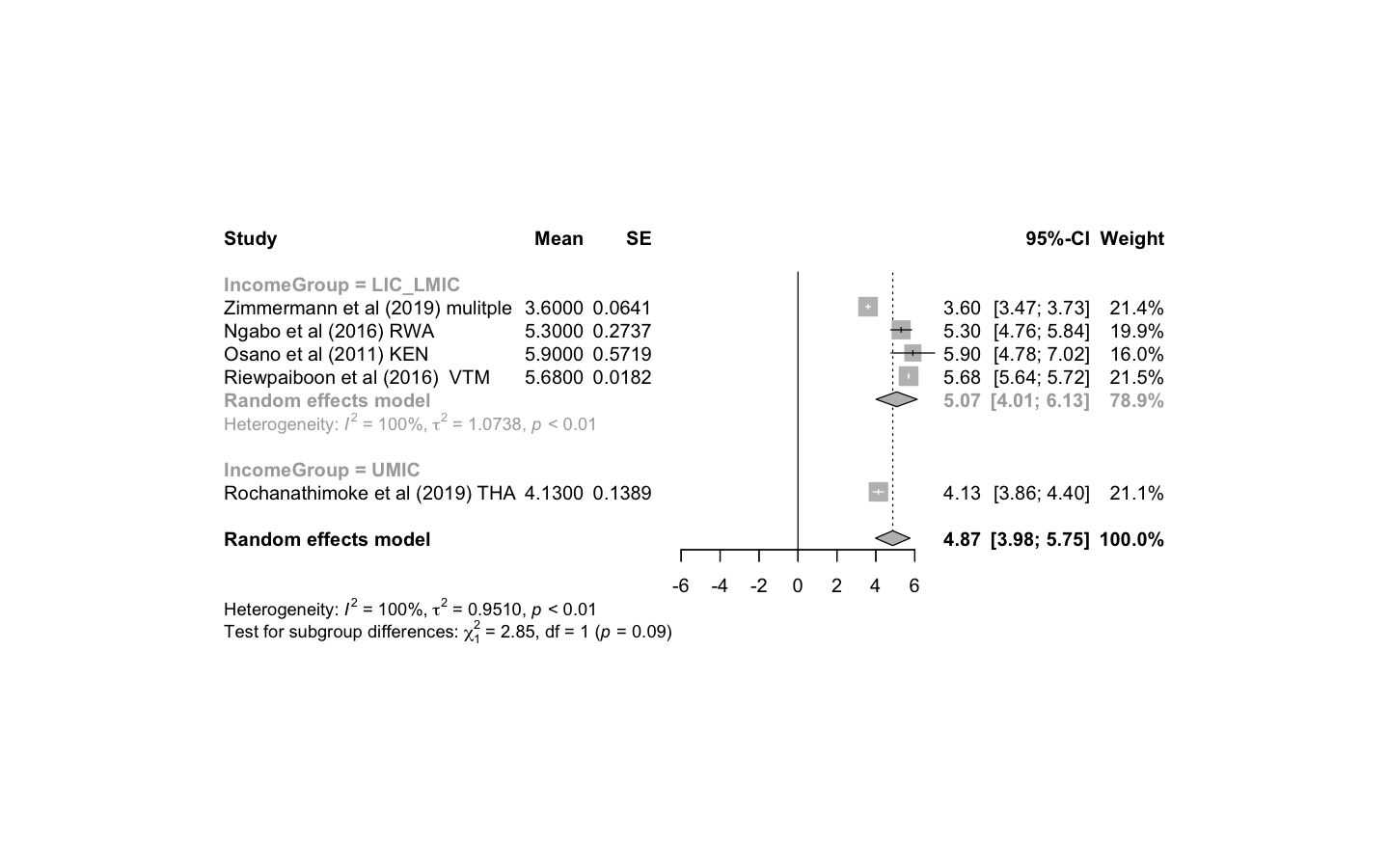

*Figure 2.3: Pooled average rotavirus-specific diarrheal disease length of hospital stay (days) based on mean and median converted to mean.*

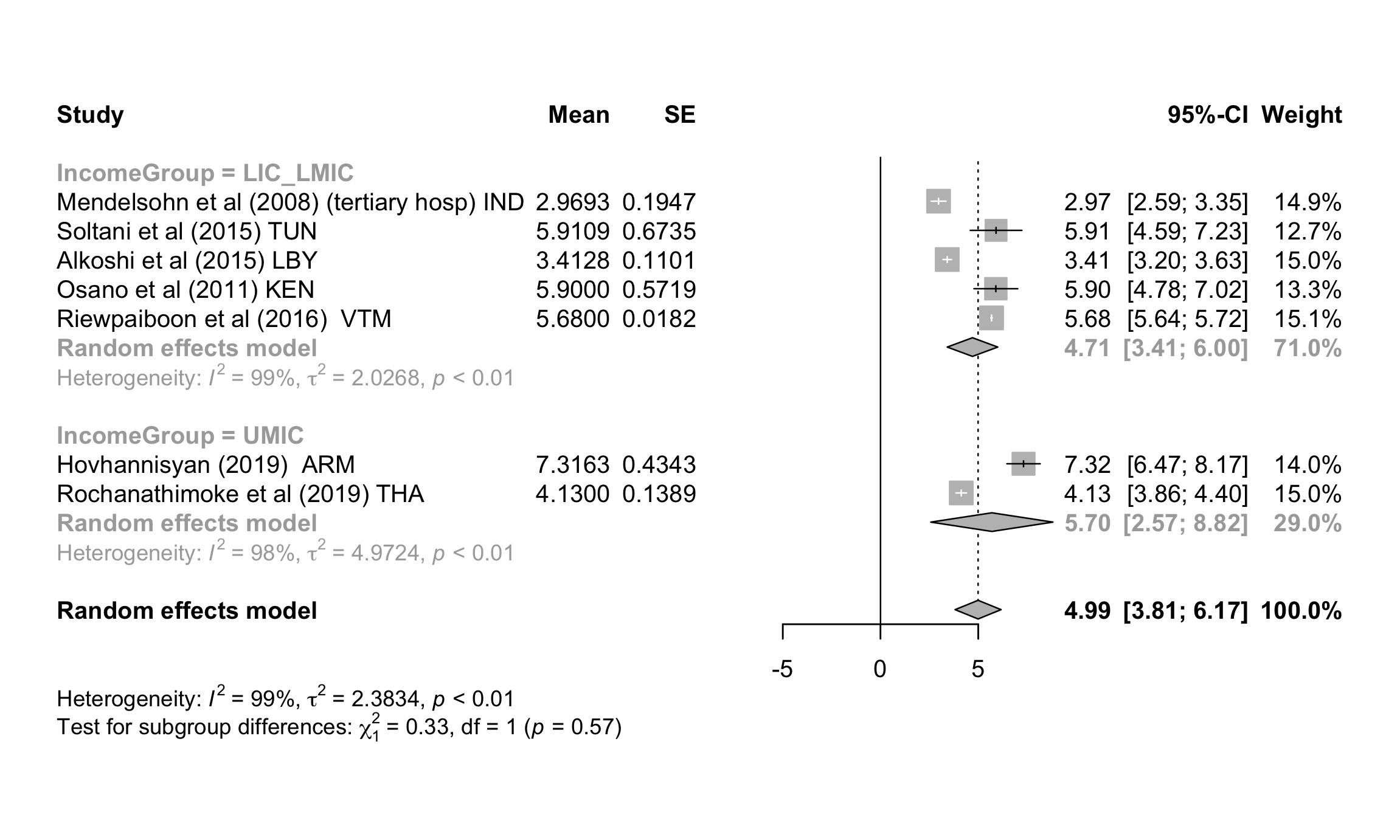

Figure 2.4: Pooled average RSV-related disease hospital length of hospital stay (days) based on mean and median converted to mean.
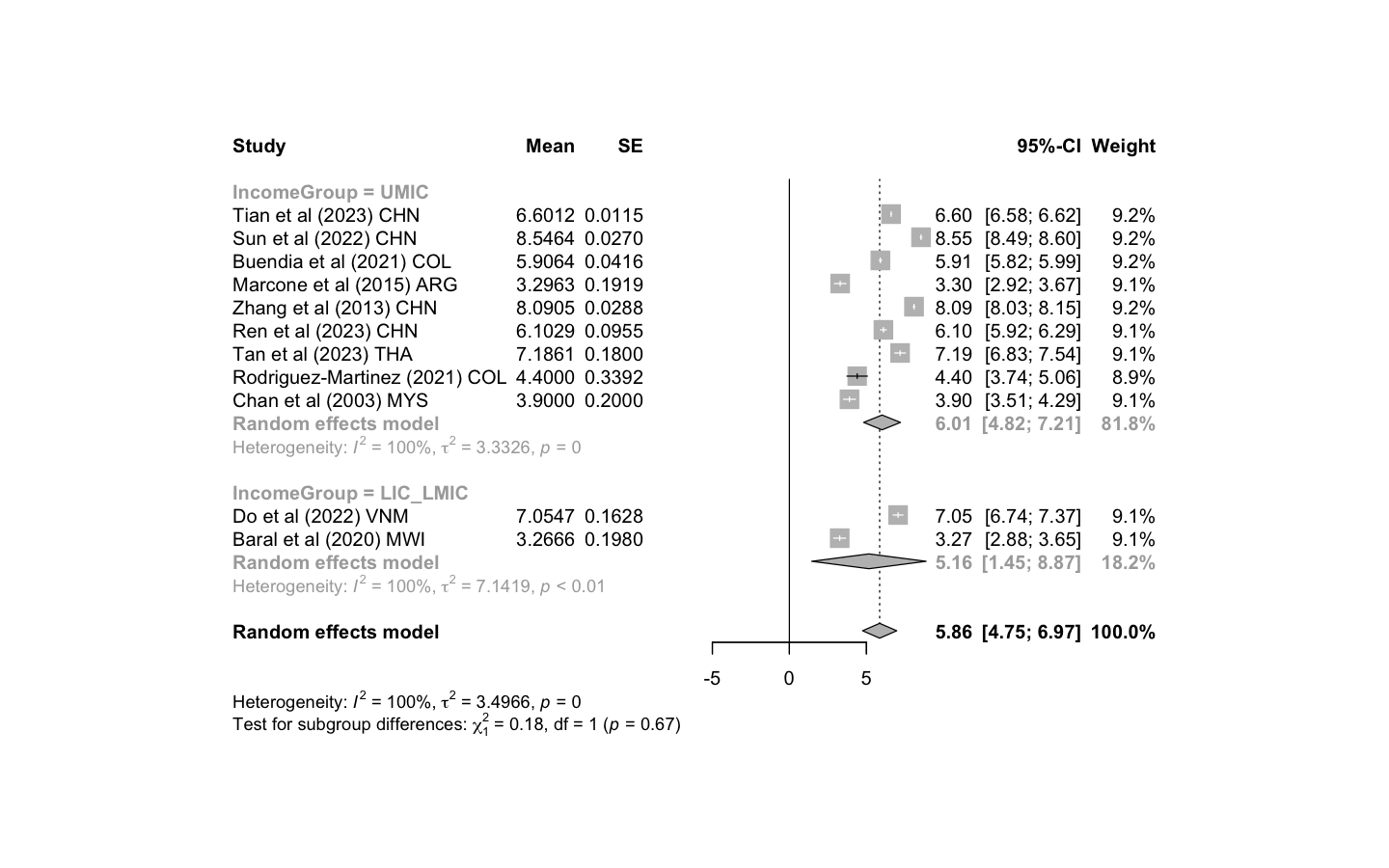

#### 2.3 Comparing the literature with the WHO-CHOICE estimates: subgroup analysis

We also performed a subgroup analysis, comparing rotavirus-specific diarrheal disease medical costs with WHO-CHOICE estimates per episode. The WHO-CHOICE inpatient cost per episode was estimated by multiplying the rotavirus-specific LoS (Figure 2.3) by the WHO-CHOICE unit cost per inpatient day. As presented in Figure 2.5, these findings are very similar to the comparison of non-rotavirus specific diarrheal disease versus WHO estimates (main text Figure 3)

Figure 2.5:Outpatient(left) and inpatient (right) direct medical costs for rotavirus-specific diarrheal disease. Red dots represent the WHO-CHOICE estimates, while blue dots represent mean costs reported in the literature (iUSD in 2022 value). Black bars represent the 95% uncertainty intervals.

| Outpatient | Inpatient |
| --- | --- |
| 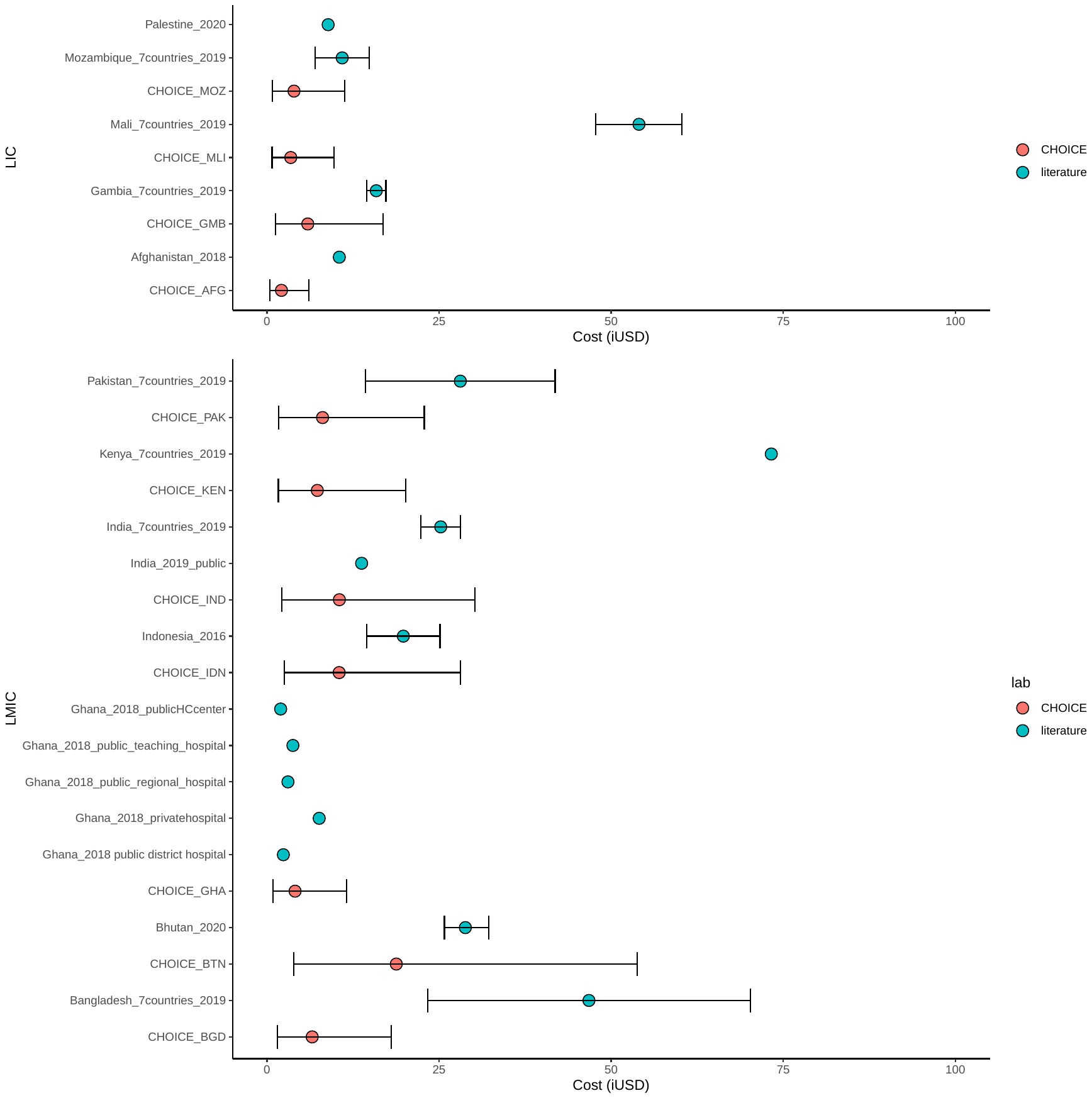 | 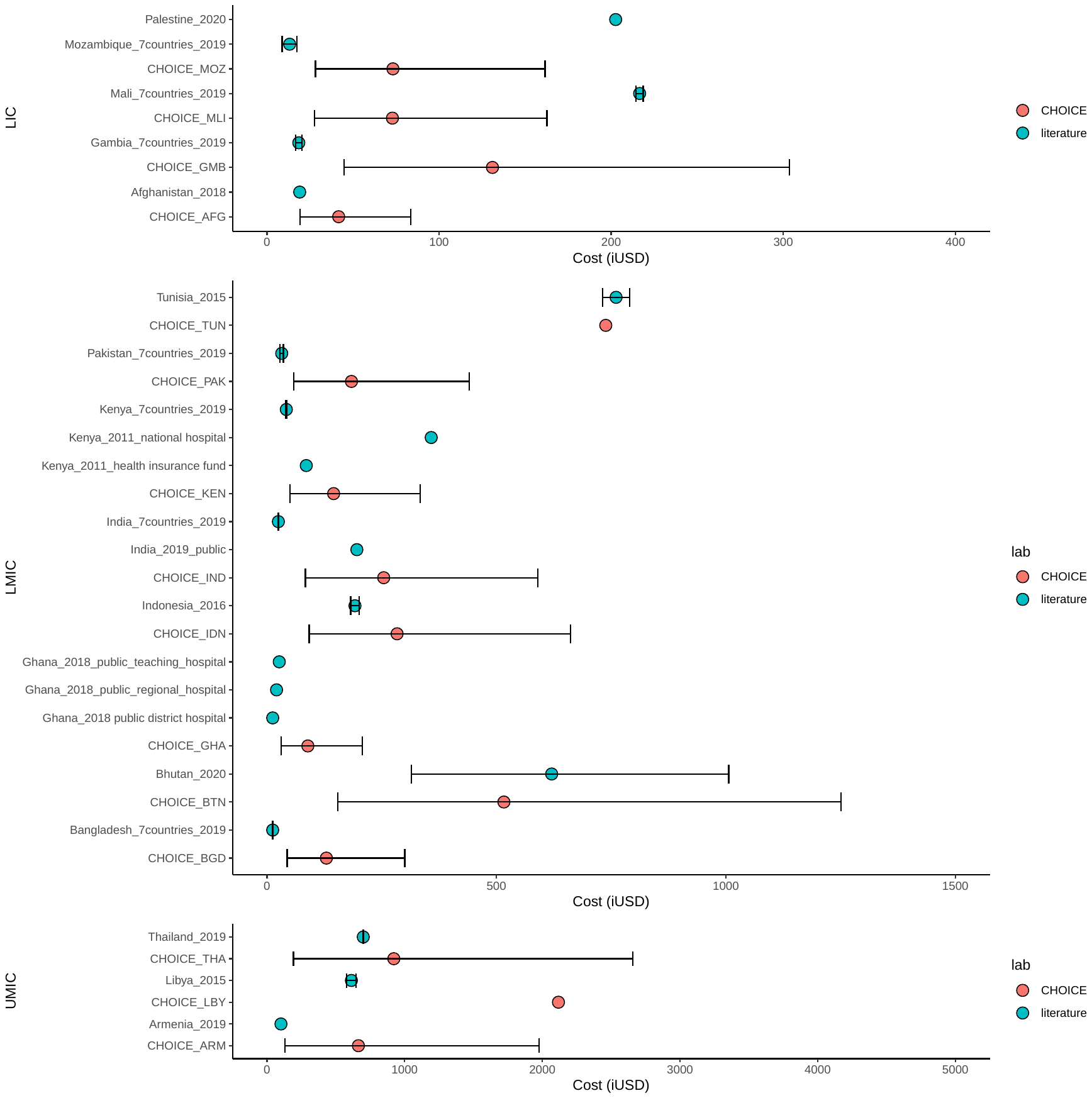 |

#### 2.4 Adjustment factors for RSV disease

As demonstrated in Figures 4 in the main text, the average outpatient and inpatient cost per RSV episode appeared to be underestimated, especially the outpatient episode, where the 95% UIs for the WHO-CHOICE estimates were not overlapping with the observed costs in LIC and LMIC countries. Hence, we performed a meta-analysis to estimate the adjustment factors using R package “metagen”.

For studies with multiple data points, the overall estimates were used; for example, data for 0-6-month and 6-59-month age groups were a subset of the overall data for children <5 years old, so we only used the data for the 0-59-month age group. If no estimates were presented for the overall age group (i.e., a South Africa study ^16^), estimates were pooled using inverse variance weighting method (IVW). For countries with multiple studies (i.e., China), IVW was also used to estimate the within-country cost per episode. The per-study adjustment factors were log-transformed, and the log mean of the meta-analyses are presented in Figure 2.6 and Figure 2.7.

The outpatient and inpatients adjustment ratios are presented in Table 2.5. For countries where no country-specific data are available for RSV disease, WHO-CHOICE outpatient and inpatient cost estimates can be multiplied by these adjustment factors to provide RSV-specific cost estimates.

Figure 2.6: Estimation of adjustment factors for RSV-related disease outpatient episodes. Meta-analysis of log mean and log standard deviation of the outpatient costs per episode from the literature divided by the country-specific WHO-CHOICE estimate.

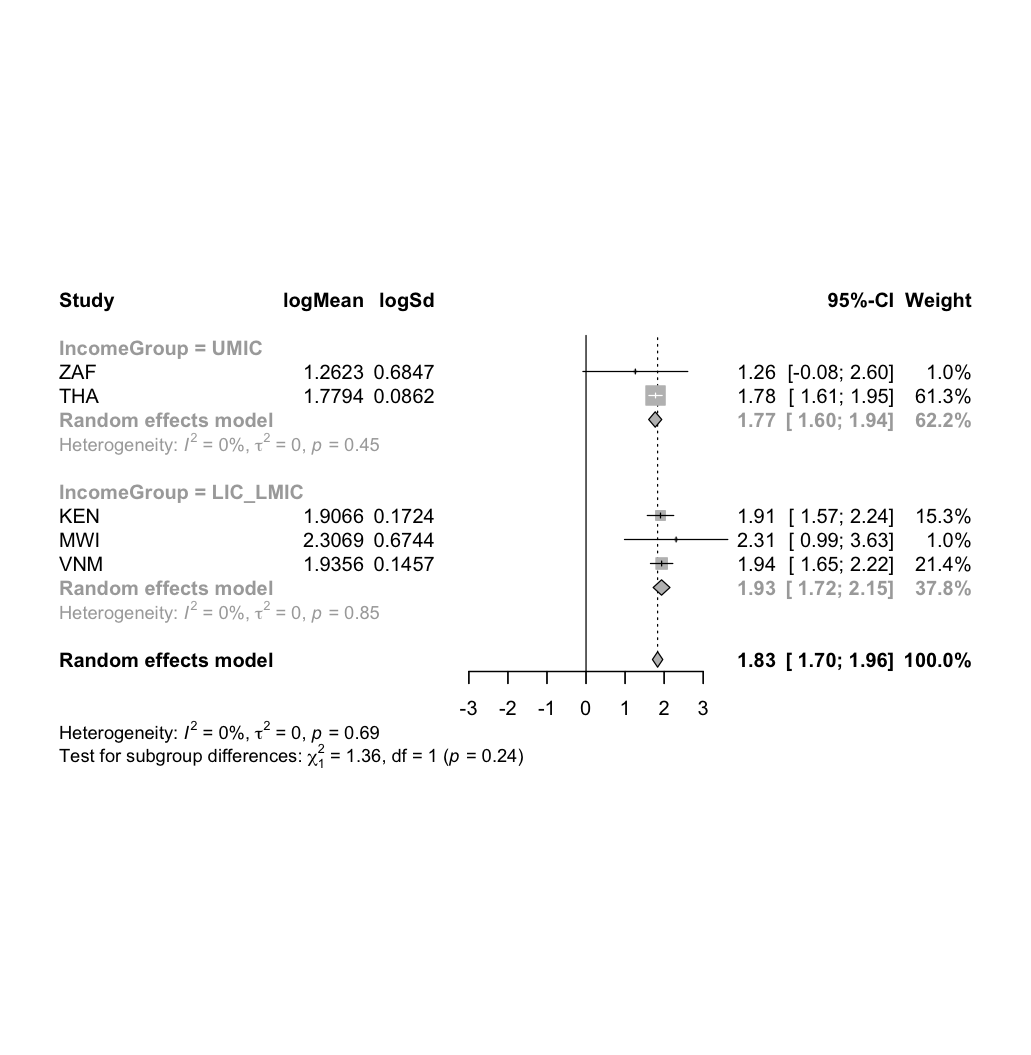

Figure 2.7: Estimation of adjustment factors for RSV disease inpatient episodes. Meta-analysis of log mean and log standard deviation of the inpatient costs per episode from the literature divided by the country-specific WHO-CHOICE estimate of inpatient cost per day times the estimated length of stay for RSV disease.

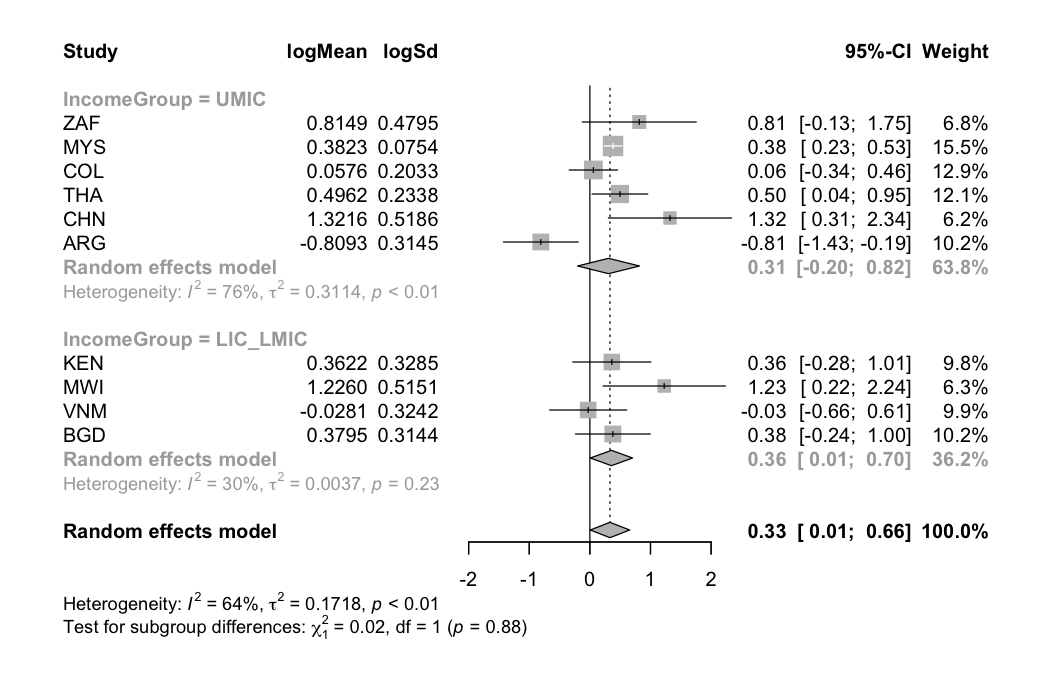

Table 2.5: Adjustment factors for RSV outpatient and inpatient costs per episode: mean and 95% credible interval

|  | LIC & LMIC | UMIC |
| --- | --- | --- |
| Outpatient | 6.89 (5.58-8.58) | 5.87 (4.95-6.96) |
| Inpatient | 1.43 (1.01-2.01) | 1.36 (0.82-2.27) |

#### 2.5 Country-specific direct medical cost and non-medical cost per outpatient and inpatient episode

Table 2.6 contains the estimated country-specific direct medical cost of diarrheal disease per outpatient and inpatient episode. For RSV disease, Table 2.7 and Table 2.8 report the country-specific direct medical costs with and without adjustment, respectively.

Table 2.6: Diarrheal disease direct medical cost per episode (iUSD in 2022 value).

| Country | ISO3 | Income Group | Source | Outpatient episode | | | Inpatient episode | | |
| --- | --- | --- | --- | --- | --- | --- | --- | --- | --- |
|  |  |  |  | Mean | Lower 95% uncertainty interval | Upper 95% uncertainty interval | Mean | Lower 95% uncertainty interval | Upper 95% uncertainty interval |
| Afghanistan | AFG | LIC | WHO-CHOICE | 2.11 | 0.43 | 6.08 | 37.16 | 20.23 | 67.11 |
| Afghanistan | AFG | LIC | Literature | 10.51 | 3.77 | 22.35 | 19.03 | 6.82 | 40.47 |
| Angola | AGO | LMIC | WHO-CHOICE | 16.22 | 3.42 | 46.18 | 412.45 | 140.11 | 934.66 |
| Albania | ALB | UMIC | WHO-CHOICE | 25.17 | 5.13 | 72.81 | 857.56 | 182.24 | 2452.48 |
| Argentina | ARG | UMIC | WHO-CHOICE | 46.03 | 10.99 | 123.73 | 1859.95 | 415.29 | 5176.71 |
| Armenia | ARM | UMIC | WHO-CHOICE | 18.46 | 4.12 | 51.27 | 565.13 | 118.25 | 1634.59 |
| Armenia | ARM | UMIC | Literature | NA | NA | NA | 101.77 | 36.48 | 216.41 |
| Azerbaijan | AZE | UMIC | WHO-CHOICE | 16.48 | 3.31 | 48.02 | 604.48 | 139.55 | 1666.12 |
| Burundi | BDI | LIC | WHO-CHOICE | 2.6 | 0.52 | 7.56 | 42.53 | 21.35 | 81.81 |
| Benin | BEN | LMIC | WHO-CHOICE | 4.71 | 1.06 | 13.04 | 89.01 | 39.02 | 178.66 |
| Burkina Faso | BFA | LIC | WHO-CHOICE | 4.66 | 0.93 | 13.63 | 79.65 | 34.9 | 161.15 |
| Burkina Faso | BFA | LIC | Literature | NA | NA | NA | 380.04 | 151.43 | 760.04 |
| Bangladesh | BGD | LMIC | WHO-CHOICE | 6.57 | 1.5 | 18.07 | 114.55 | 46.2 | 241.16 |
| Bangladesh | BGD | LMIC | Literature | 17.1 | 16.51 | 17.69 | 145.88 | 56.27 | 235.49 |
| Bulgaria | BGR | UMIC | WHO-CHOICE | 32.45 | 6.33 | 95.73 | 1286.04 | 278.78 | 3634.75 |
| Bosnia & Herzegovina | BIH | UMIC | WHO-CHOICE | 22.06 | 4.82 | 61.79 | 749.45 | 166.79 | 2098.93 |
| Belarus | BLR | UMIC | WHO-CHOICE | 23.03 | 4.89 | 65.41 | 910.75 | 209.78 | 2505.15 |
| Belize | BLZ | UMIC | WHO-CHOICE | 15.49 | 3.14 | 44.88 | 511.58 | 111.05 | 1455.77 |
| Bolivia | BOL | LMIC | WHO-CHOICE | 16.72 | 3.58 | 47.32 | 390.71 | 135.18 | 876.43 |
| Bolivia | BOL | LMIC | Literature: pooled | 57.24 | 30.96 | 83.52 | 244.5 | 61.93 | 427.06 |
| Brazil | BRA | UMIC | WHO-CHOICE | 5.13 | 1.11 | 14.49 | 193.83 | 48.68 | 521.85 |
| Brazil | BRA | UMIC | Literature | 11.45 | 4.1 | 24.35 | 371.92 | 133.31 | 790.86 |
| Bhutan | BTN | LMIC | WHO-CHOICE | 18.79 | 3.92 | 53.8 | 455.58 | 161.8 | 1003.76 |
| Bhutan | BTN | LMIC | Literature | 28.82 | 25.76 | 31.96 | 620.38 | 347.72 | 995.12 |
| Botswana | BWA | UMIC | WHO-CHOICE | 31.28 | 7.13 | 85.95 | 1303.5 | 269.43 | 3766.28 |
| Central African Republic | CAF | LIC | WHO-CHOICE | 3.46 | 0.7 | 10.02 | 58.82 | 27.09 | 117.38 |
| China | CHN | UMIC | WHO-CHOICE | 16.95 | 3.71 | 47.49 | 555.35 | 125.57 | 1547.69 |
| Côte d'Ivoire | CIV | LMIC | WHO-CHOICE | 6.03 | 1.34 | 16.74 | 113.33 | 44.81 | 242.15 |
| Cameroon | CMR | LMIC | WHO-CHOICE | 7.38 | 1.55 | 21.07 | 144.47 | 55.89 | 309.77 |
| Congo - Kinshasa | COD | LIC | WHO-CHOICE | 0.78 | 0.16 | 2.24 | 21.62 | 14.7 | 33.37 |
| Congo - Brazzaville | COG | LMIC | WHO-CHOICE | 8.58 | 1.94 | 23.68 | 207.66 | 78.88 | 445.19 |
| Colombia | COL | UMIC | WHO-CHOICE | 46.37 | 10.17 | 129.76 | 905.85 | 201.58 | 2533.53 |
| Colombia | COL | UMIC | Literature | 46.37 | 10.17 | 129.76 | 805.06 | 545.5 | 1064.62 |
| Comoros | COM | LMIC | WHO-CHOICE | 4.39 | 0.76 | 13.58 | 73.08 | 31.08 | 152.53 |
| Cape Verde | CPV | LMIC | WHO-CHOICE | 10.99 | 2.47 | 30.41 | 252.16 | 93.87 | 545.06 |
| Costa Rica | CRI | UMIC | WHO-CHOICE | 31.11 | 6.88 | 86.71 | 1178.07 | 252.46 | 3349.07 |
| Djibouti | DJI | LMIC | WHO-CHOICE | 7.32 | 1.49 | 21.18 | 145.91 | 57.28 | 309.6 |
| Dominica | DMA | UMIC | WHO-CHOICE | 28.53 | 4.33 | 93.4 | 1069.87 | 236.7 | 2997.6 |
| Dominican Republic | DOM | UMIC | WHO-CHOICE | 22.62 | 4.6 | 65.49 | 777.36 | 163.69 | 2234.51 |
| Algeria | DZA | LMIC | WHO-CHOICE | 16.58 | 3.58 | 46.7 | 470.23 | 153.01 | 1090.85 |
| Ecuador | ECU | UMIC | WHO-CHOICE | 22.82 | 5.04 | 63.66 | 811.63 | 175.1 | 2305.63 |
| Egypt | EGY | LMIC | WHO-CHOICE | 16.27 | 3.2 | 47.82 | 398.51 | 141.11 | 881.2 |
| Eritrea | ERI | LIC | WHO-CHOICE | 1.99 | 0.45 | 5.51 | 35.67 | 18.9 | 67.01 |
| Ethiopia | ETH | LIC | WHO-CHOICE | 3.17 | 0.7 | 8.85 | 58.15 | 26.34 | 117.95 |
| Ethiopia | ETH | LIC | Literature | 18.19 | 0.78 | 81.56 | 231.08 | 36.19 | 723.32 |
| Fiji | FJI | UMIC | WHO-CHOICE | 14.4 | 3.05 | 40.98 | 435.47 | 93.07 | 1251.37 |
| Micronesia (Federated States of) | FSM | LMIC | WHO-CHOICE | 9.43 | 1.96 | 27.03 | 202.21 | 77.31 | 432 |
| Gabon | GAB | UMIC | WHO-CHOICE | 31.17 | 7.11 | 85.65 | 1279.61 | 280.15 | 3599.83 |
| Georgia | GEO | UMIC | WHO-CHOICE | 15.16 | 3.01 | 44.37 | 457.68 | 103.06 | 1281.1 |
| Ghana | GHA | LMIC | WHO-CHOICE | 4.08 | 0.87 | 11.6 | 78.83 | 32.64 | 166.94 |
| Ghana | GHA | LMIC | Literature | 4.18 | 1.92 | 6.44 | 26.84 | 13.42 | 40.26 |
| Guinea | GIN | LIC | WHO-CHOICE | 5.6 | 1.22 | 15.7 | 87.99 | 37.63 | 180.14 |
| Gambia | GMB | LIC | WHO-CHOICE | 5.93 | 1.25 | 16.9 | 115.93 | 46.84 | 243.58 |
| Gambia | GMB | LIC | Literature | 15.88 | 3.73 | 41.95 | 18.51 | 1.29 | 74.58 |
| Guinea-Bissau | GNB | LIC | WHO-CHOICE | 3.7 | 0.78 | 10.55 | 67.16 | 30.58 | 133.53 |
| Equatorial Guinea | GNQ | UMIC | WHO-CHOICE | 70.27 | 14.15 | 204.41 | 3468.49 | 713.4 | 10012.77 |
| Grenada | GRD | UMIC | WHO-CHOICE | 25.4 | 5.86 | 69.43 | 931.18 | 208.47 | 2596.44 |
| Guatemala | GTM | UMIC | WHO-CHOICE | 15.85 | 3.35 | 45.11 | 441.95 | 101.98 | 1222.74 |
| Guyana | GUY | UMIC | WHO-CHOICE | 8.53 | 1.81 | 24.23 | 227.43 | 52.66 | 636.64 |
| Honduras | HND | LMIC | WHO-CHOICE | 13.03 | 2.73 | 37.25 | 292.07 | 105.71 | 640.6 |
| Haiti | HTI | LMIC | WHO-CHOICE | 3.89 | 0.75 | 11.55 | 69.08 | 30.55 | 140.37 |
| Indonesia | IDN | LMIC | WHO-CHOICE | 10.49 | 2.52 | 28.11 | 250.69 | 96.4 | 530.75 |
| Indonesia | IDN | LMIC | Literature | 19.81 | 15.06 | 25.21 | 191.79 | 182.61 | 200.79 |
| India | IND | LMIC | WHO-CHOICE | 10.52 | 2.18 | 30.24 | 225.01 | 87.65 | 473.54 |
| India | IND | LMIC | Literature, pooled | 26.25 | 8.81 | 43.69 | 277.43 | 69.19 | 485.67 |
| Iran | IRN | LMIC | WHO-CHOICE | 18.75 | 3.68 | 55.17 | 601.99 | 203.99 | 1359.65 |
| Iraq | IRQ | UMIC | WHO-CHOICE | 6.09 | 1.34 | 17.01 | 176.12 | 42.75 | 484.76 |
| Jamaica | JAM | UMIC | WHO-CHOICE | 20.02 | 3.92 | 58.93 | 681.53 | 149.78 | 1921.88 |
| Jordan | JOR | UMIC | WHO-CHOICE | 16.82 | 3.8 | 46.43 | 513.25 | 116.8 | 1427.16 |
| Kazakhstan | KAZ | UMIC | WHO-CHOICE | 32.89 | 6.56 | 96.08 | 1258.55 | 283.05 | 3495.94 |
| Kazakhstan | KAZ | UMIC | Literature | 55.84 | 20.02 | 118.74 | 562.44 | 201.6 | 1195.98 |
| Kenya | KEN | LMIC | WHO-CHOICE | 7.32 | 1.65 | 20.19 | 128.59 | 52.11 | 268.28 |
| Kenya | KEN | LMIC | Literature | 73.26 | 24.7 | 161.19 | 357.92 | 178.96 | 536.88 |
| Kyrgyzstan | KGZ | LMIC | WHO-CHOICE | 6.89 | 1.55 | 19.07 | 138.72 | 52.71 | 301.68 |
| Kyrgyzstan | KGZ | LMIC | Literature | 6.89 | 1.55 | 19.07 | 208.46 | 74.72 | 443.27 |
| Cambodia | KHM | LMIC | WHO-CHOICE | 7.53 | 1.52 | 21.87 | 149.48 | 56.8 | 324.02 |
| Kiribati | KIR | LMIC | WHO-CHOICE | 7.76 | 1.56 | 22.63 | 147.97 | 57.43 | 316.28 |
| Laos | LAO | LMIC | WHO-CHOICE | 8.48 | 1.89 | 23.56 | 172.39 | 65.6 | 371.26 |
| Lebanon | LBN | LMIC | WHO-CHOICE | 36.81 | 7.9 | 104 | 1184.06 | 395.89 | 2681.87 |
| Liberia | LBR | LIC | WHO-CHOICE | 4.01 | 0.86 | 11.3 | 60.13 | 26.46 | 124.58 |
| Libya | LBY | UMIC | WHO-CHOICE | 43.3 | 9.97 | 118.44 | 1802.83 | 403.83 | 5010.52 |
| Libya | LBY | UMIC | Literature | NA | NA | NA | 613.21 | 106.14 | 1846.04 |
| St. Lucia | LCA | UMIC | WHO-CHOICE | 28.06 | 6.19 | 78.34 | 1029.62 | 233.29 | 2852.76 |
| Sri Lanka | LKA | LMIC | WHO-CHOICE | 14.34 | 3.36 | 38.91 | 360.1 | 126.58 | 801.09 |
| Lesotho | LSO | LMIC | WHO-CHOICE | 4.76 | 1.02 | 13.43 | 92.88 | 37.73 | 197.05 |
| Morocco | MAR | LMIC | WHO-CHOICE | 12.79 | 2.73 | 36.26 | 303.86 | 111.57 | 660 |
| Moldova | MDA | UMIC | WHO-CHOICE | 11.85 | 2.41 | 34.34 | 302.61 | 73.23 | 822.16 |
| Madagascar | MDG | LIC | WHO-CHOICE | 3.14 | 0.69 | 8.79 | 54.77 | 25.43 | 109.42 |
| Maldives | MDV | UMIC | WHO-CHOICE | 22.4 | 4.52 | 65.12 | 750.43 | 172.66 | 2068.05 |
| Mexico | MEX | UMIC | WHO-CHOICE | 36.78 | 8.23 | 101.98 | 1542.72 | 337.2 | 4339.92 |
| Marshall Islands | MHL | UMIC | WHO-CHOICE | 7.85 | 1.68 | 22.2 | 197.92 | 48.04 | 542.51 |
| North Macedonia | MKD | UMIC | WHO-CHOICE | 29.01 | 6.08 | 82.87 | 1063.48 | 234.01 | 2987.52 |
| Mali | MLI | LIC | WHO-CHOICE | 3.46 | 0.75 | 9.73 | 64.65 | 29.02 | 130.56 |
| Mali | MLI | LIC | literature | 54.04 | 8.33 | 170.2 | 216.55 | 152.7 | 292.83 |
| Myanmar (Burma) | MMR | LMIC | WHO-CHOICE | 3.08 | 0.68 | 8.63 | 59.37 | 27.12 | 119.2 |
| Montenegro | MNE | UMIC | WHO-CHOICE | 36.17 | 7.31 | 105.07 | 1455.29 | 306.87 | 4164.16 |
| Mongolia | MNG | LMIC | WHO-CHOICE | 10.14 | 2.14 | 28.87 | 233.36 | 85.28 | 511.34 |
| Mozambique | MOZ | LIC | WHO-CHOICE | 3.93 | 0.81 | 11.29 | 64.94 | 29.52 | 129.71 |
| Mozambique | MOZ | LIC | Literature | 10.93 | 0.13 | 57.3 | 13.18 | 0.07 | 70.74 |
| Mauritania | MRT | LMIC | WHO-CHOICE | 6.74 | 1.51 | 18.69 | 141.47 | 54.81 | 303.31 |
| Mauritius | MUS | UMIC | WHO-CHOICE | 40.6 | 9.3 | 111.3 | 1612.68 | 349 | 4557.18 |
| Malawi | MWI | LIC | WHO-CHOICE | 2.89 | 0.61 | 8.25 | 50.36 | 24.22 | 98.47 |
| Malawi | MWI | LIC | Literature: pooled | 4.19 | 3.91 | 4.47 | 27.55 | 13.73 | 41.37 |
| Malaysia | MYS | UMIC | WHO-CHOICE | 32.79 | 6.96 | 93.12 | 1349.84 | 301.2 | 3762.44 |
| Namibia | NAM | UMIC | WHO-CHOICE | 16.41 | 3.4 | 47.16 | 527.79 | 118.98 | 1473.96 |
| Niger | NER | LIC | WHO-CHOICE | 2.3 | 0.47 | 6.67 | 39.11 | 20.58 | 72.67 |
| Nigeria | NGA | LMIC | WHO-CHOICE | 11.05 | 2.32 | 31.51 | 219.13 | 79.06 | 484.97 |
| Nicaragua | NIC | LMIC | WHO-CHOICE | 10.94 | 2.48 | 30.15 | 246.84 | 90.15 | 540.32 |
| Nepal | NPL | LMIC | WHO-CHOICE | 4.3 | 0.95 | 12.02 | 79.45 | 33.55 | 165.55 |
| Pakistan | PAK | LMIC | WHO-CHOICE | 8.08 | 1.73 | 22.85 | 162.61 | 61.21 | 353.51 |
| Pakistan | PAK | LMIC | literature | 28.09 | 0.28 | 148.63 | 32.17 | 1.33 | 145.18 |
| Peru | PER | UMIC | WHO-CHOICE | 22.46 | 5.45 | 59.88 | 812.79 | 172.04 | 2329.77 |
| Philippines | PHL | LMIC | WHO-CHOICE | 12.22 | 2.7 | 34.09 | 278.37 | 98.51 | 619.84 |
| Palau | PLW | UMIC | WHO-CHOICE | 43.39 | 9.64 | 120.72 | 1881.64 | 392.8 | 5403.75 |
| Papua New Guinea | PNG | LMIC | WHO-CHOICE | 8.12 | 1.64 | 23.62 | 153.32 | 57.61 | 334.52 |
| Paraguay | PRY | UMIC | WHO-CHOICE | 16.41 | 3.55 | 46.2 | 507.85 | 114.49 | 1418.82 |
| Russia | RUS | UMIC | WHO-CHOICE | 40.82 | 8.12 | 119.4 | 1867.33 | 375.47 | 5455.09 |
| Rwanda | RWA | LIC | WHO-CHOICE | 4.37 | 0.98 | 12.15 | 78.63 | 33.38 | 163.28 |
| Rwanda | RWA | LIC | Literature | NA | NA | NA | 130.78 | 45.54 | 282.65 |
| Sudan | SDN | LIC | WHO-CHOICE | 16.24 | 3.32 | 46.89 | 310.97 | 104.48 | 713.06 |
| Senegal | SEN | LMIC | WHO-CHOICE | 5.88 | 1.23 | 16.81 | 109.14 | 44.51 | 228.51 |
| Solomon Islands | SLB | LMIC | WHO-CHOICE | 9.02 | 1.77 | 26.57 | 179.71 | 66.08 | 395.35 |
| Sierra Leone | SLE | LIC | WHO-CHOICE | 4.86 | 1.07 | 13.55 | 78.53 | 34.26 | 159.56 |
| El Salvador | SLV | LMIC | WHO-CHOICE | 18.15 | 4.06 | 50.3 | 466.05 | 169.58 | 1011.54 |
| Serbia | SRB | UMIC | WHO-CHOICE | 28.93 | 6.5 | 80.05 | 1097.04 | 241.69 | 3079.48 |
| Sao Tome and Principe | STP | LMIC | WHO-CHOICE | 7.6 | 1.69 | 21.17 | 142.41 | 55.46 | 304.16 |
| Suriname | SUR | UMIC | WHO-CHOICE | 32.61 | 6.88 | 92.91 | 1049.23 | 236.34 | 2915.01 |
| Eswatini | SWZ | LMIC | WHO-CHOICE | 12.46 | 2.2 | 38.37 | 303.57 | 109.51 | 666.71 |
| Syria | SYR | LIC | WHO-CHOICE | 17.09 | 3.4 | 49.96 | 409.63 | 143.72 | 910.53 |
| Chad | TCD | LIC | WHO-CHOICE | 4.3 | 0.96 | 11.92 | 78.56 | 33.61 | 162.16 |
| Togo | TGO | LIC | WHO-CHOICE | 3.65 | 0.79 | 10.26 | 62.03 | 29.61 | 119.31 |
| Thailand | THA | UMIC | WHO-CHOICE | 21.92 | 4.5 | 63.19 | 784.53 | 174.45 | 2197.32 |
| Thailand | THA | UMIC | Literature | NA | NA | NA | 623.01 | 465.69 | 780.34 |
| Tajikistan | TJK | LMIC | WHO-CHOICE | 4.34 | 1.01 | 11.84 | 623.01 | 465.69 | 780.34 |
| Turkmenistan | TKM | UMIC | WHO-CHOICE | 22.52 | 4.83 | 63.68 | 765.62 | 162.55 | 2192.53 |
| Timor-Leste | TLS | LMIC | WHO-CHOICE | 4.89 | 0.99 | 14.21 | 89.14 | 36.93 | 186.85 |
| Tonga | TON | UMIC | WHO-CHOICE | 15.52 | 3.29 | 44.12 | 427.11 | 96.4 | 1195.31 |
| Tunisia | TUN | LMIC | WHO-CHOICE | 22.07 | 4.37 | 64.71 | 650.06 | 214.2 | 1491.25 |
| Tunisia | TUN | LMIC | Literature | NA | NA | NA | 760.74 | 404.7 | 1265.21 |
| Turkey | TUR | UMIC | WHO-CHOICE | 31.58 | 6.84 | 88.87 | 1333.3 | 288.27 | 3772.38 |
| Tuvalu | TUV | UMIC | WHO-CHOICE | 8.42 | 1.92 | 23.16 | 217.45 | 51.38 | 603.04 |
| Tanzania | TZA | LMIC | WHO-CHOICE | 4.35 | 0.94 | 12.25 | 78.31 | 32.9 | 164.06 |
| Tanzania | TZA | LMIC | literature | 11.08 | 3.97 | 23.56 | 103.76 | 37.19 | 220.64 |
| Uganda | UGA | LIC | WHO-CHOICE | 4.21 | 0.86 | 12.15 | 76.05 | 32.67 | 156.88 |
| Ukraine | UKR | LMIC | WHO-CHOICE | 12.91 | 3.12 | 34.5 | 341 | 123.27 | 746.4 |
| Uzbekistan | UZB | LMIC | WHO-CHOICE | 5.64 | 1.19 | 16.05 | 124.88 | 48.53 | 268.75 |
| St. Vincent & Grenadines | VCT | UMIC | WHO-CHOICE | 27.69 | 5.95 | 78.2 | 1058.15 | 224.6 | 3023.54 |
| Vietnam | VNM | LMIC | WHO-CHOICE | 10.7 | 2.27 | 30.43 | 222.06 | 83.99 | 476.55 |
| Vanuatu | VUT | LMIC | WHO-CHOICE | 11.63 | 2.62 | 32.16 | 273.4 | 100.97 | 592.88 |
| Samoa | WSM | LMIC | WHO-CHOICE | 13.29 | 2.82 | 37.73 | 308.41 | 111.65 | 675.64 |
| Yemen | YEM | LIC | WHO-CHOICE | 8.01 | 1.81 | 22.09 | 163.06 | 63.04 | 348.12 |
| South Africa | ZAF | UMIC | WHO-CHOICE | 25.19 | 5.63 | 69.83 | 913.69 | 199.84 | 2576.47 |
| Zambia | ZMB | LIC | WHO-CHOICE | 4.31 | 0.99 | 11.78 | 82.49 | 34.59 | 172.3 |

*NA: not available*

Table 2.7: RSV disease direct medical cost per outpatient and inpatient episode with adjustment factors applied (iUSD in 2022 value).

| Country | ISO3 | Income Group | Source | Outpatient episode | | | Inpatient episode | | |
| --- | --- | --- | --- | --- | --- | --- | --- | --- | --- |
|  |  |  |  | Mean | Lower 95% uncertainty interval | Upper 95% uncertainty interval | Mean | Lower 95% uncertainty interval | Upper 95% uncertainty interval |
| Afghanistan | AFG | LIC | WHO-CHOICE | 13.93 | 2.86 | 40.17 | 43.76 | 13.99 | 101.14 |
| Angola | AGO | LMIC | WHO-CHOICE | 107.6 | 22.71 | 306.41 | 759.31 | 225.4 | 1826.91 |
| Albania | ALB | UMIC | WHO-CHOICE | 143.12 | 29.17 | 414.09 | 1014.45 | 296.83 | 2459.56 |
| Argentina | ARG | UMIC | WHO-CHOICE | 262.6 | 62.68 | 705.9 | 2255.87 | 705.32 | 5277.24 |
| Argentina | ARG | UMIC | Literature: pooled | NA | NA | NA | 991.82 | 925.88 | 1057.76 |
| Armenia | ARM | UMIC | WHO-CHOICE | 114.91 | 25.62 | 319.09 | 876.28 | 247.77 | 2163.27 |
| Azerbaijan | AZE | UMIC | WHO-CHOICE | 85.05 | 17.06 | 247.83 | 484.39 | 151.96 | 1131.06 |
| Burundi | BDI | LIC | WHO-CHOICE | 19.32 | 3.9 | 56.12 | 65.19 | 19.27 | 157.2 |
| Benin | BEN | LMIC | WHO-CHOICE | 27.34 | 6.14 | 75.68 | 112.89 | 37.34 | 256 |
| Burkina Faso | BFA | LIC | WHO-CHOICE | 32.31 | 6.44 | 94.43 | 134.16 | 42.88 | 310.06 |
| Bangladesh | BGD | LMIC | WHO-CHOICE | 48.6 | 11.06 | 133.63 | 228.05 | 71.3 | 533.51 |
| Bangladesh | BGD | LMIC | Literature | NA | NA | NA | 253.94 | 231.55 | 276.32 |
| Bulgaria | BGR | UMIC | WHO-CHOICE | 191.35 | 37.33 | 564.48 | 1728.83 | 521.77 | 4122.74 |
| Bosnia & Herzegovina | BIH | UMIC | WHO-CHOICE | 136.81 | 29.93 | 383.29 | 1163.93 | 354.98 | 2759.95 |
| Belarus | BLR | UMIC | WHO-CHOICE | 136.26 | 28.93 | 386.99 | 1234.45 | 392.12 | 2862.79 |
| Belize | BLZ | UMIC | WHO-CHOICE | 86.79 | 17.62 | 251.54 | 567.26 | 165.64 | 1376.85 |
| Bolivia | BOL | LMIC | WHO-CHOICE | 116.34 | 24.9 | 329.22 | 775.84 | 234.26 | 1849.67 |
| Brazil | BRA | UMIC | WHO-CHOICE | 31.02 | 6.68 | 87.54 | 261.41 | 81.61 | 612.03 |
| Bhutan | BTN | LMIC | WHO-CHOICE | 121.76 | 25.39 | 348.62 | 808.3 | 253.41 | 1888.09 |
| Botswana | BWA | UMIC | WHO-CHOICE | 207.43 | 47.28 | 570.01 | 2440.54 | 703.17 | 5965.72 |
| Central African Republic | CAF | LIC | WHO-CHOICE | 24.74 | 5.03 | 71.63 | 96.36 | 29.55 | 227.84 |
| China | CHN | UMIC | WHO-CHOICE | 90.03 | 19.7 | 252.15 | 502.45 | 153.71 | 1189.45 |
| China | CHN | UMIC | Literature: pooled | NA | NA | NA | 1399.84 | 1261.39 | 1538.3 |
| Côte d'Ivoire | CIV | LMIC | WHO-CHOICE | 48.51 | 10.81 | 134.74 | 256.5 | 77.83 | 609.9 |
| Cameroon | CMR | LMIC | WHO-CHOICE | 44.49 | 9.33 | 127.06 | 210.78 | 65.27 | 495.68 |
| Congo - Kinshasa | COD | LIC | WHO-CHOICE | 5.51 | 1.15 | 15.77 | 15.99 | 4.78 | 38.31 |
| Congo - Brazzaville | COG | LMIC | WHO-CHOICE | 68.8 | 15.55 | 189.8 | 496.41 | 158.16 | 1149.3 |
| Colombia | COL | UMIC | WHO-CHOICE | 276.85 | 60.71 | 774.7 | 1260.67 | 386.92 | 2979.14 |
| Colombia | COL | UMIC | Literature: pooled | NA | NA | NA | 1802.7 | 1424.39 | 2181.01 |
| Comoros | COM | LMIC | WHO-CHOICE | 29.78 | 5.19 | 92.18 | 116.97 | 34.52 | 282.32 |
| Cape Verde | CPV | LMIC | WHO-CHOICE | 72.91 | 16.38 | 201.74 | 453.38 | 143.87 | 1052.04 |
| Costa Rica | CRI | UMIC | WHO-CHOICE | 205.59 | 45.48 | 572.93 | 2186.42 | 651.13 | 5251.42 |
| Djibouti | DJI | LMIC | WHO-CHOICE | 51.46 | 10.48 | 148.95 | 276.36 | 87.36 | 642.59 |
| Dominica | DMA | UMIC | WHO-CHOICE | 165.69 | 25.14 | 542.35 | 1366.54 | 418.95 | 3231.24 |
| Dominican Republic | DOM | UMIC | WHO-CHOICE | 129.05 | 26.26 | 373.61 | 927.03 | 267.8 | 2262.92 |
| Algeria | DZA | LMIC | WHO-CHOICE | 134.55 | 29.09 | 378.98 | 1193.42 | 340.44 | 2932.53 |
| Ecuador | ECU | UMIC | WHO-CHOICE | 136.15 | 30.05 | 379.83 | 1122.76 | 332.87 | 2703.14 |
| Egypt | EGY | LMIC | WHO-CHOICE | 106.29 | 20.91 | 312.39 | 713.6 | 221.36 | 1676.6 |
| Eritrea | ERI | LIC | WHO-CHOICE | 13.15 | 2.95 | 36.42 | 41.06 | 11.68 | 101.04 |
| Ethiopia | ETH | LIC | WHO-CHOICE | 19.74 | 4.35 | 55.08 | 75.5 | 22.31 | 182.12 |
| Fiji | FJI | UMIC | WHO-CHOICE | 96.13 | 20.33 | 273.51 | 807.6 | 229.58 | 1988.14 |
| Micronesia (Federated States of) | FSM | LMIC | WHO-CHOICE | 61.47 | 12.79 | 176.17 | 347.89 | 111.36 | 803.33 |
| Gabon | GAB | UMIC | WHO-CHOICE | 175.77 | 40.07 | 482.95 | 1481.82 | 451.53 | 3515.43 |
| Georgia | GEO | UMIC | WHO-CHOICE | 86.28 | 17.15 | 252.44 | 534.06 | 160.92 | 1274.7 |
| Ghana | GHA | LMIC | WHO-CHOICE | 24.53 | 5.2 | 69.72 | 104.17 | 30.33 | 253.24 |
| Guinea | GIN | LIC | WHO-CHOICE | 42.35 | 9.24 | 118.77 | 173.79 | 55.23 | 402.92 |
| Gambia | GMB | LIC | WHO-CHOICE | 47.64 | 10.04 | 135.75 | 262.32 | 82.44 | 611.95 |
| Guinea-Bissau | GNB | LIC | WHO-CHOICE | 25.1 | 5.29 | 71.54 | 104.94 | 33.53 | 242.58 |
| Equatorial Guinea | GNQ | UMIC | WHO-CHOICE | 382.1 | 76.93 | 1111.56 | 3531.06 | 1027.15 | 8587.87 |
| Grenada | GRD | UMIC | WHO-CHOICE | 143 | 32.97 | 390.83 | 1067.87 | 329.98 | 2514.24 |
| Guatemala | GTM | UMIC | WHO-CHOICE | 83.92 | 17.73 | 238.93 | 392.81 | 121.15 | 925.81 |
| Guyana | GUY | UMIC | WHO-CHOICE | 53.89 | 11.45 | 152.97 | 351.79 | 102.21 | 856.12 |
| Honduras | HND | LMIC | WHO-CHOICE | 88.46 | 18.52 | 252.85 | 549.93 | 171.1 | 1289.91 |
| Haiti | HTI | LMIC | WHO-CHOICE | 29.7 | 5.72 | 88.1 | 131.02 | 40.32 | 309.15 |
| Indonesia | IDN | LMIC | WHO-CHOICE | 81.59 | 19.61 | 218.6 | 579.62 | 190.9 | 1317.37 |
| India | IND | LMIC | WHO-CHOICE | 74.03 | 15.31 | 212.78 | 442.11 | 146.36 | 1001.93 |
| Iran | IRN | LMIC | WHO-CHOICE | 120.49 | 23.64 | 354.55 | 1061.81 | 320.36 | 2532.49 |
| Iraq | IRQ | UMIC | WHO-CHOICE | 40.33 | 8.88 | 112.65 | 302.67 | 89.29 | 730.67 |
| Jamaica | JAM | UMIC | WHO-CHOICE | 116.12 | 22.75 | 341.9 | 859.26 | 257.78 | 2055.69 |
| Jordan | JOR | UMIC | WHO-CHOICE | 97.66 | 22.06 | 269.54 | 644.18 | 197.55 | 1522.96 |
| Kazakhstan | KAZ | UMIC | WHO-CHOICE | 208.96 | 41.68 | 610.43 | 2112.37 | 660.62 | 4940.88 |
| Kenya | KEN | LMIC | WHO-CHOICE | 71.91 | 16.25 | 198.4 | 393 | 126.63 | 904.23 |
| Kenya | KEN | LMIC | Literature | 56.27 | 28.14 | 84.4 | 281.32 | 140.66 | 421.98 |
| Kyrgyzstan | KGZ | LMIC | WHO-CHOICE | 47.08 | 10.57 | 130.3 | 249.49 | 74.9 | 596.67 |
| Cambodia | KHM | LMIC | WHO-CHOICE | 39.64 | 8.01 | 115.12 | 169.29 | 51.52 | 401.88 |
| Kiribati | KIR | LMIC | WHO-CHOICE | 52.1 | 10.44 | 151.9 | 260.13 | 81.22 | 609 |
| Laos | LAO | LMIC | WHO-CHOICE | 59.12 | 13.17 | 164.23 | 327.16 | 102.03 | 766.41 |
| Lebanon | LBN | LMIC | WHO-CHOICE | 250.76 | 53.85 | 708.51 | 2328.59 | 706.42 | 5537.52 |
| Liberia | LBR | LIC | WHO-CHOICE | 30.54 | 6.58 | 86.17 | 109.79 | 31.3 | 269.85 |
| Libya | LBY | UMIC | WHO-CHOICE | 279.41 | 64.32 | 764.23 | 3167.75 | 993.09 | 7399.55 |
| St. Lucia | LCA | UMIC | WHO-CHOICE | 167.5 | 36.92 | 467.56 | 1436.36 | 450.37 | 3354.88 |
| Sri Lanka | LKA | LMIC | WHO-CHOICE | 100.99 | 23.64 | 274.03 | 726.89 | 222.22 | 1721.4 |
| Lesotho | LSO | LMIC | WHO-CHOICE | 31.61 | 6.79 | 89.27 | 150.77 | 45.07 | 361.36 |
| Morocco | MAR | LMIC | WHO-CHOICE | 85.75 | 18.3 | 243.01 | 561.27 | 178.07 | 1302.52 |
| Moldova | MDA | UMIC | WHO-CHOICE | 66.01 | 13.42 | 191.24 | 322.12 | 101.55 | 750.14 |
| Madagascar | MDG | LIC | WHO-CHOICE | 20.23 | 4.43 | 56.65 | 74 | 22.16 | 177.18 |
| Maldives | MDV | UMIC | WHO-CHOICE | 174.61 | 35.22 | 507.52 | 2027.06 | 639.22 | 4719.83 |
| Mexico | MEX | UMIC | WHO-CHOICE | 214.63 | 48 | 595.1 | 2011.15 | 614.1 | 4765.82 |
| Marshall Islands | MHL | UMIC | WHO-CHOICE | 37.74 | 8.08 | 106.77 | 102.45 | 30.82 | 244.74 |
| North Macedonia | MKD | UMIC | WHO-CHOICE | 166.97 | 34.99 | 477.03 | 1318.87 | 402.1 | 3127.9 |
| Mali | MLI | LIC | WHO-CHOICE | 19.5 | 4.23 | 54.86 | 72.38 | 22.2 | 171.1 |
| Myanmar (Burma) | MMR | LMIC | WHO-CHOICE | 21.38 | 4.68 | 59.85 | 92.87 | 28.19 | 220.77 |
| Montenegro | MNE | UMIC | WHO-CHOICE | 214.32 | 43.29 | 622.57 | 1987.22 | 585.25 | 4801.49 |
| Mongolia | MNG | LMIC | WHO-CHOICE | 61.7 | 13.03 | 175.68 | 360.07 | 110.96 | 849.01 |
| Mozambique | MOZ | LIC | WHO-CHOICE | 31.1 | 6.44 | 89.36 | 128.18 | 40.26 | 299.12 |
| Mauritania | MRT | LMIC | WHO-CHOICE | 41.42 | 9.27 | 114.8 | 212.87 | 65.8 | 501.12 |
| Mauritius | MUS | UMIC | WHO-CHOICE | 236.36 | 54.16 | 647.9 | 2085.87 | 631.27 | 4966.77 |
| Malawi | MWI | LIC | WHO-CHOICE | 21.12 | 4.43 | 60.3 | 80.92 | 24.68 | 191.88 |
| Malawi | MWI | LIC | Literature: pooled | 19.29 | 16.44 | 22.17 | 137.28 | 121.45 | 153.1 |
| Malaysia | MYS | UMIC | WHO-CHOICE | 208.28 | 44.21 | 591.6 | 2265.97 | 704.23 | 5318.35 |
| Malaysia | MYS | UMIC | Literature: pooled | NA | NA | NA | 903.63 | 714.69 | 1036.83 |
| Namibia | NAM | UMIC | WHO-CHOICE | 98.02 | 20.29 | 281.64 | 725.45 | 220.65 | 1722.78 |
| Niger | NER | LIC | WHO-CHOICE | 13.31 | 2.71 | 38.56 | 37.55 | 11.54 | 88.66 |
| Nigeria | NGA | LMIC | WHO-CHOICE | 74.54 | 15.68 | 212.62 | 401.84 | 121.1 | 959 |
| Nicaragua | NIC | LMIC | WHO-CHOICE | 83.83 | 19.01 | 230.95 | 557.71 | 172.73 | 1311.44 |
| Nepal | NPL | LMIC | WHO-CHOICE | 31.81 | 7 | 88.86 | 148.37 | 44.7 | 354.13 |
| Pakistan | PAK | LMIC | WHO-CHOICE | 54.46 | 11.66 | 154.07 | 290.74 | 88.66 | 689.47 |
| Peru | PER | UMIC | WHO-CHOICE | 128.34 | 31.16 | 342.14 | 975.59 | 283.78 | 2372.77 |
| Philippines | PHL | LMIC | WHO-CHOICE | 99.85 | 22.03 | 278.6 | 698.27 | 210.82 | 1664.83 |
| Palau | PLW | UMIC | WHO-CHOICE | 249.49 | 55.4 | 694.05 | 2334.51 | 683.94 | 5656.33 |
| Papua New Guinea | PNG | LMIC | WHO-CHOICE | 64.7 | 13.04 | 188.11 | 354 | 106.66 | 844.96 |
| Paraguay | PRY | UMIC | WHO-CHOICE | 91.59 | 19.81 | 257.9 | 556.17 | 168.82 | 1322.17 |
| Russia | RUS | UMIC | WHO-CHOICE | 206.34 | 41.04 | 603.54 | 1371.88 | 387.51 | 3388.55 |
| Rwanda | RWA | LIC | WHO-CHOICE | 28.47 | 6.35 | 79.05 | 119.16 | 36.08 | 283.67 |
| Sudan | SDN | LIC | WHO-CHOICE | 108.81 | 22.26 | 314.3 | 575.76 | 165.36 | 1409.77 |
| Senegal | SEN | LMIC | WHO-CHOICE | 41.22 | 8.63 | 117.83 | 198.19 | 62.24 | 462.53 |
| Solomon Islands | SLB | LMIC | WHO-CHOICE | 53.82 | 10.55 | 158.43 | 262.55 | 78.92 | 627.45 |
| Sierra Leone | SLE | LIC | WHO-CHOICE | 34.33 | 7.59 | 95.73 | 136.15 | 43.04 | 316.62 |
| El Salvador | SLV | LMIC | WHO-CHOICE | 125.71 | 28.14 | 348.39 | 923.49 | 297.63 | 2124.54 |
| Serbia | SRB | UMIC | WHO-CHOICE | 170.66 | 38.32 | 472.3 | 1475.17 | 450.66 | 3494.78 |
| Sao Tome and Principe | STP | LMIC | WHO-CHOICE | 58.86 | 13.04 | 163.89 | 312.45 | 97.34 | 732.37 |
| Suriname | SUR | UMIC | WHO-CHOICE | 174.3 | 36.76 | 496.61 | 990.42 | 308.91 | 2320.07 |
| Eswatini | SWZ | LMIC | WHO-CHOICE | 91.36 | 16.14 | 281.25 | 647.79 | 201.4 | 1520.08 |
| Syria | SYR | LIC | WHO-CHOICE | 121.64 | 24.22 | 355.59 | 845.09 | 259.89 | 1994.88 |
| Chad | TCD | LIC | WHO-CHOICE | 30.3 | 6.78 | 83.97 | 135.58 | 41.54 | 320.68 |
| Togo | TGO | LIC | WHO-CHOICE | 27.58 | 5.99 | 77.51 | 112.53 | 37.71 | 253.27 |
| Thailand | THA | UMIC | WHO-CHOICE | 143.42 | 29.47 | 413.44 | 1411.51 | 430.83 | 3345.6 |
| Thailand | THA | UMIC | Literature: pooled | 208.81 | 208.12 | 209.5 | 2777.43 | 2705.6 | 2849.25 |
| Tajikistan | TJK | LMIC | WHO-CHOICE | 31.14 | 7.21 | 84.93 | 157.73 | 50.09 | 365.85 |
| Turkmenistan | TKM | UMIC | WHO-CHOICE | 121.24 | 25.98 | 342.89 | 737.26 | 214.62 | 1792.38 |
| Timor-Leste | TLS | LMIC | WHO-CHOICE | 31.98 | 6.45 | 92.95 | 139.71 | 42.48 | 331.82 |
| Tonga | TON | UMIC | WHO-CHOICE | 95.38 | 20.22 | 271.09 | 634.11 | 190.66 | 1515.21 |
| Tunisia | TUN | LMIC | WHO-CHOICE | 153.56 | 30.38 | 450.19 | 1310.43 | 384.82 | 3171.09 |
| Turkey | TUR | UMIC | WHO-CHOICE | 162.52 | 35.21 | 457.29 | 1067.46 | 321.58 | 2548.07 |
| Tuvalu | TUV | UMIC | WHO-CHOICE | 48.67 | 11.08 | 133.81 | 256.48 | 75.8 | 618.55 |
| Tanzania | TZA | LMIC | WHO-CHOICE | 31.85 | 6.9 | 89.63 | 143.5 | 42.65 | 345 |
| Uganda | UGA | LIC | WHO-CHOICE | 26.56 | 5.45 | 76.61 | 108.37 | 33.02 | 257.07 |
| Ukraine | UKR | LMIC | WHO-CHOICE | 93.62 | 22.62 | 250.17 | 719.17 | 226.03 | 1677.58 |
| Uzbekistan | UZB | LMIC | WHO-CHOICE | 39.5 | 8.35 | 112.42 | 230.74 | 69.87 | 549.24 |
| St. Vincent & Grenadines | VCT | UMIC | WHO-CHOICE | 176.54 | 37.94 | 498.6 | 1782.94 | 524.5 | 4310.47 |
| Vietnam | VNM | LMIC | WHO-CHOICE | 70.13 | 14.85 | 199.4 | 387.91 | 123.86 | 897.04 |
| Vietnam | VNM | LMIC | Literature: pooled | 83.94 | 79.34 | 88.53 | 348.52 | 331.62 | 365.43 |
| Vanuatu | VUT | LMIC | WHO-CHOICE | 83.22 | 18.73 | 230.07 | 558.3 | 176.81 | 1296.88 |
| Samoa | WSM | LMIC | WHO-CHOICE | 87.37 | 18.56 | 248.04 | 552.24 | 172.6 | 1292.14 |
| Yemen | YEM | LIC | WHO-CHOICE | 60.83 | 13.77 | 167.71 | 351.98 | 111.06 | 819.27 |
| South Africa | ZAF | UMIC | WHO-CHOICE | 156.15 | 34.92 | 432.97 | 1421.84 | 429.06 | 3390.88 |
| South Africa | ZAF | UMIC | Literature: pooled | 59.9 | 57.13 | 62.67 | 1409.66 | 1294.31 | 1525.01 |
| Zambia | ZMB | LIC | WHO-CHOICE | 29.47 | 6.79 | 80.54 | 137.18 | 41.43 | 327.02 |

Table 2.8: RSV disease direct medical cost per outpatient and inpatient episode without adjustment factors (iUSD in 2022 value).

| Country | ISO3 | Income Group | Source | Outpatient episode | | | Inpatient episode | | |
| --- | --- | --- | --- | --- | --- | --- | --- | --- | --- |
|  |  |  |  | Mean | Lower 95% uncertainty interval | Upper 95% uncertainty interval | Mean | Lower 95% uncertainty interval | Upper 95% uncertainty interval |
| Afghanistan | AFG | LIC | WHO-CHOICE | 2.11 | 0.43 | 6.08 | 32.99 | 10.54 | 76.24 |
| Angola | AGO | LMIC | WHO-CHOICE | 16.22 | 3.42 | 46.18 | 568.37 | 168.72 | 1367.5 |
| Albania | ALB | UMIC | WHO-CHOICE | 25.17 | 5.13 | 72.81 | 837.63 | 245.09 | 2030.85 |
| Argentina | ARG | UMIC | WHO-CHOICE | 46.03 | 10.99 | 123.73 | 1842.24 | 576 | 4309.62 |
| Argentina | ARG | UMIC | Literature: pooled | NA | NA | NA | 991.82 | 925.88 | 1057.76 |
| Armenia | ARM | UMIC | WHO-CHOICE | 18.46 | 4.12 | 51.27 | 545.51 | 154.24 | 1346.69 |
| Azerbaijan | AZE | UMIC | WHO-CHOICE | 16.48 | 3.31 | 48.02 | 588.03 | 184.48 | 1373.07 |
| Burundi | BDI | LIC | WHO-CHOICE | 2.6 | 0.52 | 7.56 | 40.69 | 12.03 | 98.12 |
| Benin | BEN | LMIC | WHO-CHOICE | 4.71 | 1.06 | 13.04 | 106.84 | 35.33 | 242.28 |
| Burkina Faso | BFA | LIC | WHO-CHOICE | 4.66 | 0.93 | 13.63 | 93.55 | 29.9 | 216.21 |
| Bangladesh | BGD | LMIC | WHO-CHOICE | 6.57 | 1.5 | 18.07 | 143.32 | 44.81 | 335.3 |
| Bangladesh | BGD | LMIC | Literature | NA | NA | NA | 253.94 | 231.55 | 276.32 |
| Bulgaria | BGR | UMIC | WHO-CHOICE | 32.45 | 6.33 | 95.73 | 1266.32 | 382.18 | 3019.8 |
| Bosnia & Herzegovina | BIH | UMIC | WHO-CHOICE | 22.06 | 4.82 | 61.79 | 731.73 | 223.16 | 1735.1 |
| Belarus | BLR | UMIC | WHO-CHOICE | 23.03 | 4.89 | 65.41 | 894.71 | 284.2 | 2074.9 |
| Belize | BLZ | UMIC | WHO-CHOICE | 15.49 | 3.14 | 44.88 | 493.21 | 144.02 | 1197.11 |
| Bolivia | BOL | LMIC | WHO-CHOICE | 16.72 | 3.58 | 47.32 | 537.24 | 162.22 | 1280.83 |
| Brazil | BRA | UMIC | WHO-CHOICE | 5.13 | 1.11 | 14.49 | 177.64 | 55.46 | 415.9 |
| Bhutan | BTN | LMIC | WHO-CHOICE | 18.79 | 3.92 | 53.8 | 629.46 | 197.34 | 1470.34 |
| Botswana | BWA | UMIC | WHO-CHOICE | 31.28 | 7.13 | 85.95 | 1280.39 | 368.9 | 3129.82 |
| Central African Republic | CAF | LIC | WHO-CHOICE | 3.46 | 0.7 | 10.02 | 63.9 | 19.59 | 151.07 |
| China | CHN | UMIC | WHO-CHOICE | 16.95 | 3.71 | 47.49 | 538.17 | 164.64 | 1274 |
| China | CHN | UMIC | Literature: pooled | NA | NA | NA | 1399.84 | 1261.39 | 1538.3 |
| Côte d'Ivoire | CIV | LMIC | WHO-CHOICE | 6.03 | 1.34 | 16.74 | 141.64 | 42.98 | 336.78 |
| Cameroon | CMR | LMIC | WHO-CHOICE | 7.38 | 1.55 | 21.07 | 186 | 57.6 | 437.42 |
| Congo - Kinshasa | COD | LIC | WHO-CHOICE | 0.78 | 0.16 | 2.24 | 10.86 | 3.25 | 26.03 |
| Congo - Brazzaville | COG | LMIC | WHO-CHOICE | 8.58 | 1.94 | 23.68 | 275.99 | 87.93 | 638.97 |
| Colombia | COL | UMIC | WHO-CHOICE | 46.37 | 10.17 | 129.76 | 888.08 | 272.57 | 2098.64 |
| Colombia | COL | UMIC | Literature: pooled | NA | NA | NA | 1802.7 | 1424.39 | 2181.01 |
| Comoros | COM | LMIC | WHO-CHOICE | 4.39 | 0.76 | 13.58 | 84.27 | 24.87 | 203.39 |
| Cape Verde | CPV | LMIC | WHO-CHOICE | 10.99 | 2.47 | 30.41 | 339.43 | 107.71 | 787.62 |
| Costa Rica | CRI | UMIC | WHO-CHOICE | 31.11 | 6.88 | 86.71 | 1157.79 | 344.8 | 2780.83 |
| Djibouti | DJI | LMIC | WHO-CHOICE | 7.32 | 1.49 | 21.18 | 188.01 | 59.43 | 437.16 |
| Dominica | DMA | UMIC | WHO-CHOICE | 28.53 | 4.33 | 93.4 | 1051.71 | 322.43 | 2486.83 |
| Dominican Republic | DOM | UMIC | WHO-CHOICE | 22.62 | 4.6 | 65.49 | 757.26 | 218.75 | 1848.51 |
| Algeria | DZA | LMIC | WHO-CHOICE | 16.58 | 3.58 | 46.7 | 651.12 | 185.74 | 1599.96 |
| Ecuador | ECU | UMIC | WHO-CHOICE | 22.82 | 5.04 | 63.66 | 792.5 | 234.96 | 1908.01 |
| Egypt | EGY | LMIC | WHO-CHOICE | 16.27 | 3.2 | 47.82 | 548.17 | 170.04 | 1287.93 |
| Eritrea | ERI | LIC | WHO-CHOICE | 1.99 | 0.45 | 5.51 | 30.92 | 8.8 | 76.09 |
| Ethiopia | ETH | LIC | WHO-CHOICE | 3.17 | 0.7 | 8.85 | 62.98 | 18.6 | 151.91 |
| Fiji | FJI | UMIC | WHO-CHOICE | 14.4 | 3.05 | 40.98 | 416.83 | 118.49 | 1026.13 |
| Micronesia (Federated States of) | FSM | LMIC | WHO-CHOICE | 9.43 | 1.96 | 27.03 | 268.21 | 85.86 | 619.35 |
| Gabon | GAB | UMIC | WHO-CHOICE | 31.17 | 7.11 | 85.65 | 1260.59 | 384.12 | 2990.59 |
| Georgia | GEO | UMIC | WHO-CHOICE | 15.16 | 3.01 | 44.37 | 440.34 | 132.68 | 1051 |
| Ghana | GHA | LMIC | WHO-CHOICE | 4.08 | 0.87 | 11.6 | 92.49 | 26.93 | 224.84 |
| Guinea | GIN | LIC | WHO-CHOICE | 5.6 | 1.22 | 15.7 | 105.45 | 33.51 | 244.48 |
| Gambia | GMB | LIC | WHO-CHOICE | 5.93 | 1.25 | 16.9 | 145.28 | 45.66 | 338.91 |
| Guinea-Bissau | GNB | LIC | WHO-CHOICE | 3.7 | 0.78 | 10.55 | 75.76 | 24.21 | 175.11 |
| Equatorial Guinea | GNQ | UMIC | WHO-CHOICE | 70.27 | 14.15 | 204.41 | 3435.28 | 999.29 | 8354.92 |
| Grenada | GRD | UMIC | WHO-CHOICE | 25.4 | 5.86 | 69.43 | 913.71 | 282.34 | 2151.27 |
| Guatemala | GTM | UMIC | WHO-CHOICE | 15.85 | 3.35 | 45.11 | 425.22 | 131.14 | 1002.19 |
| Guyana | GUY | UMIC | WHO-CHOICE | 8.53 | 1.81 | 24.23 | 210.35 | 61.12 | 511.92 |
| Honduras | HND | LMIC | WHO-CHOICE | 13.03 | 2.73 | 37.25 | 396.41 | 123.34 | 929.82 |
| Haiti | HTI | LMIC | WHO-CHOICE | 3.89 | 0.75 | 11.55 | 78.53 | 24.17 | 185.29 |
| Indonesia | IDN | LMIC | WHO-CHOICE | 10.49 | 2.52 | 28.11 | 337.17 | 111.05 | 766.33 |
| India | IND | LMIC | WHO-CHOICE | 10.52 | 2.18 | 30.24 | 300.57 | 99.51 | 681.17 |
| Iran | IRN | LMIC | WHO-CHOICE | 18.75 | 3.68 | 55.17 | 838.57 | 253.01 | 2000.05 |
| Iraq | IRQ | UMIC | WHO-CHOICE | 6.09 | 1.34 | 17.01 | 159.43 | 47.03 | 384.88 |
| Jamaica | JAM | UMIC | WHO-CHOICE | 20.02 | 3.92 | 58.93 | 663.35 | 199.01 | 1587 |
| Jordan | JOR | UMIC | WHO-CHOICE | 16.82 | 3.8 | 46.43 | 496.23 | 152.18 | 1173.18 |
| Kazakhstan | KAZ | UMIC | WHO-CHOICE | 32.89 | 6.56 | 96.08 | 1241.41 | 388.24 | 2903.68 |
| Kenya | KEN | LMIC | WHO-CHOICE | 7.32 | 1.65 | 20.19 | 163.27 | 52.61 | 375.66 |
| Kenya | KEN | LMIC | Literature | 56.27 | 28.14 | 84.4 | 281.32 | 140.66 | 421.98 |
| Kyrgyzstan | KGZ | LMIC | WHO-CHOICE | 6.89 | 1.55 | 19.07 | 177.87 | 53.4 | 425.38 |
| Cambodia | KHM | LMIC | WHO-CHOICE | 7.53 | 1.52 | 21.87 | 193.19 | 58.8 | 458.63 |
| Kiribati | KIR | LMIC | WHO-CHOICE | 7.76 | 1.56 | 22.63 | 190.98 | 59.63 | 447.1 |
| Laos | LAO | LMIC | WHO-CHOICE | 8.48 | 1.89 | 23.56 | 225.79 | 70.42 | 528.94 |
| Lebanon | LBN | LMIC | WHO-CHOICE | 36.81 | 7.9 | 104 | 1668.6 | 506.2 | 3968.03 |
| Liberia | LBR | LIC | WHO-CHOICE | 4.01 | 0.86 | 11.3 | 65.82 | 18.77 | 161.78 |
| Libya | LBY | UMIC | WHO-CHOICE | 43.3 | 9.97 | 118.44 | 1785.44 | 559.74 | 4170.61 |
| St. Lucia | LCA | UMIC | WHO-CHOICE | 28.06 | 6.19 | 78.34 | 1012.84 | 317.58 | 2365.67 |
| Sri Lanka | LKA | LMIC | WHO-CHOICE | 14.34 | 3.36 | 38.91 | 493.5 | 150.87 | 1168.7 |
| Lesotho | LSO | LMIC | WHO-CHOICE | 4.76 | 1.02 | 13.43 | 112.5 | 33.63 | 269.64 |
| Morocco | MAR | LMIC | WHO-CHOICE | 12.79 | 2.73 | 36.26 | 413.11 | 131.06 | 958.69 |
| Moldova | MDA | UMIC | WHO-CHOICE | 11.85 | 2.41 | 34.34 | 286.47 | 90.31 | 667.1 |
| Madagascar | MDG | LIC | WHO-CHOICE | 3.14 | 0.69 | 8.79 | 58.14 | 17.42 | 139.22 |
| Maldives | MDV | UMIC | WHO-CHOICE | 22.4 | 4.52 | 65.12 | 734.1 | 231.49 | 1709.27 |
| Mexico | MEX | UMIC | WHO-CHOICE | 36.78 | 8.23 | 101.98 | 1523.26 | 465.12 | 3609.66 |
| Marshall Islands | MHL | UMIC | WHO-CHOICE | 7.85 | 1.68 | 22.2 | 181.34 | 54.55 | 433.19 |
| North Macedonia | MKD | UMIC | WHO-CHOICE | 29.01 | 6.08 | 82.87 | 1045 | 318.6 | 2478.4 |
| Mali | MLI | LIC | WHO-CHOICE | 3.46 | 0.75 | 9.73 | 72.21 | 22.15 | 170.69 |
| Myanmar (Burma) | MMR | LMIC | WHO-CHOICE | 3.08 | 0.68 | 8.63 | 64.69 | 19.64 | 153.77 |
| Montenegro | MNE | UMIC | WHO-CHOICE | 36.17 | 7.31 | 105.07 | 1433.1 | 422.06 | 3462.64 |
| Mongolia | MNG | LMIC | WHO-CHOICE | 10.14 | 2.14 | 28.87 | 312.75 | 96.38 | 737.44 |
| Mozambique | MOZ | LIC | WHO-CHOICE | 3.93 | 0.81 | 11.29 | 72.6 | 22.8 | 169.42 |
| Mauritania | MRT | LMIC | WHO-CHOICE | 6.74 | 1.51 | 18.69 | 181.73 | 56.17 | 427.8 |
| Mauritius | MUS | UMIC | WHO-CHOICE | 40.6 | 9.3 | 111.3 | 1592.25 | 481.88 | 3791.39 |
| Malawi | MWI | LIC | WHO-CHOICE | 2.89 | 0.61 | 8.25 | 51.84 | 15.81 | 122.93 |
| Malawi | MWI | LIC | Literature: pooled | 21.84 | 18.61 | 22.17 | 137.28 | 121.45 | 153.1 |
| Malaysia | MYS | UMIC | WHO-CHOICE | 32.79 | 6.96 | 93.12 | 1332.14 | 414.01 | 3126.6 |
| Malaysia | MYS | UMIC | Literature: pooled | NA | NA | NA | 903.63 | 714.69 | 1036.83 |
| Namibia | NAM | UMIC | WHO-CHOICE | 16.41 | 3.4 | 47.16 | 510.51 | 155.27 | 1212.33 |
| Niger | NER | LIC | WHO-CHOICE | 2.3 | 0.47 | 6.67 | 35.8 | 11 | 84.52 |
| Nigeria | NGA | LMIC | WHO-CHOICE | 11.05 | 2.32 | 31.51 | 292.55 | 88.17 | 698.19 |
| Nicaragua | NIC | LMIC | WHO-CHOICE | 10.94 | 2.48 | 30.15 | 331.95 | 102.81 | 780.57 |
| Nepal | NPL | LMIC | WHO-CHOICE | 4.3 | 0.95 | 12.02 | 93.33 | 28.12 | 222.77 |
| Pakistan | PAK | LMIC | WHO-CHOICE | 8.08 | 1.73 | 22.85 | 211.9 | 64.62 | 502.52 |
| Peru | PER | UMIC | WHO-CHOICE | 22.46 | 5.45 | 59.88 | 792.8 | 230.61 | 1928.2 |
| Philippines | PHL | LMIC | WHO-CHOICE | 12.22 | 2.7 | 34.09 | 377.03 | 113.83 | 898.93 |
| Palau | PLW | UMIC | WHO-CHOICE | 43.39 | 9.64 | 120.72 | 1857.07 | 544.07 | 4499.54 |
| Papua New Guinea | PNG | LMIC | WHO-CHOICE | 8.12 | 1.64 | 23.62 | 198.69 | 59.86 | 474.26 |
| Paraguay | PRY | UMIC | WHO-CHOICE | 16.41 | 3.55 | 46.2 | 490.56 | 148.91 | 1166.2 |
| Russia | RUS | UMIC | WHO-CHOICE | 40.82 | 8.12 | 119.4 | 1839.06 | 519.47 | 4542.48 |
| Rwanda | RWA | LIC | WHO-CHOICE | 4.37 | 0.98 | 12.15 | 92.15 | 27.9 | 219.38 |
| Sudan | SDN | LIC | WHO-CHOICE | 16.24 | 3.32 | 46.89 | 423.79 | 121.71 | 1037.67 |
| Senegal | SEN | LMIC | WHO-CHOICE | 5.88 | 1.23 | 16.81 | 135.61 | 42.59 | 316.47 |
| Solomon Islands | SLB | LMIC | WHO-CHOICE | 9.02 | 1.77 | 26.57 | 236.33 | 71.04 | 564.8 |
| Sierra Leone | SLE | LIC | WHO-CHOICE | 4.86 | 1.07 | 13.55 | 91.96 | 29.07 | 213.84 |
| El Salvador | SLV | LMIC | WHO-CHOICE | 18.15 | 4.06 | 50.3 | 644.16 | 207.6 | 1481.92 |
| Serbia | SRB | UMIC | WHO-CHOICE | 28.93 | 6.5 | 80.05 | 1078.62 | 329.52 | 2555.32 |
| Sao Tome and Principe | STP | LMIC | WHO-CHOICE | 7.6 | 1.69 | 21.17 | 183.05 | 57.03 | 429.07 |
| Suriname | SUR | UMIC | WHO-CHOICE | 32.61 | 6.88 | 92.91 | 1032.12 | 321.91 | 2417.75 |
| Eswatini | SWZ | LMIC | WHO-CHOICE | 12.46 | 2.2 | 38.37 | 412.81 | 128.35 | 968.68 |
| Syria | SYR | LIC | WHO-CHOICE | 17.09 | 3.4 | 49.96 | 564.1 | 173.48 | 1331.58 |
| Chad | TCD | LIC | WHO-CHOICE | 4.3 | 0.96 | 11.92 | 92.04 | 28.2 | 217.71 |
| Togo | TGO | LIC | WHO-CHOICE | 3.65 | 0.79 | 10.26 | 68.4 | 22.92 | 153.94 |
| Thailand | THA | UMIC | WHO-CHOICE | 21.92 | 4.5 | 63.19 | 766.77 | 234.04 | 1817.41 |
| Thailand | THA | UMIC | Literature: pooled | 208.81 | 208.12 | 209.5 | 2777.43 | 2705.6 | 2849.25 |
| Tajikistan | TJK | LMIC | WHO-CHOICE | 4.34 | 1.01 | 11.84 | 103.99 | 33.02 | 241.2 |
| Turkmenistan | TKM | UMIC | WHO-CHOICE | 22.52 | 4.83 | 63.68 | 745.9 | 217.14 | 1813.4 |
| Timor-Leste | TLS | LMIC | WHO-CHOICE | 4.89 | 0.99 | 14.21 | 107.14 | 32.58 | 254.47 |
| Tonga | TON | UMIC | WHO-CHOICE | 15.52 | 3.29 | 44.12 | 409.81 | 123.22 | 979.24 |
| Tunisia | TUN | LMIC | WHO-CHOICE | 22.07 | 4.37 | 64.71 | 907.45 | 266.48 | 2195.92 |
| Turkey | TUR | UMIC | WHO-CHOICE | 31.58 | 6.84 | 88.87 | 1313.31 | 395.65 | 3134.92 |
| Tuvalu | TUV | UMIC | WHO-CHOICE | 8.42 | 1.92 | 23.16 | 200.61 | 59.29 | 483.82 |
| Tanzania | TZA | LMIC | WHO-CHOICE | 4.35 | 0.94 | 12.25 | 91.73 | 27.27 | 220.54 |
| Uganda | UGA | LIC | WHO-CHOICE | 4.21 | 0.86 | 12.15 | 88.47 | 26.96 | 209.86 |
| Ukraine | UKR | LMIC | WHO-CHOICE | 12.91 | 3.12 | 34.5 | 466.12 | 146.5 | 1087.29 |
| Uzbekistan | UZB | LMIC | WHO-CHOICE | 5.64 | 1.19 | 16.05 | 158.12 | 47.88 | 376.37 |
| St. Vincent & Grenadines | VCT | UMIC | WHO-CHOICE | 27.69 | 5.95 | 78.2 | 1037.6 | 305.24 | 2508.53 |
| Vietnam | VNM | LMIC | WHO-CHOICE | 10.7 | 2.27 | 30.43 | 296.5 | 94.67 | 685.65 |
| Vietnam | VNM | LMIC | Literature: pooled | 83.94 | 79.34 | 88.53 | 348.52 | 331.62 | 365.43 |
| Vanuatu | VUT | LMIC | WHO-CHOICE | 11.63 | 2.62 | 32.16 | 369.71 | 117.09 | 858.8 |
| Samoa | WSM | LMIC | WHO-CHOICE | 13.29 | 2.82 | 37.73 | 419.68 | 131.17 | 981.98 |
| Yemen | YEM | LIC | WHO-CHOICE | 8.01 | 1.81 | 22.09 | 212.45 | 67.03 | 494.5 |
| South Africa | ZAF | UMIC | WHO-CHOICE | 25.19 | 5.63 | 69.83 | 895.05 | 270.1 | 2134.56 |
| South Africa | ZAF | UMIC | Literature: pooled | 59.9 | 57.13 | 62.67 | 1409.66 | 1294.31 | 1525.01 |
| Zambia | ZMB | LIC | WHO-CHOICE | 4.31 | 0.99 | 11.78 | 97.66 | 29.49 | 232.81 |
